# PRE-CISE: A PRE-calibration Coverage, Identifiability, and SEnsitivity analysis workflow to streamline model calibration

**DOI:** 10.64898/2026.02.27.26346591

**Authors:** Valeria Gracia, Jeremy D. Goldhaber-Fiebert, Fernando Alarid-Escudero

## Abstract

**Purpose:** We introduce PRE-CISE, a pre-calibration workflow integrating coverage analysis, local sensitivity, and collinearity diagnostics to streamline model calibration. We demonstrate PRE-CISE’s benefits using two testbeds (a Sick-Sicker Markov model and an SIR transmission model) and a COVID-19 case study.

**Methods:** PRE-CISE uses coverage analysis to verify that model outputs generated from the prior distribution span calibration targets, followed by local sensitivities to quantify parameter influence, guiding the resizing of prior bounds to improve coverage. Identifiability is assessed via collinearity analysis; large indices indicate practical nonidentifiability. Bayesian calibration was used: for the testbed models (3 and 2 parameters, respectively) to match their targets; for the COVID-19 model (11 parameters) to match daily confirmed incident cases.

**Results:** Coverage analyses flagged initial misfits; local sensitivities identified that the Sick-to-Sicker transition probability has a greater effect on model outputs, and resizing its prior distribution bounds improved coverage. Collinearity analyses indicated that using multiple calibration targets over different time points enabled identification of all three parameters. For the SIR testbed, collinearity indices declined as target points accumulated, crossing the identifiability threshold once the data spanned the epidemic peak. In the COVID-19 model, local sensitivity analyses prioritized time-varying detection rates and contact-reduction effects, reducing the search space. Daily incident case calibration targets yielded collinearity indices below practical thresholds for all parameter combinations, whereas indices for weekly calibration targets were larger. PRE-CISE achieved greater calibration efficiency gains for denser posterior sampling and more complex models.

**Conclusions:** PRE-CISE provides a practical, transparent pathway that helps modelers refine prior distribution bounds and calibration targets before intensive calibration, improving uncertainty reporting and strengthening the reliability of model-based health policy analyses.

## Introduction

Health policy models synthesize diverse empirical evidence and expert knowledge to project the long-term benefits and harms of observed and yet-to-be-observed interventions [1–3]. By integrating disease natural history and intervention effects, these models support policy makers as they weigh trade-offs, allocate resources, and design strategies under uncertainty. While some models parameters can be obtained or estimated from individual-level data or obtained from the literature [1, 2], other parameters are not directly observable or are difficult to measure for ethical, logistical, or feasibility reasons — for example, rates governing unobserved transitions, transmission probabilities, or time-varying detection processes — necessitating calibration to ensure model outputs align with credible calibration targets [4–6].

Model calibration is often computationally intensive, requiring repeated simulations across high-dimensional parameter spaces [4, 7, 8]. Efficient calibration depends on defining a well-informed region of the parameter space within which to search for a well-fitting subregion [5]. A common and consequential challenge is nonidentifiability, wherein multiple distinct parameter sets fit the calibration targets equally well [9]. If not addressed, nonidentifiability can undermine decision-making because different well-fitting regions may yield divergent projections and, in turn, different optimal strategies or recommendations [6]. Recognizing, diagnosing, and addressing nonidentifiability as part of the pre-calibration (PRE-CISE) and calibration process is therefore essential to ensure robust, policy-relevant conclusions.

Techniques from applied mathematics and inverse modeling, such as sensitivity analysis and collinearity indices, are applicable to the calibration of health decision models and can potentially improve calibration efficiency and reliability [9–11]. Sensitivity analysis allows researchers to quantify the influence of each model parameter on its projected outcomes [10, 12]. The collinearity index serves as a diagnostic tool to assess the degree of nonidentifiability in model calibration [9, 10, 12]. Together, these tools can guide the specification of the prior parameter distribution to enable a more efficient calibration search, indicate which additional or alternative calibration targets (and data granularity) would help reduce or eliminate nonidentifiability — for instance, combining multiple calibration targets or using higher-resolution data such as daily rather than weekly observations — and generally reduce computational burden.

Our paper operationalizes these techniques in a practical workflow that precedes and informs calibration, improving the definition of the parameter space, enhancing identifiability, and streamlining computation.

We illustrate the approach with three examples: a four-state Sick-Sicker cohort Markov model and a Susceptible-Infectious-Recovered (SIR) transmission model as testbeds, and an applied, age-structured, multicompartment COVID-19 transmission model previously used to study the Mexico City Metropolitan Area [13]. Across these examples, we demonstrate how coverage, sensitivity, and collinearity analyses can be used to set and adjust priors, select and combine calibration targets, anticipate and diagnose non-identifiability, and ultimately achieve more efficient calibration that supports clearer, more reliable policy recommendations.

## Methods

We introduce a pre-calibration workflow comprising three analyses, described below, and apply them to three illustrative models. After PRE-CISE, we implement Bayesian calibration in a four-state Sick-Sicker Markov model (calibrating to prevalence, survival, and Sick-Sicker proportions at multiple time points), in an SIR transmission model (calibrated to daily incident infections), and in an age-structured COVID-19 transmission model for the Mexico City Metropolitan Area (calibrating to incident case counts), including an examination of how target resolution affects identifiability and calibration performance.

### Coverage analysis

A coverage analysis aims to verify that when the model is evaluated on samples from the prior parameter space, it can generate outputs that span the calibration targets, thereby preventing wasted computation in infeasible search regions of the space. In practical terms, it asks whether predictions generated with samples drawn from the prior produce interquantile ranges that “cover” the calibration targets across all outcomes and time points of interest. This diagnostic does not actually identify parameters that fit the targets; instead, it confirms that the prior distribution is broad and well-centered, allowing for the simulation of calibration targets while remaining scientifically plausible.

To conduct the coverage analysis, we first specify prior distributions for all calibrated parameters using current evidence or domain knowledge (e.g., from previous empirical or modeling studies). Depending on parameter type, these may be uniform distributions with lower and upper bounds or parametric priors whose 95% intervals reflect plausible values (e.g., logit or beta for probabilities, lognormal for rates or hazards). We then draw a sample of *N* parameter sets from the prior distribution and run the model for each set to produce the corresponding outputs. For each calibration target, we summarize the predicted outputs across simulations using an interquantile range, often 95%, and compare these intervals to the calibration targets. If calibration targets fall outside these intervals or are only marginally covered, we iteratively resize or shift the relevant prior distribution’s bounds. We use local sensitivity analysis, as described below, to expand or re-center the prior parameter space, ensuring that adjustments stay consistent with biological and clinical plausibility. This process is repeated until visual and quantitative inspection indicate satisfactory coverage across all calibration targets, at which point the refined prior distribution bounds serve as inputs to subsequent sensitivity, identifiability, and calibration steps.

### Local sensitivity analysis

Local sensitivity analysis quantifies how small, local changes in each calibrated parameter affect the model outputs used as calibration targets. We implement a one-at-a-time (OAT) perturbation approach around a baseline input parameter vector (set at the mean or midpoint of the current prior bounds). For each parameter in turn, we perturb it by a small proportion while holding others fixed and re-simulate the model to estimate the derivative of each output with respect to that parameter. To facilitate comparison across heterogeneous parameters and outcomes, we scale these derivatives to obtain elasticities (the proportional change in output per proportional change in the parameter), assembling them into a sensitivity matrix, whose rows correspond to model outputs (including time-indexed points for time series) and whose columns correspond to parameters:

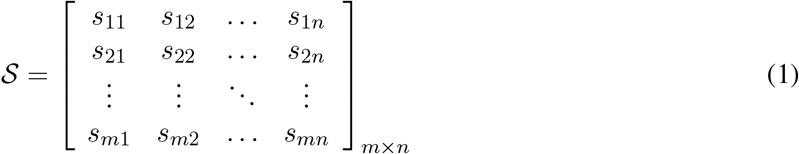

whose (*i, j*)-th element *s*_*ij*_ is defined as

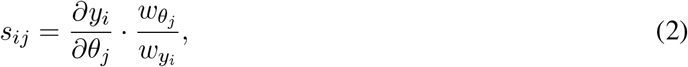

where *y*_*i*_ is the output variable *i*, for *i* = 1, …, *m*; *θ*_*j*_ is the parameter *j*, for *j* = 1, …, *n*; *w*_*yi*_ is the scaling of variable *y*_*i*_, typically set equal to the *i*-th model output evaluated at *θ*, the point at which the perturbation is applied; and *w*_*θj*_ is the scaling of parameter *θ*_*j*_, typically set to the value of parameter *j* at which the perturbation is applied. We summarize each parameter’s overall influence using norms of its elasticity column (e.g., *L*_1_ = ∑|*s*_*ij*_|*/n*, and 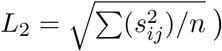, and we record the sign of the elasticities to indicate the direction of effect. A larger norm (*L*_1_ or *L*_2_) indicates a greater influence of the parameter on the model output [12].

### Using local sensitivity values to improve coverage

We use local sensitivity analysis to directly inform and improve coverage before computationally costly calibration, specifically to select key parameters that must have their prior distribution bounds adjusted to better cover the calibration targets. When coverage analysis shows that specific calibration targets are under- or over-predicted, the elasticity magnitudes identify which parameters most strongly control those outputs, and the elasticity signs indicate how to shift their priors. Based on this, we adjust the priors, always within biologically and clinically plausible ranges.

Using the sensitivity matrix, we determine how much and in which direction each calibrated parameter should be adjusted to produce an output change that covers the calibration targets. We then use this percentage change in outputs to refine the prior distribution bounds of the calibrated parameters, providing better coverage of the calibration targets. From Equation (2), we can write:

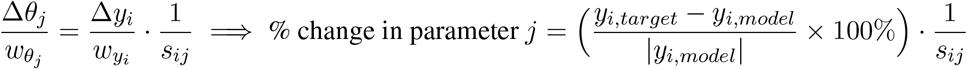

For each model output, we first estimate the percentage change in the calibrated parameter. This percentage change is then used to adjust the bounds of the prior distribution, resulting in revised bounds for each model output. To determine the final prior distribution bound, we combine the revised bounds using a predefined criterion (minimum, maximum, or mean), depending on the desired level of coverage conservatism. For example, if the calibrated parameter has a negative effect on the outputs, *s*_*ij*_ *<* 0, and the model is overestimating the outputs, Δ*y*_*i*_*/w*_*yi*_ *>* 0, then decrease the prior distribution bounds (Figure 1).

**Figure 1:**
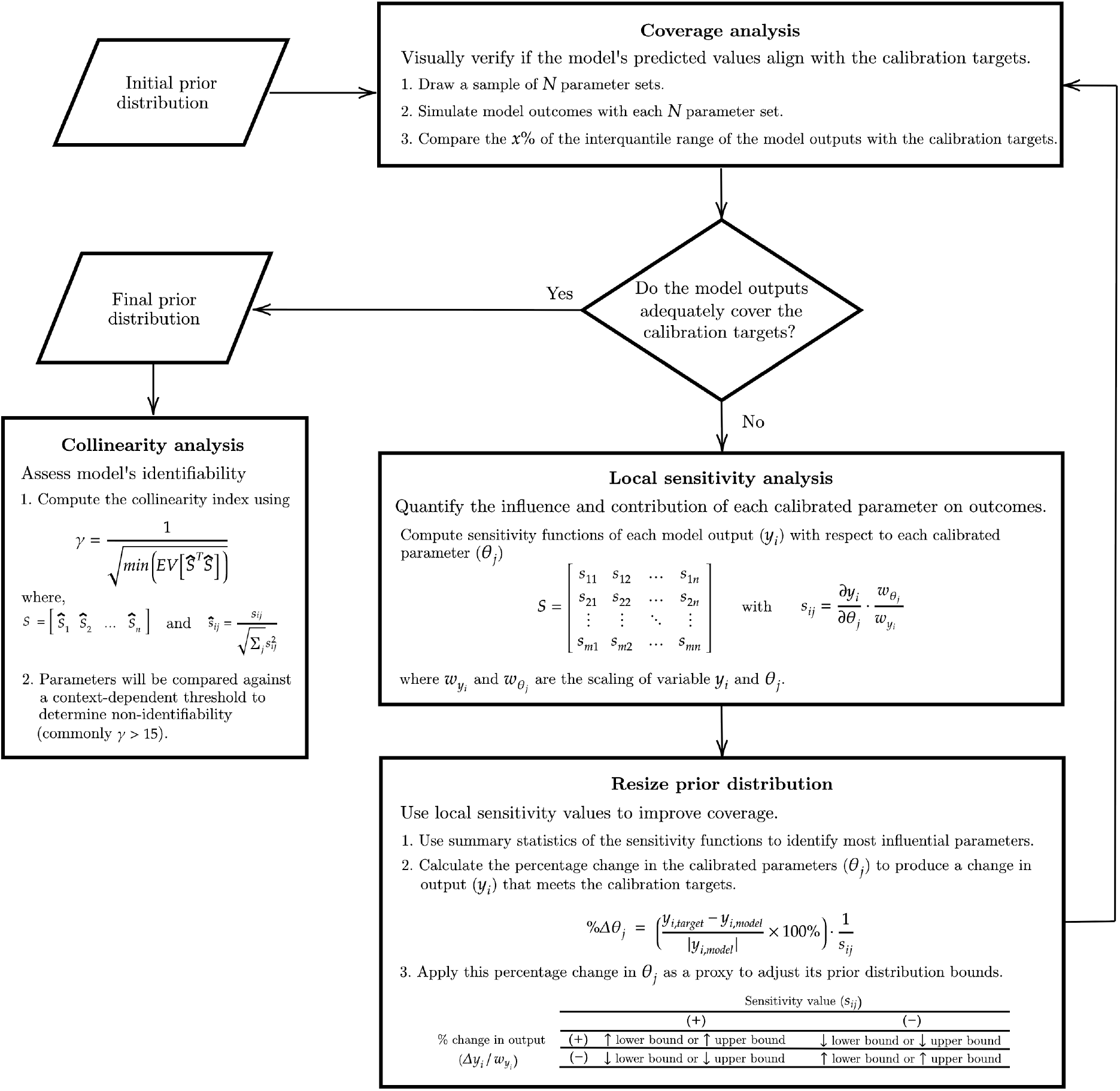
Pre-CISE workflow

If multiple outputs are uncovered, we prioritize the influential parameters that simultaneously improve several calibration targets in coordinated directions. Parameters with negligible influence can be given tighter bounds to reduce the dimensionality of the search space, provided the revised range remains plausible, and the posterior is checked for truncation. After each adjustment, we rerun coverage analysis to verify progress and repeat the sensitivity-guided refinement until the simulated interquantile ranges satisfactorily cover all calibration targets.

We recommend a conservative approach (i.e., generously widening bounds and cautiously narrowing them) at each iteration to reduce the risk of truncated posteriors during calibration. Since coverage iterations are relatively quick, the cost of proceeding cautiously is low.

### Collinearity analysis

Collinearity analysis leverages the local sensitivity matrix to quantify the degree of linear dependence among parameters with respect to the chosen calibration targets, providing a diagnostic for identifiability prior to full calibration.

After assembling the sensitivity matrix from scaled elasticities, we evaluate parameter identifiability by computing a collinearity index based on the smallest eigenvalue of the product of the columns of the sensitivity matrix corresponding to the parameters in the set, and its transpose:

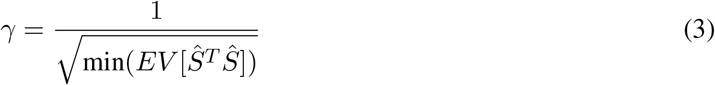

where,

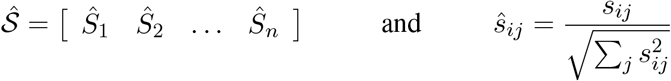

Intuitively, when columns of the sensitivity matrix (*Ŝ*_*i*_) are nearly linearly dependent, multiple parameter combinations can produce indistinguishable changes in the outputs, and the collinearity index becomes large. Following the inverse modeling literature [9–11, 14], thresholds around 15 indicate practical non-identifiability: parameter sets with indices above this level cannot be uniquely recovered from the available calibration targets. By repeating this calculation across different subsets of parameters and target sets, we characterize which combinations are identifiable and how identifiability changes with the number and nature of calibration targets.

We use the collinearity analysis to diagnose nonidentifiability and guide model refinement and data selection. First, the analysis reveals which parameter combinations can be estimated from a given target or combination of targets, holding the remaining parameters at their baseline values [9]. This depends on the information in the targets rather than on their number: a target observed at *T* time points contributes *T* rows to *S*, so it can jointly identify at most *T* parameters, and fewer when the time profiles of their sensitivities are nearly proportional. A single parameter is never limited by collinearity (*γ* = 1 by construction) but requires the target to be sufficiently sensitive to it. Adding targets or combining outcomes across multiple time points can reduce collinearity and enable identification of more parameters. Second, it highlights the role of data resolution: higher-resolution targets (e.g., daily time series rather than weekly aggregates) can improve identifiability, though whether the improvement is decision-relevant depends on the model and the target set. When the collinearity index signals nonidentifiability, we could add or reweight targets, increase temporal resolution, fix or tightly constrain low-influence parameters, or reparameterize the model to reduce the number of calibrated parameters. If, despite these steps, calibration remains nonidentifiable, analysts must report the existence of multiple, distinct well-fitting solutions or, ideally, provide a posterior distribution that explicitly reflects the nonidentifiability. Transparent reporting of this uncertainty enables decision makers to understand the range of plausible parameterizations and the potential implications for policy recommendations [6].

### PRE-CISE workflow

First, we define the prior distribution for the parameters we wish to calibrate and run a coverage analysis. We then conduct a sensitivity analysis to determine which parameters most influence the model outputs and use the sensitivity values to refine the prior distribution bounds to improve coverage. This process is repeated until we (visually) confirm that the model-predicted outputs cover the calibration targets. Finally, we assess the model’s identifiability using a collinearity analysis, comparing the collinearity index of different combinations of parameters and targets (Figure 1).

### Sick-Sicker Testbed

We illustrate PRE-CISE using the Sick-Sicker cohort state-transition model (cSTM), a previously published four-state model [15, 16] in which a hypothetical cohort of healthy individuals can transition to Sick, Sicker, or Dead states. Sick individuals can either get worse, progressing to a Sicker state, or recover and become healthy again. Healthy individuals are at risk of dying from all causes, and those in the Sick and Sicker states face a disease-specific mortality that increases with severity.

We calibrated three parameters of the Sick-Sicker cSTM: the probability of transitioning from Sick to Sicker and the hazard ratios for increased mortality for the Sick and Sicker states. We used the survival probability, prevalence, and the proportion of those with the disease who are in the Sick state (vs. Sicker) as calibration targets at 10, 20, and 30 years. We generated the calibration targets from a ground truth, using a known set of model input parameter values. We then used the Sick-Sicker cSTM to simulate 1,000 individuals over 30 years using a microsimulation [17]. We used the microsimulation output as data to produce three sets of targets: survival, disease prevalence, and the proportion of sick individuals in the Sick state. For the first two target types, we had annual calibration targets, whereas for the third, we had data only at 10, 20, and 30 years of follow-up.

We compared the model outputs to the calibrated targets using a likelihood function, assuming the targets follow normal distributions with means derived from the model-predicted outputs. We defined uniform prior distributions for all calibrated parameters. We used the Incremental Mixture Importance Sampling (IMIS) algorithm [18, 19] to calibrate the unknown parameters. Our implementation of the algorithm begins by sampling 10,000 values from the prior distributions, then iteratively samples 1,000 values over a maximum of 30 iterations, producing a posterior distribution from which we take 10,000 samples.

### SIR Testbed

We illustrate our approach using a well-known dataset describing an influenza outbreak at a British boarding school in early 1978 [20–22]. During this outbreak, all boys at the school were affected, and the epidemic was extinguished within two weeks. We modeled transmission with a deterministic Susceptible-Infectious-Recovered model in a closed population of *N* = 763 individuals:

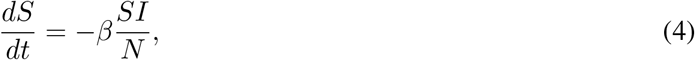

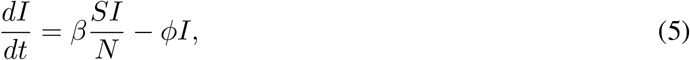

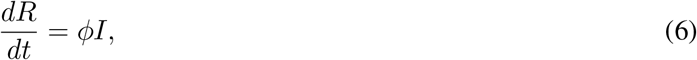

where *S, I*, and *R* denote the number of susceptible, infectious, and recovered individuals, *β* is the transmission rate per day, *ϕ* is the recovery rate per day, and *N* = *S* +*I* +*R* is constant. We initialized the system with a single index case, *I*(0) = 1, *S*(0) = *N* − 1, and *R*(0) = 0. The two calibrated parameters were *β* and *ϕ*; the basic reproduction number is *R*_0_ = *β/ϕ* and the initial exponential growth rate is *r* = *β −ϕ*. We use *ϕ* to denote the recovery rate instead of the usual *γ* to prevent confusion with the collinearity index. Model-predicted daily incident infections 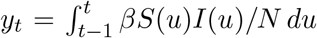, were compared with the observed daily counts using a negative binomial likelihood, with mean parameters derived from the model outcomes, and a dispersion parameter of 5. We assigned uniform priors to both calibrated parameters based on epidemiological plausibility. We calibrated the model using the IMIS algorithm. The algorithm was initialized by drawing 10,000 parameter sets from the prior distributions, after which 1,000 samples were iteratively drawn for up to 20 iterations, yielding a posterior distribution from which we drew 10,000 samples.

### COVID-19 Case Study

We present how PRE-CISE improves calibration in a realistic age-structured, multi-compartment, Susceptible-Exposed-Infected-Recovered (SEIR) model of COVID-19 that includes both community and household transmission in the Mexico City Metropolitan Area (MCMA) [13].

We calibrated the model using daily confirmed COVID-19 cases in the MCMA from February 24 to December 7, 2020 [23]. The model included 11 unknown parameters, such as transmission rates within the community and households, as well as parameters governing time-varying daily detection rate and the effectiveness of previous non-pharmaceutical interventions (NPIs) in the MCMA, modeled as a proportional reduction in effective contacts.

We compared the model results with empirical data using a negative binomial likelihood, with mean parameters derived from the model outcomes, and a dispersion parameter of one to account for overdispersion in the target data. Uniform prior distributions for the model input parameters with defined ranges were determined based on existing evidence, epidemiological theory, and plausibility considerations. We used the IMIS algorithm for model calibration. Our application involved sampling 10,000 parameter sets from the prior distributions, followed by iterative sampling of 1,000 values for up to 55 iterations. We drew a sample of 1,000 values from the joint posterior distribution of the calibrated model parameters to represent their uncertainty.

## Results

### Sick-Sicker Testbed

Figure 2-A illustrates PRE-CISE’s step-by-step iterative coverage after resizing and recentering the prior distributions using the local sensitivity results. The Supplementary Material presents an example with a different starting prior distribution (eFigure 3).

**Figure 2:**
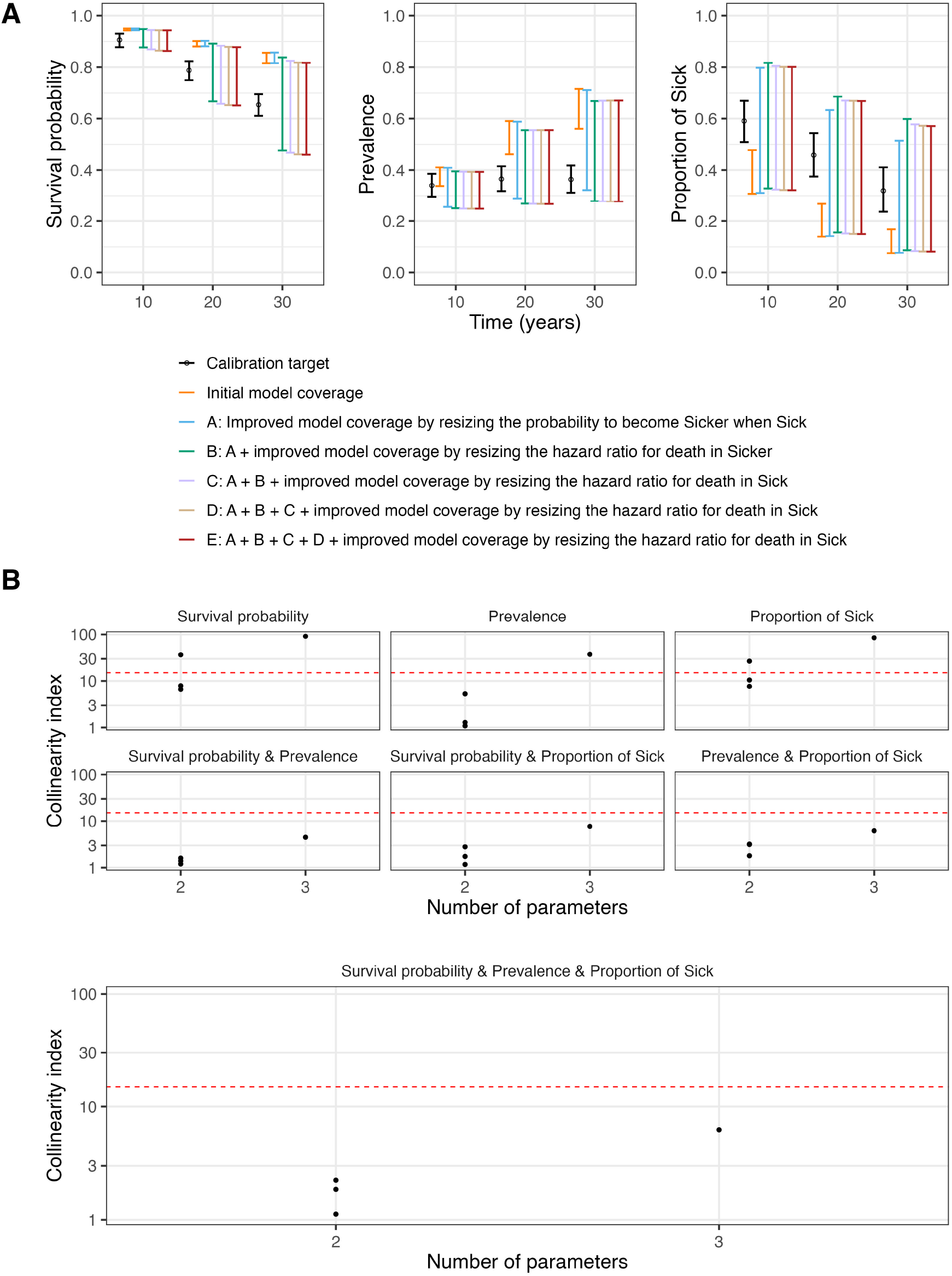
PRE-CISE diagnostics for the Sick-Sicker testbed model. (A) Coverage analysis after iteratively resizing and recentering the prior distributions using the local sensitivity results; the model intervals represent the intermediate stages of this iterative process. (B) Collinearity index for all parameter combinations. Black dots represent the values for different parameter combinations. For two-parameter sets, up to three parameter pairs are shown; overlapping points may appear as fewer. For three-parameter sets, there is a single combination, shown as one point. The red dashed line shows the highest value considered identifiable, which is 15 in this case.

The initial coverage analysis reveals that the 95% interquantile range of the model’s outputs, evaluated in the prior parameter space, is substantially outside the calibration targets’ range, overestimating prevalence and survival probability while underestimating the proportion of sick individuals in the Sick state.

A local sensitivity analysis around the mean of these initial prior distributions quantifies the overall influence of each calibrated parameter on the model outputs (eFigure 1-A). An evaluation of the summary statistics of the sensitivity matrix shows that the transition probability from Sick to Sicker has the greatest impact on both the prevalence and the proportion of sick individuals in the Sick state, as evidenced by a larger L2 norm, followed by the hazard ratio of death of sicker and sick individuals (Table 1).

**Table 1:** Summary of the initial local sensitivity analysis.

| Parameter | Value | Mean | $L_1$ norm | $L_2$ norm |
| --- | --- | --- | --- | --- |
| *Sick-Sicker Testbed* |  |  |  |  |
| Probability to become Sicker when Sick | 0.15 | -0.216 | **0.449** | **0.621** |
| Hazard ratio of death in Sick vs Healthy | 1.35 | -0.016 | 0.016 | 0.021 |
| Hazard ratio of death in Sicker vs Healthy | 1.50 | -0.012 | 0.028 | 0.033 |
| *SIR Testbed* |  |  |  |  |
| Transmission rate | 1.90 | -0.72 | **3.49** | **3.90** |
| Recovery rate | 0.65 | -1.16 | 1.16 | 1.28 |
| *COVID-19 Case Study* |  |  |  |  |
| Transmission probability per effective contact per day |  |  |  |  |
| Community | 0.21 | 14.48 | 56.03 | 84.69 |
| Household | 0.25 | 38.49 | 90.56 | 112.10 |
| Effectiveness of NPI as a proportional reduction in effective contacts |  |  |  |  |
| (2020/03/25 to 2020/04/21) | 0.50 | 14.37 | 94.61 | 152.31 |
| (2020/04/21 to 2020/05/23) | 0.45 | 42.16 | 50.26 | 86.69 |
| (2020/05/23 to 2020/08/22) | 0.45 | 20.19 | 41.36 | 66.49 |
| (2020/08/22 to 2020/10/31) | 0.49 | 1.11 | 4.20 | 11.9 |
| (2020/10/31 to 2020/12/07) | 0.59 | 0.33 | 0.33 | 1.09 |
| Time-varying daily detection rate generalized logit parameters |  |  |  |  |
| Initial daily rate | 0.10 | -21.57 | 43.13 | 56.28 |
| Final daily rate | 0.16 | 19.08 | 43.08 | 58.42 |
| Rate of change between the initial and final detection rates | 0.76 | 18.60 | 149.33 | 243.19 |
| Time of the sigmoid's midpoint | 69.65 | -18.47 | **183.90** | **358.52** |

Note: $L_1 = \sum |S_{ij}|/n$ , $L_2 = \sqrt{\sum (S_{ij}^2)/n}$
Bold values indicate the largest $L_1$ and $L_2$ norms within each model block (i.e., the most influential calibrated parameter).

We used the sensitivity results to iteratively resize the prior parameter space, beginning with the most influential parameter, the probability of becoming Sicker when Sick. Prevalence needed to decrease and has a positive elasticity, while the proportion of sick individuals in the Sick state needed to increase and has a negative elasticity. We therefore applied the maximum overall percentage change to reduce the lower bound of its prior distribution, keeping the revised range biologically and clinically plausible.

Repeating the sensitivity analysis around the mean of the updated bounds showed that the hazard ratios for death in the Sicker and Sick states both have a negative impact on survival and prevalence, with a greater effect in the Sicker state and a negligible effect on the proportion of Sick. Because survival was overestimated, we raised their upper bounds. We iterated this procedure until the model outputs visually covered all calibration targets (Figure 2-A).

Figure 2-B shows the collinearity analysis evaluated on the mean of the prior distribution after resizing the bounds. This analysis combines multiple calibration targets to assess when different parameter combinations are identifiable. We found that a single calibration target identifies only some two-parameter combinations, depending on the target and the identifiability threshold. Specifically, when prevalence is used as the sole target, all two-parameter collinearity indices fall below 10. When the proportion of sick individuals in the Sick state is used alone, only one of the three two-parameter indices falls below 10, and two fall below 20. When survival is used alone, two of the three two-parameter indices fall below both the 10 and 20 cutoffs. When any single target is used, the three-parameter indices fall above the 10 and 20 cutoffs. When any two targets are used, all indices fall below 10, indicating that recovering all three calibrated parameters requires at least two calibration targets.

### SIR Testbed

The initial PRE-CISE diagnostic confirmed that the prior parameter space used as input for the coverage analysis encompassed the 95% intervals for all calibration targets, although the prior bounds were excessively wide (Figure 3-A). Local sensitivity analysis around the prior means identified the transmission rate as the most influential parameter, followed by the recovery rate (eFigure 4-A). The transmission rate exhibited positive elasticity during the early epidemic period, reversing sign as the epidemic subsided, while the recovery rate showed consistently negative elasticity throughout. These elasticity estimates were then used to refine the bounds of the prior distribution.

**Figure 3:**
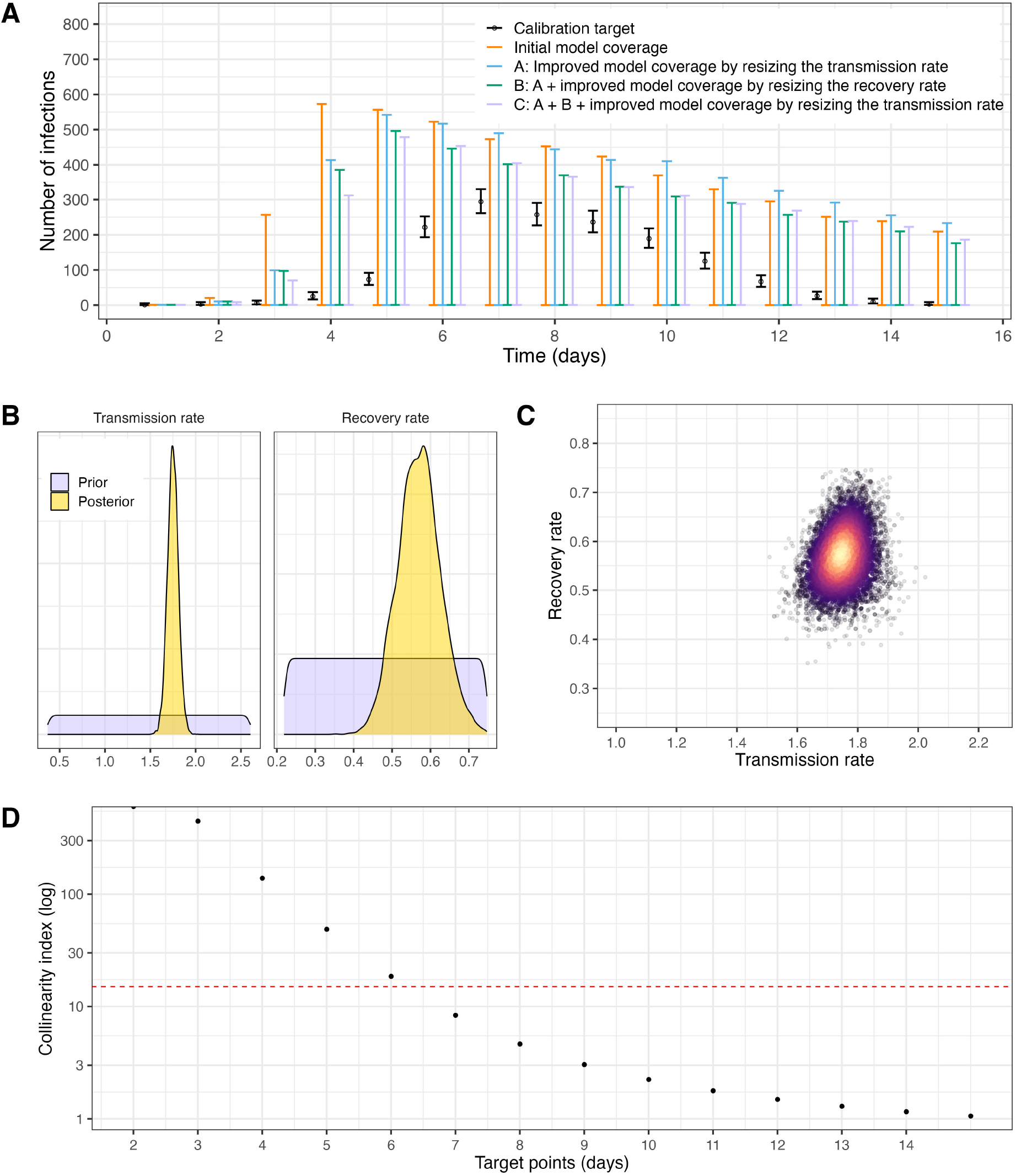
PRE-CISE diagnostics for the SIR testbed model. (A) Coverage analysis after iteratively resizing and recentering the prior distributions using the local sensitivity results; the model intervals represent the intermediate stages of this iterative process. (B) Prior and posterior marginal distributions of calibrated parameters. (C) Joint distribution of calibrated parameters. (D) Collinearity index as target points are sequentially added. The red dashed line shows the highest value considered identifiable, which is 15 in this case.

We began by resizing the prior bounds of the transmission rate given its influence on the model output. Using its sensitivity results we increased the lower bound and decreased the upper bound of the prior distribution, with adjustments derived from output-specific changes aggregated across days, while ensuring that the revised range remained epidemiologically plausible. The recovery rate bounds were then resized through an updated OAT sensitivity analysis around the mean of the updated input parameter space with the resulting elasticity estimates used to calculate proportional adjustments to both bounds. We repeated this process until visual assessment confirmed satisfactory coverage. Calibration was then performed using this updated prior distribution.

Successful calibration was confirmed by comparing the prior and recovered posterior distributions (Figure 3-B), examining their joint posterior distribution (Figure 3-C), and assessing the model fit against the calibration targets (eFigure 5).

Finally, to investigate how identifiability evolves as data accumulate, we computed collinearity indices by sequentially incorporating calibration target points. Figure 3-D shows that the collinearity index, *γ*, decreases as more time points become available for calibration. In this example, identifiability would be achieved after 6 to 7 days of outbreak data were available, assuming identifiability thresholds of *γ <* 20 and *γ <* 10, respectively. As the number of targets increases beyond this threshold, identifiability improves substantially. This pattern has a mechanistic interpretation. During the initial exponential phase, incidence is mainly determined by the net growth rate *r* = *β − ϕ*, so any pair (*β, ϕ*) preserving that difference generates near-identical trajectories. The columns of the sensitivity matrix become nearly linearly dependent, which the collinearity index is designed to detect. The two parameters become separately identifiable only once susceptible depletion bends the epidemic curve near its peak, which coincides with the collinearity index crossing the conventional threshold. The diagnostic therefore flags nonidentifiability for the correct structural reason, and resolves it once the data first contain the necessary information.

### COVID-19 Case Study

We applied PRE-CISE to the age-structured COVID-19 model to evaluate whether the prior parameter distributions and selected calibration targets were sufficient to support efficient and identifiable calibration. The coverage analysis (Figure 4) indicated that the predicted results from the model obtained from the initial priors generally covered the ranges of the calibration targets, but the implied parameter space was wider than necessary, generating many trajectories that were only weakly consistent with the targets. We therefore resized the prior bounds, tightening ranges for parameters with little support from the targets while preserving plausibility, to concentrate sampling on parameter regions that produced target-consistent behavior and to improve calibration efficiency.

**Figure 4:**
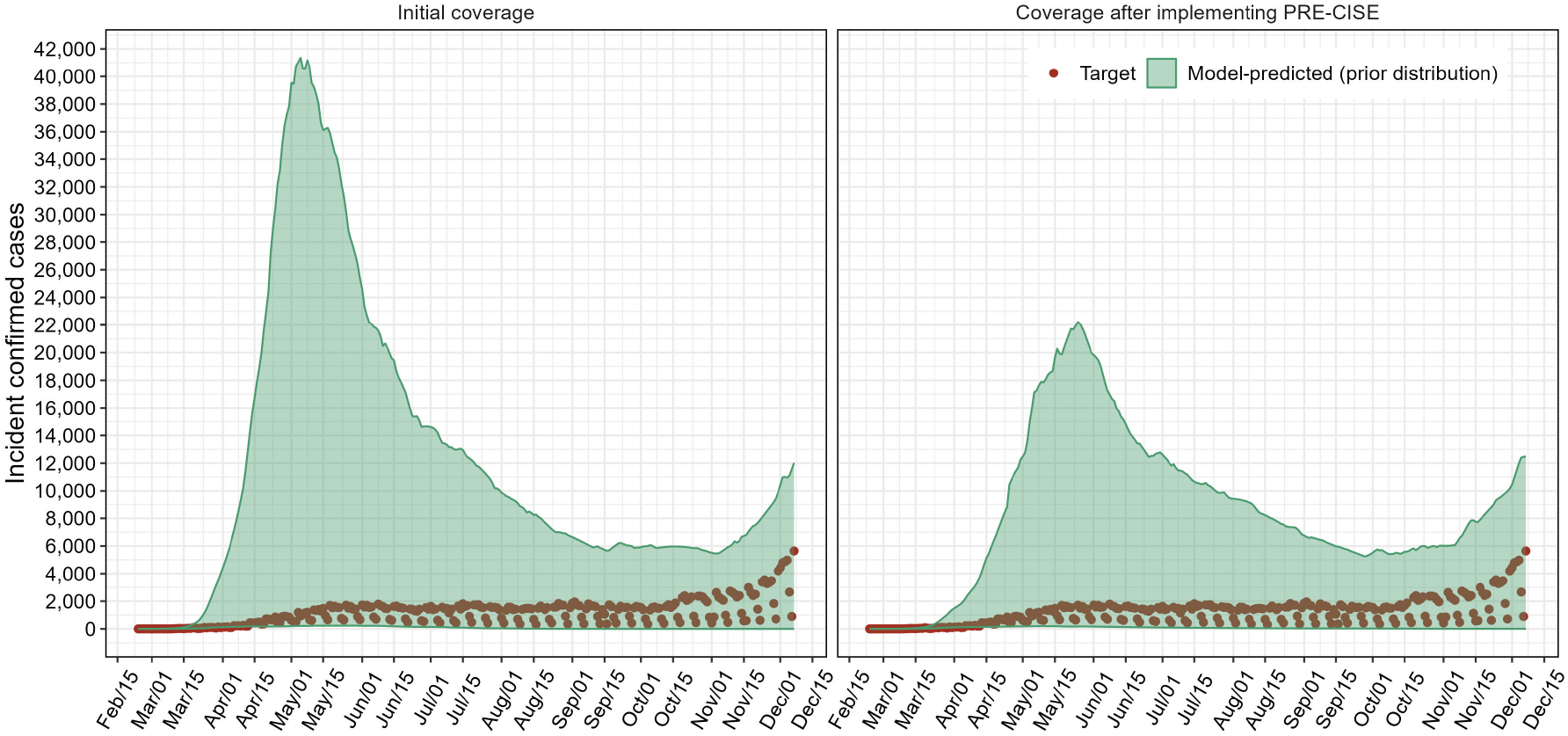
PRE-CISE diagnostics for the COVID-19 case study. (A) Coverage analysis using simulations drawn from the initial priors (left) and the resized priors after PRE-CISE (right). Shaded regions show the 95% prior predictive interval for daily incident cases; points show observed daily confirmed COVID-19 incident cases in the Mexico City Metropolitan Area (MCMA) from February 24, 2020, to December 7, 2020.

An OAT sensitivity analysis around the prior means showed that model-predicted incident cases were most responsive to parameters governing the time-varying daily detection rate, with secondary influence from parameters affecting the effectiveness of NPIs (proportional reduction in effective contacts) and the transmission probabilities (Table 1). These elasticity patterns provided a practical basis for prioritizing which parameters to refine prior to calibration and which outputs were most informative for distinguishing among parameter effects.

First, we adjusted the prior bounds for the parameter corresponding to the timing at which the detection rate lies between the lower and upper bounds. Because this parameter exhibited a negative average elasticity across outputs, we updated its lower bound by applying output-specific bound changes aggregated across outcomes, while ensuring that the revised range remained plausible. Using the updated input space, we repeated the sensitivity analysis and identified the initial NPI effectiveness (proportional reduction in effective contacts) as the next most influential parameter. This parameter also showed negative elasticity, so we increased its lower bound to a plausible change supported by the sensitivity results. Finally, we iteratively updated the upper bounds on the community and household transmission probabilities, which had positive elasticities, thereby tightening the space (primarily via the community transmission upper bound) until the coverage plots indicated that the revised priors generated trajectories that consistently spanned the calibration targets (Figure 4 and step-by-step process in eFigure 8). After implementing PRE-CISE, the reduced input parameter space lowered the total number of model evaluations required for calibration by 10% (41,600 versus 46,000; Appendix D), a figure that already includes the 3,600 evaluations consumed by PRE-CISE itself.

We then assessed identifiability via collinearity analysis of the sensitivity matrix. All parameter combinations yield indices below 4 for both daily and weekly targets, so identifiability in this model is unaffected by any cutoff between 10 and 20 (eFigures 7 and 9-B, respectively). Temporal aggregation therefore reduced the information available to distinguish parameters without, in this model, crossing into practical nonidentifiability. The SIR testbed (Figure 3-D) shows the same principle. Specifically, decreasing the number of informative target points causes the collinearity index to cross the threshold, indicating that target information content, rather than just nominal temporal resolution, determines identifiability.

## Discussion

We developed and demonstrated PRE-CISE, a pre-calibration workflow that integrates coverage, local sensitivity, and collinearity analyses to support efficient and identifiable calibration. We applied PRE-CISE to two testbeds, a four-state Sick-Sicker Markov and an SIR transmission model, and to an age-structured COVID-19 transmission model for the Mexico City Metropolitan Area, then calibrated all three using Bayesian methods. Across models, coverage analysis identified prior spaces that did not exhibit target-consistent behavior, elasticities identified parameters requiring substantial bound adjustments, and collinearity indices showed which parameter-target combinations were recoverable.

Prior predictive coverage and sensitivity analysis are established in the statistical and inverse modeling literature [9–11, 14], and identifiability diagnostics are recommended in calibration guidance [6]. They are rarely implemented, however, as a structured pre-calibration workflow in health decision modeling. PRE-CISE contributes an explicit sequence with decision rules for when to resize a prior bound, in which direction, and by how much, and for determining whether a given set of calibration targets can recover the parameters of interest. Hence, it bridges inverse-modeling methods to model-based health policy applications and emphasizes transparent reporting when nonidentifiability persists.

Beyond our three examples, PRE-CISE applies across model classes, including deterministic compartmental and differential equation models, microsimulations and discrete-event simulations, agent-based models, and hybrid mechanistic–statistical frameworks. We provide documented code for PRE-CISE steps (Appendix E); a model only needs to accept a vector of calibrated parameters and return outputs mapped to calibration targets. The workflow is also useful in building emulators and surrogate models. First, prior predictive coverage guides the placement and range of simulator runs. Second, local sensitivities support variable selection and dimensionality reduction. Finally, collinearity analysis identifies informative combinations of outputs and targets for defining loss functions. Iterating these steps with active learning [24] yields emulators that generalize better in the domain that matters for calibration.

In practice, PRE-CISE is broadly applicable across calibration contexts and benefits a range of decision-modeling stakeholders. The workflow can be combined with standard sampling designs, such as Latin hypercube or Sobol sequences and with calibration methods such as IMIS or Markov chain Monte Carlo. For analysts, it serves as a protocol for refining priors and anticipating identifiability problems before conducting an in-depth search. For decision-makers, it clarifies what can be learned from the data and provides criteria to evaluate the rigor of calibration. For journal editors and reviewers, it provides criteria to assess the rigor and transparency of calibration. For those involved in funding studies, collecting data, and maintaining repositories, it highlights the value of higher-resolution, complementary data in reducing nonidentifiability. For educators, it provides a foundation for standardizing good calibration practices across health modeling domains.

The PRE-CISE framework has some limitations. It relies on local, OAT sensitivities computed around a single baseline parameter vector, which keeps the workflow inexpensive and model-agnostic but carries additional costs. Perturbing parameters individually cannot reveal interactions, so two parameters that are jointly influential but individually flat will be underweighted. A local derivative can misrepresent behavior when an output responds nonlinearly across the prior range, so an elasticity estimated at the midpoint may not generalize to the bounds. And a collinearity index built from a local sensitivity matrix characterizes identifiability only near the baseline, potentially missing multimodality, curved ridges, or disconnected well-fitting regions. We therefore position this step as a screening diagnostic: it reliably detects the most common pre-calibration problems, but does not substitute for global sensitivity analysis or full posterior exploration in models with strong nonlinearity or interaction structure. The sensitivity matrix can be generalized by averaging Jacobians over prior draws or by replacing derivatives with variance-based measures such as Sobol indices, at additional computational cost.

A related concern is whether using prior predictive coverage to resize prior bounds constitutes using the data to inform the prior. The procedure neither shifts nor concentrates priors toward the data: narrowing excludes regions where model outputs lie far from the targets and have negligible likelihood, while widening prevents the prior from unjustifiably asserting that plausible values are impossible. This distinction addresses the reason for resizing, not bound placement. As shown in Appendix A, a bound from first-order elasticity can meet coverage requirements, yet exclude significant parameter values of non-negligible posterior support, because linearization underestimates how far the bound must move for a nonlinearly responding output. We therefore recommend widening the bounds generously, narrowing sparingly, and checking the posterior marginals for mass accumulating at a prior bound. Coverage shows that the prior predictive can cover targets, but whether bounds are scientifically plausible depends on domain knowledge.

Collinearity indices are interpreted against a heuristic threshold (commonly *γ ≈* 15)[9–11, 14] that is context-dependent and treated as a continuum of increasing nonidentifiability rather than a sharp boundary. We report all parameter-target combinations so readers can apply their own cutoff. Finally, our demonstrations use three models and a limited set of calibration targets; the workflow cannot resolve structural model misspecification or measurement error and adds computational steps before calibration.

PRE-CISE is simple to implement, model-agnostic, and reproducible. By calibrating with priors that reproduce targets and vetted parameter sets for identifiability, modelers avoid failures that would otherwise surface after extensive computation. Efficiency gains were modest for low-dimensional models with small posterior samples and larger for higher-dimensional models and denser posterior sampling, indicating that the benefit scales with the cost of the calibration it precedes. When identifiability cannot be achieved, PRE-CISE helps communicate uncertainty clearly to decision makers, encouraging the reporting of multiple solutions that fit well or a posterior distribution indicating nonidentifiability when a single definitive region cannot be determined.

## Data Availability

Code used to illustrate PRE-CISE on testbeds (Sick-Sicker Markov model and SIR transmission model) is available at

https://github.com/vgraciaol/PRE-CISE

