## Supplementary Material for "PRE-CISE: A PRE-calibration Coverage, Identifiability, and SEnsitivity analysis workflow to streamline model calibration"

### Appendix A

eFigure 1 shows local and global sensitivity analyses of the Sick-Sicker testbed model. Both methods consistently identify the probability of becoming Sicker when Sick as the dominant driver of prevalence and proportion of Sick, and the hazard ratio for death in Sicker as the dominant driver of survival probability, with the hazard ratio for death in Sick a distant third. The strong similarity between first-order and total Sobol' indices suggests that interactions between parameters are minimal, with each parameter mainly affecting the model outputs through its own effect.

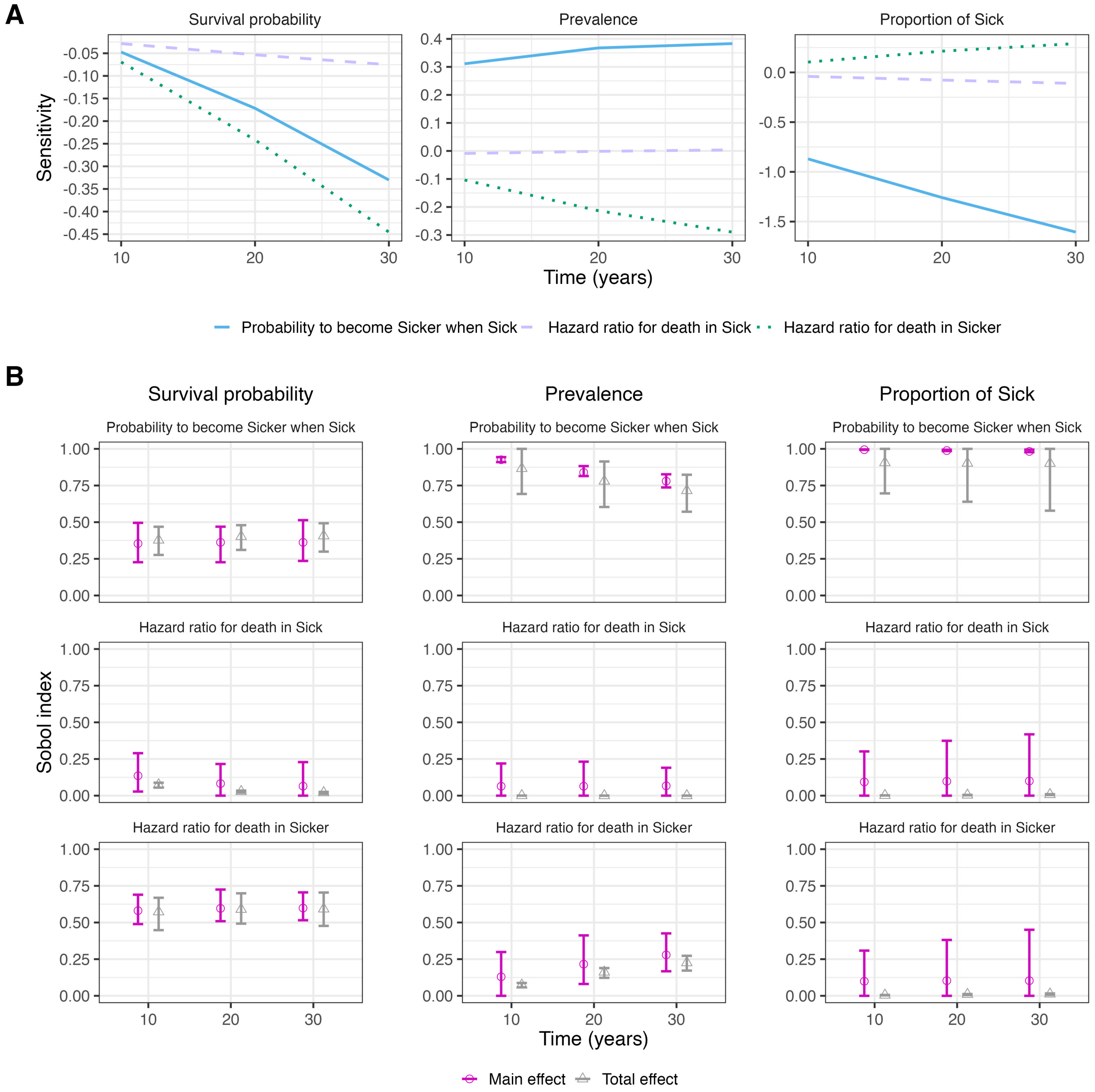


eFigure 1: Local and global sensitivity analysis of the Sick-Sicker testbed model. (A) Local sensitivity analysis evaluated at the mean of the prior distribution. (B) Global sensitivity analysis displays first-order Sobol indices (main effects), which quantify each parameter's contribution to output variance, and total indices (total effects) that account for parameter interactions. The bootstrap confidence interval bounds were manually restricted to the plausible range $[0, 1]$.

We next illustrate the impact of prior bound specification on calibration outcomes in the Sick-Sicker testbed model, using the hazard ratio for death in Sicker as an example. Both runs shown here widened the upper bound of this parameter's prior distribution from its initial value, but by different amounts: a less permissive adjustment yielded the interval $[1, 8.82]$, and a more permissive adjustment yielded $[1, 21.25]$. Both achieved reasonable prior predictive coverage (eFigures 2-A and 3-A), yet they produced notably different posterior distributions: under the less permissive bound the marginal posterior of the hazard ratio for death in Sicker is visibly truncated against its upper limit, whereas under the more permissive bound it is not (eFigures 2-B and 2-C and 3-B and 3-C). The mechanism provides guidance. Bound adjustments in PRE-CISE are based on a first-order (local) elasticity. When the model output reacts nonlinearly to a parameter across the prior range, this linear approximation underestimates the necessary bound movement, leading to a bound that can ensure coverage yet exclude parameter values with significant posterior support. Coverage analysis alone cannot detect this: it checks if the prior predictive covers the targets, but not whether the prior bounds include the region with significant likelihood. These are different criteria. We recommend generously widening bounds and narrowing them cautiously at each step. Importantly, after calibration, examine the posterior marginals for any mass accumulating at a prior bound. If truncation occurs, the boundary should be expanded again and the calibration redone. In our runs, the more permissive bounds increased total runtime by less than 3% while improving the effective sample size (Appendix D), so the cost of erring toward wider bounds is small relative to the risk of understating posterior uncertainty.

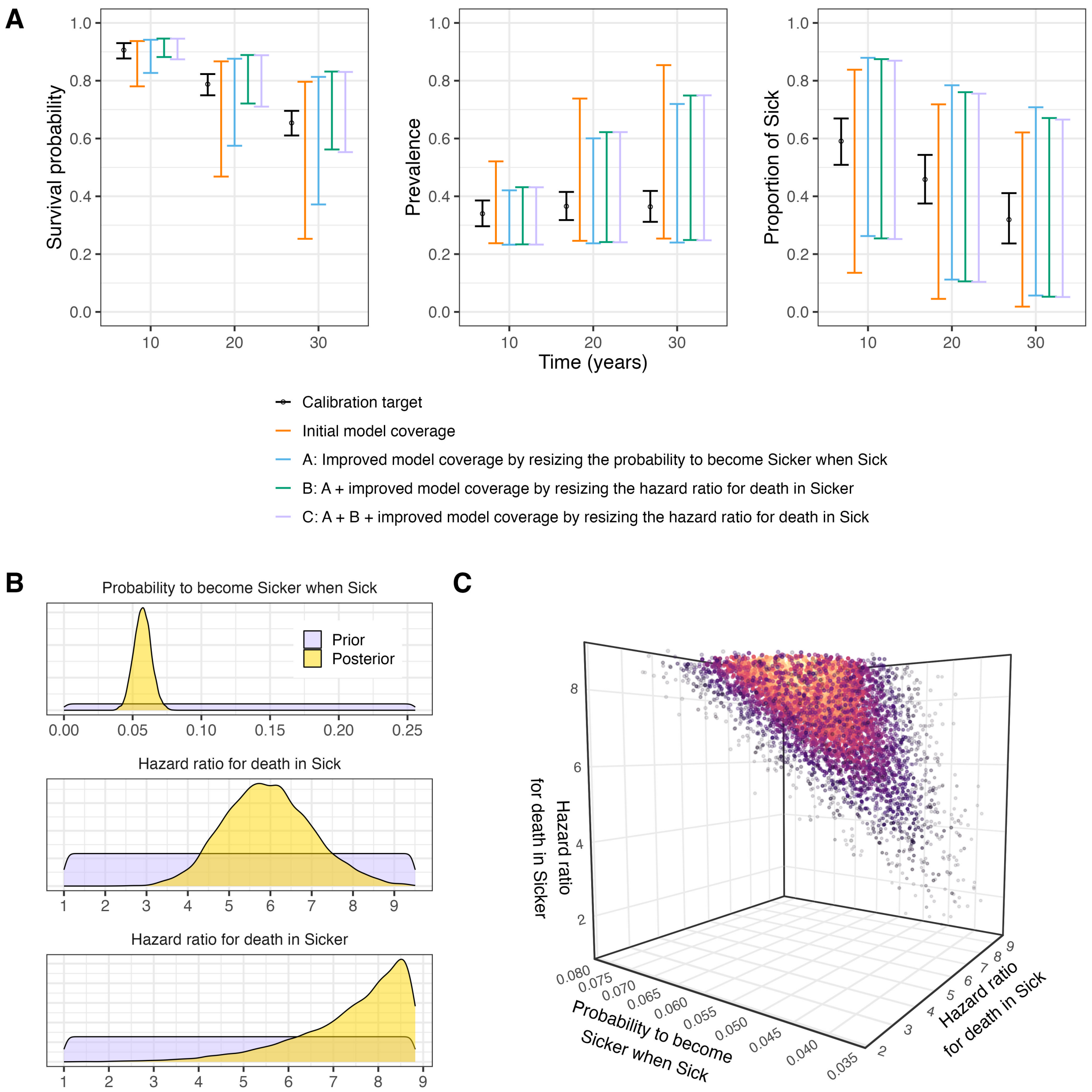


eFigure 2: Less permissive widening of the prior bounds in the Sick-Sicker testbed model. (A) Coverage analysis after iteratively resizing and recentering the prior distributions using the local sensitivity results; the model intervals represent the intermediate stages of this iterative process. (B) Prior and posterior marginal distributions of calibrated parameters. (C) Joint distribution of calibrated parameters.

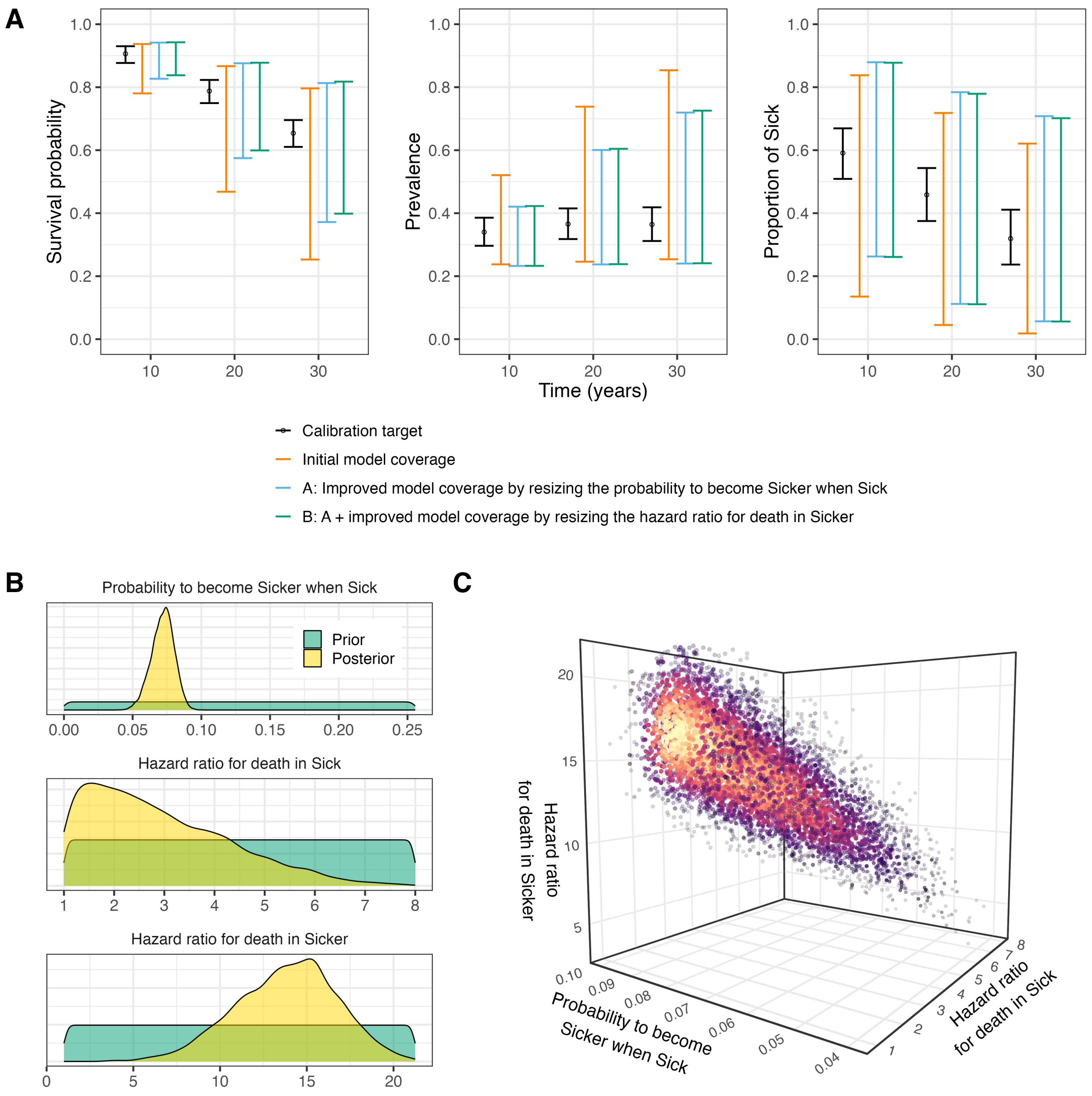


eFigure 3: More permissive widening of the prior bounds in the Sick-Sicker testbed model. (A) Coverage analysis after iteratively resizing and recentering the prior distributions using the local sensitivity results; the model intervals represent the intermediate stages of this iterative process. (B) Prior and posterior marginal distributions of calibrated parameters. (C) Joint distribution of calibrated parameters.

#### Appendix B

eFigure 4 shows the local and global sensitivity analyses of the SIR testbed model. Both methods consistently rank the transmission rate above the recovery rate as the dominant driver of model outputs during the early epidemic period. However, the global analysis reveals that the total Sobol' indices exceed the first-order indices for the transmission rate at earlier time points and for the recovery rate at later time points, indicating non-negligible parameter interactions that a local sensitivity analysis cannot capture.

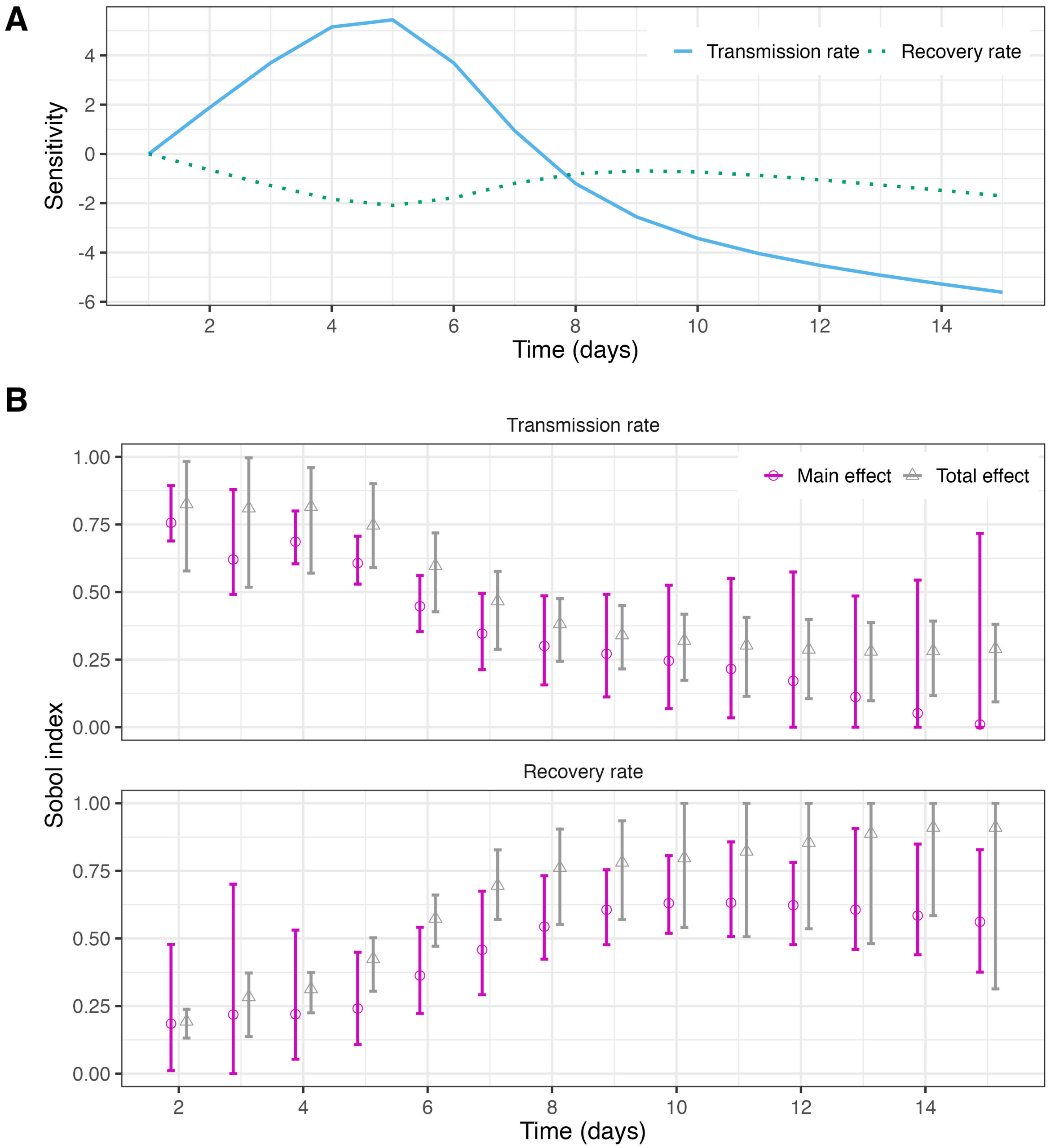


eFigure 4: Local and global sensitivity analysis of the SIR testbed model. (A) Local sensitivity analysis evaluated at the mean of the prior distribution. (B) Global sensitivity analysis displays first-order Sobol indices (main effects), which quantify each parameter's contribution to output variance, and total indices (total effects) that account for parameter interactions. The bootstrap confidence interval bounds were manually restricted to the plausible range $[0, 1]$.

Fit between SIR testbed model-predicted outputs and observed values following calibration.

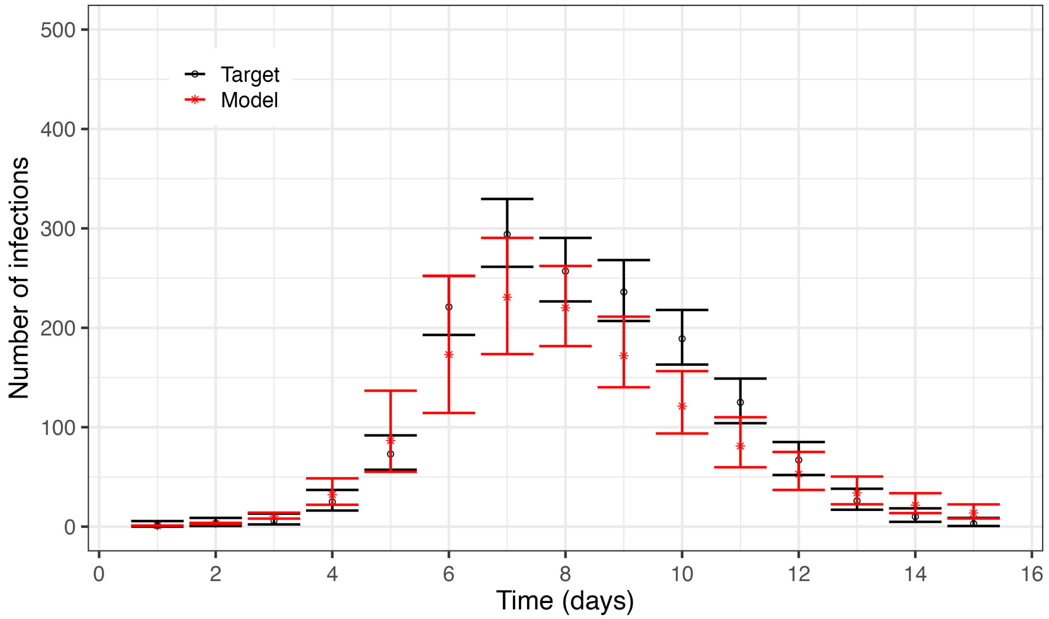


eFigure 5: Fit between model-predicted outputs and observed values following calibration in the SIR testbed model. Black dots and bars represent observed infections and their 95% confidence intervals, respectively. Red dots represent the mean posterior model-predicted values, and red bars indicate the model's 95% posterior predictive interval.

Implementation of PRE-CISE in the SIR testbed model using different starting points in the input parameter space.

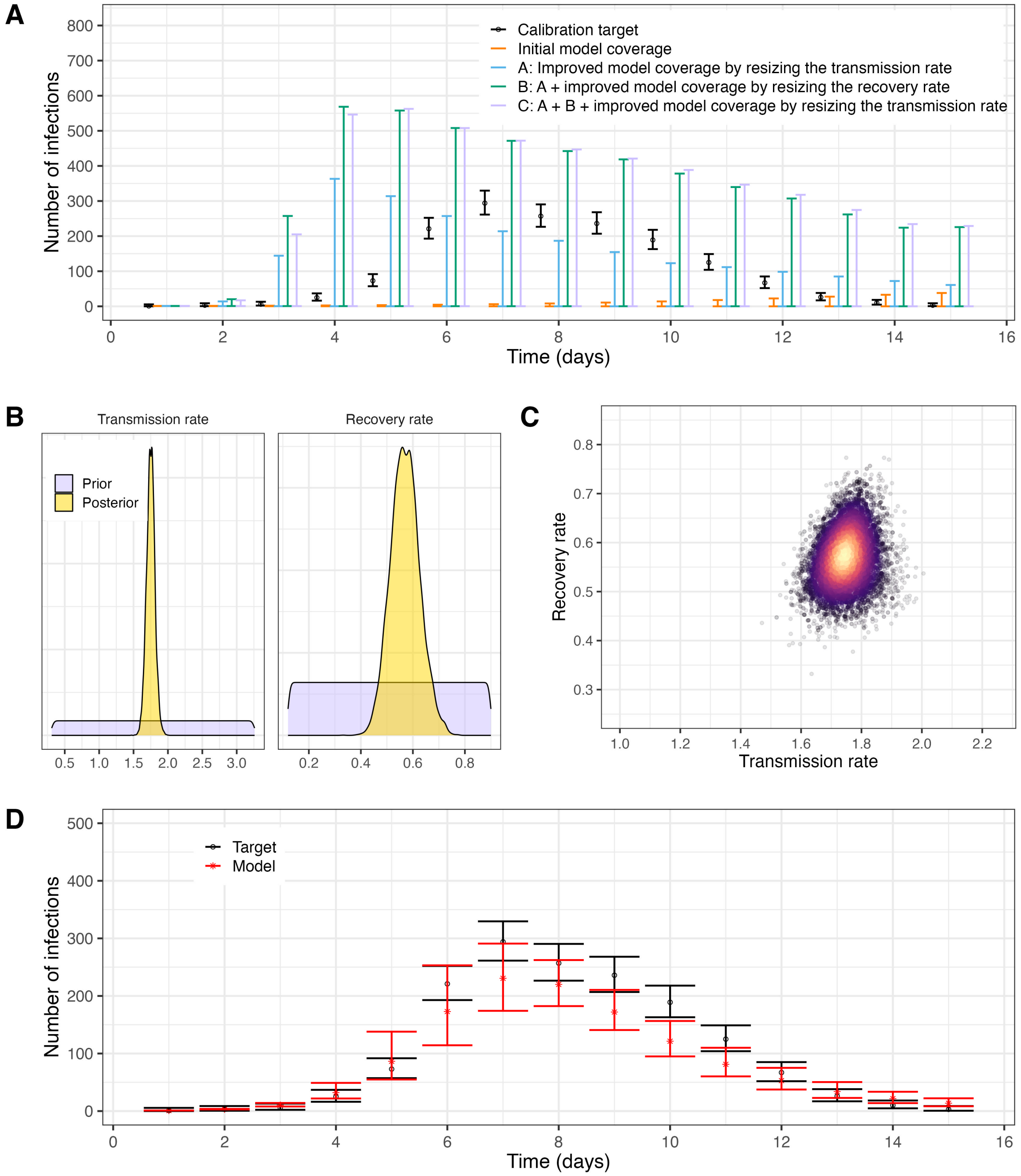


eFigure 6: PRE-CISE diagnostics for the SIR testbed model. (A) Coverage analysis after iteratively resizing and recentering the prior distributions using the local sensitivity results; the model intervals represent the intermediate stages of this iterative process. (B) Prior and posterior marginal distributions of calibrated parameters. (C) Joint distribution of calibrated parameters. (D) Fit between model-predicted outputs and observed values following calibration. Black dots and bars represent observed infections and their 95% confidence intervals, respectively. Red dots represent the mean posterior model-predicted values, and red bars indicate the model's 95% posterior predictive interval.

#### Appendix C

Collinearity analysis for the COVID-19 case study using daily incident cases as targets.

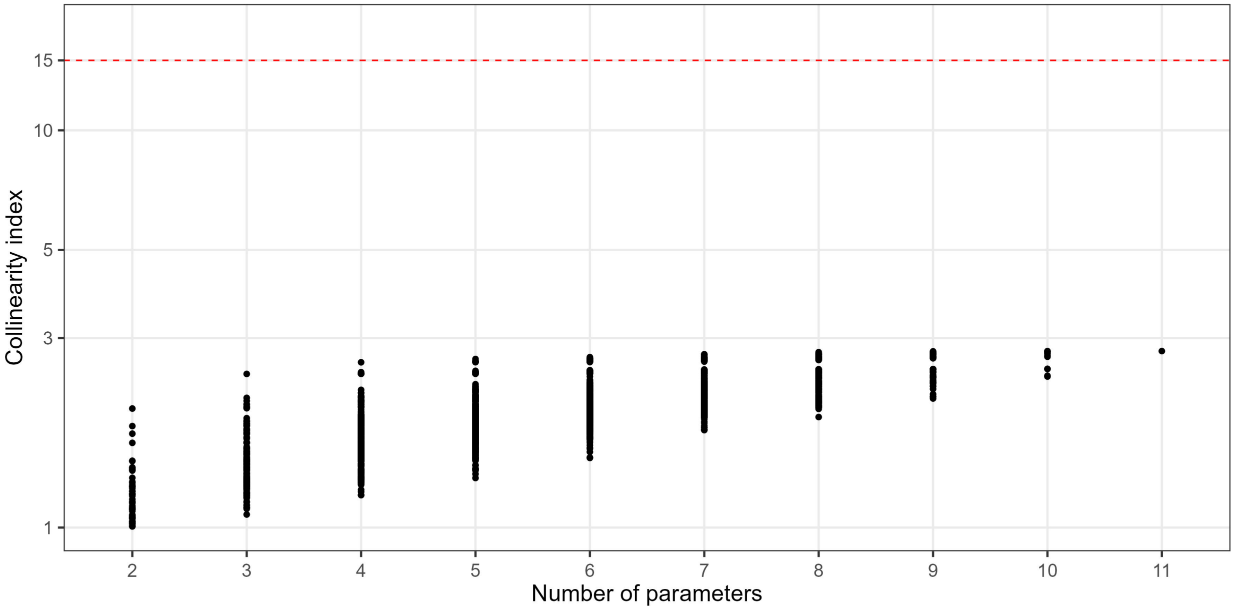


eFigure 7: Collinearity index for all parameter combinations using daily incident cases as targets. Black dots represent the values for different parameter combinations (e.g., for two-parameter sets, up to 55 parameter pairs are shown; overlapping points may appear as fewer). The red dashed line shows the highest value considered identifiable, which is 15 in this case.

Step-by-step process of the coverage analysis on the COVID-19 case study using daily incident cases as calibration targets.

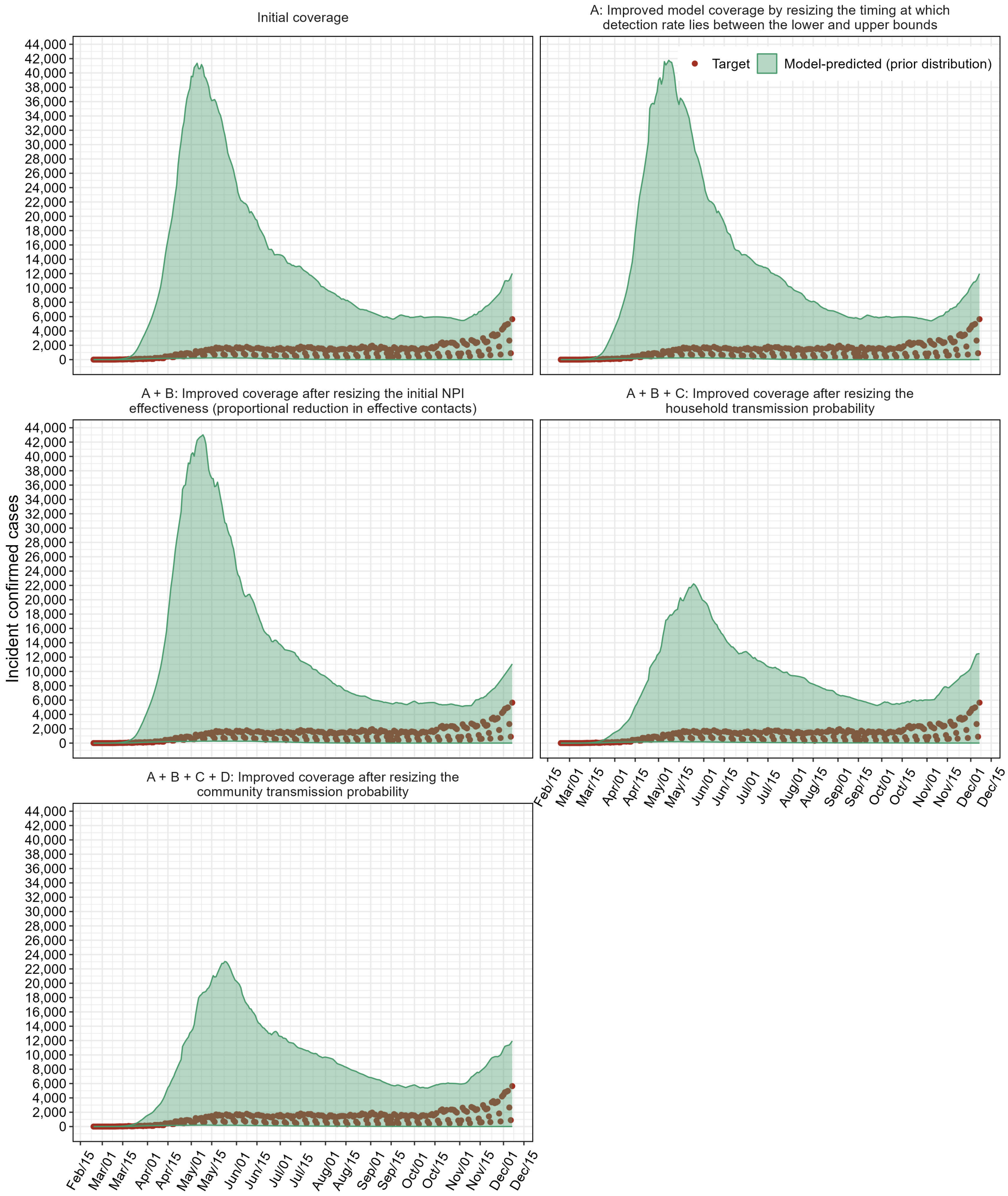


eFigure 8: Coverage analysis of model outputs across various input parameter spaces. Panels show the iterative process of resizing and recentering the prior distributions using the local sensitivity results. Shaded regions show the 95% prior predictive interval for daily incident cases; points show observed daily confirmed COVID-19 incident cases in MCMA from February 24, 2020, to December 7, 2020.

Implementation of PRE-CISE on the COVID-19 case study, which includes community and household transmission in the Mexico City Metropolitan Area, using weekly confirmed COVID-19 cases as calibration targets.

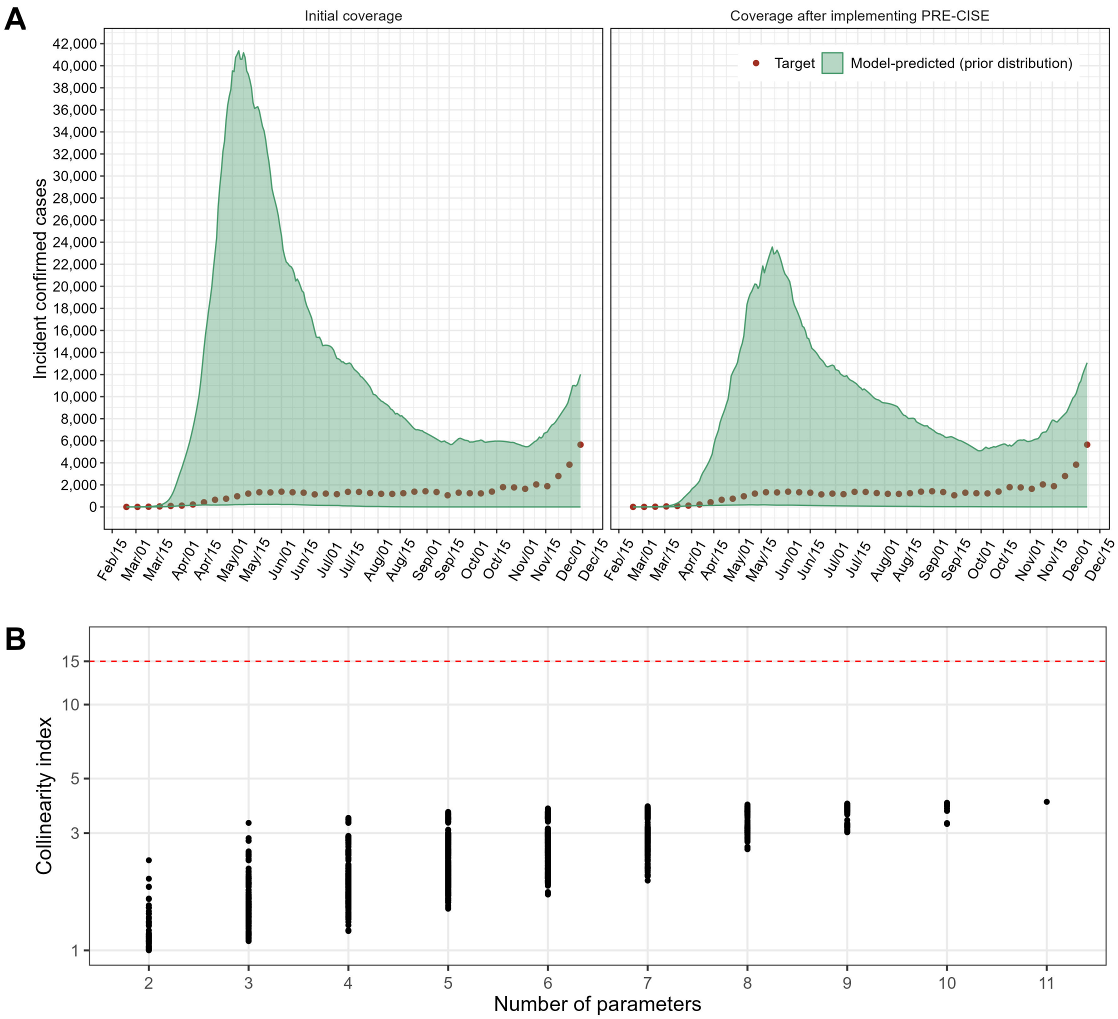


eFigure 9: (A) Coverage analysis using simulations drawn from the initial priors (left) and the resized priors after PRE-CISE (right). Shaded areas depict the 95% prior predicted interval for weekly incident cases; points show observed weekly confirmed COVID-19 incident cases in MCMA from February 24, 2020, to December 7, 2020. (B) Collinearity index for all parameter combinations using weekly incident cases as targets. Black dots represent the values for different parameter combinations (e.g., for two-parameter sets, up to 55 parameter pairs are shown; overlapping points may appear as fewer). The red dashed line shows the highest value considered identifiable, which is 15 in this case.

#### Appendix D

We evaluated PRE-CISE's performance by calibrating the testbeds and case study models with and without our proposed approach. For each model, we defined prior distributions for all parameters and performed an initial calibration without resizing the prior bounds. We then applied PRE-CISE to the same prior distributions, resized the bounds, and performed calibration. We recorded the number of model evaluations required at each stage: coverage analysis (baseline and resizing iterations) and calibration (baseline and incremental-sample-size iterations). For the Sick-Sicker and SIR testbed models, we assessed performance using two posterior sample sizes (1,000 and 10,000) to evaluate how PRE-CISE scales with sampling density. For the COVID-19 case study, we drew 1,000 samples from the posterior distribution.

Results are presented in eTables 1 and 2. Across scenarios, the net effect of PRE-CISE on total model evaluations ranged from a 2% increase to a 10% reduction. The gains were smallest, and in one case slightly negative, for low-dimensional testbeds calibrated to small posterior samples, where calibration converges in few IMIS iterations and the fixed cost of the coverage and sensitivity steps is not recovered. Gains were highest for denser posterior sampling and for the higher-dimensional COVID-19 case study. This pattern is expected: the value of pre-calibration diagnostics scales with the cost of the calibration they precede.

eTable 1: Performance of PRE-CISE in terms of model evaluations for the testbeds and the case study, and different posterior sample sizes.

| **Posterior sample size** | **PRE-CISE** | **Coverage analysis model evaluations** (baseline + resizing iterations) | **Calibration model evaluations** (baseline + incremental sample size) | **Total model evaluations** | |
|---|---|---|---|---|---|
| *Sick-Sicker Testbed* | | | | | |
| 1,000 | No | 0 | 15,000 | 15,000 | (-1%) |
| | Yes | 900 | 14,000 | 14,900 | |
| 10,000 | No | 0 | 30,000 | 30,000 | (-7%) |
| | Yes | 900 | 27,000 | 27,900 | |
| *SIR Testbed* | | | | | |
| 1,000 | No | 0 | 13,000 | 13,000 | (2%) |
| | Yes | 1,200 | 12,000 | 13,200 | |
| 10,000 | No | 0 | 23,000 | 23,000 | (-3%) |
| | Yes | 1,200 | 21,000 | 22,200 | |
| *COVID-19 Case Study* | | | | | |
| 1,000 | No | 0 | 46,000 | 46,000 | (-10%) |
| | Yes | 3,600 | 38,000 | 41,600 | |

*Note:* Relative change in the total number of model evaluations when PRE-CISE is applied is shown in parentheses; negative values indicate fewer evaluations.

eTable 2: Effective sample size (ESS) per 1,000 total model evaluations, with and without PRE-CISE

| **Model** | **Posterior sample size** | **Without PRE-CISE** | **With PRE-CISE** | **Relative change** |
|---|---|---|---|---|
| *Sick-Sicker Testbed* | 1,000 | 86.7 | 105.8 | 22% |
| | 10,000 | 321.9 | 339.8 | 6% |
| *SIR Testbed* | 1,000 | 144.7 | 98.8 | -32% |
| | 10,000 | 441.9 | 438.4 | -1% |
| *COVID-19 Case Study* | 1,000 | 13.0 | 15.7 | 21% |

*Note:* ESS is the IMIS effective sample size divided by the total number of model evaluations from eTable 1, including those consumed by PRE-CISE itself, and multiplied by 1,000. Positive relative changes indicate a more efficient posterior sample with PRE-CISE. Note that the sign convention differs from eTable 1, where negative values indicate fewer model evaluations.

### Appendix E

##### helper_functions.R

```r
#' Number of ticks for \code{ggplot2} plots
#'
#' Function for determining number of ticks on axis of \code{ggplot2} plots.
#' @param n Integer giving the desired number of ticks on axis of
#' \code{ggplot2} plots. Non-integer values are rounded down.
#' @section Details:
#' Based on function \code{pretty} (\code{dampack} package).
#' @export
number_ticks <- function(n) {
  function(limits) {
    pretty(limits, n + 1)
  }
}
```

##### SickSicker_functions.R

```r
# Model-decision functions -----------------------------------------------

#' Sick Sicker Model
#'
#' @param vparams Vector containing the model parameters.
#'
#' @return List containing the epidemiological outputs of the model
#' @export
sick_sicker_cstm <- function(v_params) {

  with(as.list(v_params), {

    ## Markov model parameters
    # age at baseline
    age     <- 25
    # maximum age of follow up
    max_age <- 55
    # time horizon, number of cycles
    n_t     <- max_age - age
    # the 4 states of the model: Healthy (H), Sick (S1), Sicker (S2), Dead (D)
    v_n     <- c("H", "S1", "S2", "D")
    # number of health states
    n_s     <- length(v_n)

    # Transition probabilities and hazard ratios
    # probability to die when healthy
    p_HD    <- 0.005
    # probability to become sick when healthy
    p_HS1   <- 0.15
    # probability to become healthy when sick
    p_S1H   <- 0.5
    # CALIBRATED: probability to become sicker when sick
    # p_S1S2  <- 0.105
    # CALIBRATED: hazard ratio of death in sick vs healthy
    # hr_S1   <- 3
    # CALIBRATED: hazard ratio of death in sicker vs healthy
    # hr_S2   <- 10

    # Compute internal parameters as a function of external parameters
    # rate of death in healthy
```

```
r_HD    <- - log(1 - p_HD)
# rate of death in sick
r_S1D   <- hr_S1 * r_HD
# rate of death in sicker
r_S2D   <- hr_S2 * r_HD
# probability to die in sick
p_S1D   <- 1 - exp(-r_S1D)
# probability to die in sicker
p_S2D   <- 1 - exp(-r_S2D)

### Initialization
# Create the cohort trace
# (n_t + 1 because R doesn't understand  Cycle 0)
m_M <- matrix(NA, nrow = n_t + 1 ,
              ncol = n_s,
              dimnames = list(0:n_t, v_n))
# Initialize Markov trace
m_M[1, ] <- c(1, 0, 0, 0)

# Create transition probability matrix for NO treatment
m_P <- matrix(0,
              nrow = n_s,
              ncol = n_s,
              dimnames = list(v_n, v_n))

# Fill in the transition probability array
# From Healthy
m_P["H", "H"]  <- 1 - (p_HS1 + p_HD)
m_P["H", "S1"] <- p_HS1
m_P["H", "D"]  <- p_HD

# From Sick
m_P["S1", "H"]  <- p_S1H
m_P["S1", "S1"] <- 1 - (p_S1H + p_S1S2 + p_S1D)
m_P["S1", "S2"] <- p_S1S2
m_P["S1", "D"]  <- p_S1D

# From Sicker
m_P["S2", "S2"] <- 1 - p_S2D
m_P["S2", "D"]  <- p_S2D

# From Dead
m_P["D", "D"] <- 1

# Check rows add up to 1
if (!isTRUE(all.equal(as.numeric(rowSums(m_P)), as.numeric(rep(1, n_s))))) {
  stop("This is not a valid transition Matrix")
}

### Process
for (t in 1:n_t){  # throughout the number of cycles
  # estimate the Markov trace for cycle the next cycle (t + 1)
  m_M[t + 1, ] <- m_M[t, ] %*% m_P
}

### Epidemiological outputs
# Overall Survival (OS)
# Calculate the overall survival (OS) probability for no treatment
v_os <- 1 - m_M[, "D"]

# Disease prevalence
v_prev <- rowSums(m_M[, c("S1", "S2")])/v_os
```

```r
    # Proportion of sick in S1 state
    v_prop_S1 <- m_M[, "S1"] / v_prev

    # Return output
    out <- data.frame(time     = 1:30,
                      PropSick = v_prop_S1[-1],
                      Prev     = v_prev[-1],
                      Surv     = v_os[-1])

    return(out)
  }
  )
}


# Visualization functions -------------------------------------------------

#' Plot model outputs vs targets
#'
#' \code{plot_model_out_vs_targets} plots model outputs vs targets.
#'
#' @param df_targets Data.frame containing the calibration targets.
#'
#' @return A ggplot2 object.
#'
#' @export
plot_targets <- function(df_targets){

  # Bind data.frames
  df_Surv <- df_targets %>% filter(abbrev_outcome == "Surv")
  df_Prev <- df_targets %>% filter(abbrev_outcome == "Prev")
  df_PropSick <- df_targets %>% filter(abbrev_outcome == "PropSick")

  # Plot Survival ("Surv")
  plotSurv <- ggplot(df_Surv,
                     aes(x = time, y = value, ymin = lb, ymax = ub)) +
    geom_point(shape = 1) + geom_errorbar() +
    scale_y_continuous(breaks = number_ticks(6)) +
    scale_x_continuous(breaks = number_ticks(6)) +
    ylab("Survival probability") + xlab("Time (years)") +
    theme_bw(base_size = 18) +
    theme(legend.position = "top",
          strip.background = element_rect(color = "white",
                                          fill = "white"),
          axis.text.x = element_text(angle = 0,
                                     vjust = 0.5,
                                     hjust = 0.5)) +
    coord_cartesian(ylim = c(0,1))

  # Plot Prevalence ("Prev")
  plotPrev <- ggplot(df_Prev,
                     aes(x = time, y = value, ymin = lb, ymax = ub)) +
    geom_point(size = 1, shape = 1) + geom_errorbar() +
    scale_y_continuous(breaks = number_ticks(6)) +
    scale_x_continuous(breaks = number_ticks(6)) +
    ylab("Prevalence") + xlab("Time (years)") +
    theme_bw(base_size = 18) +
    theme(legend.position = "top",
          strip.background = element_rect(color = "white",
                                          fill = "white"),
          axis.text.x = element_text(angle = 0,
                                     vjust = 0.5,
```

```r
                                        hjust = 0.5)) +
    coord_cartesian(ylim = c(0,1))

  # Plot Proportion who are Sick ("PropSick")
  plotPropSick <- ggplot(df_PropSick,
                         aes(x = time, y = value, ymin = lb, ymax = ub)) +
    geom_point(size = 1, shape = 1) + geom_errorbar() +
    scale_y_continuous(breaks = number_ticks(6)) +
    scale_x_continuous(breaks = number_ticks(6)) +
    ylab("Proportion of sick") + xlab("Time (years)") +
    theme_bw(base_size = 18) +
    theme(legend.position = "top",
          strip.background = element_rect(color = "white",
                                          fill = "white"),
          axis.text.x = element_text(angle = 0,
                                     vjust = 0.5,
                                     hjust = 0.5)) +
    coord_cartesian(ylim = c(0,1))

  # Gather plots
  gg_targets <- ggarrange(plotSurv, NULL,
                          plotPrev, NULL,
                          plotPropSick,
                          heights = 1,
                          widths = c(1,0.05,1,0.05,1),
                          ncol = 5, nrow = 1)

  return(gg_targets)

}

#' Plot model outputs vs targets
#'
#' \code{plot_model_out_vs_targets} plots model outputs vs targets.
#'
#' @param df_targets Data.frame containing the calibration targets.
#' @param df_model Data.frame containing the model outputs.
#'
#' @return A ggplot2 object.
#'
#' @export
plot_model_out_vs_targets <- function(df_targets, df_model){

  # Times to be plotted
  v_times <- sort(unique(df_targets$time))

  # Bind data.frames
  df_Surv <- rbind(
    df_targets %>% select(time, Outcome, abbrev_outcome, type, value, lb, ub),
    df_model %>% select(time, Outcome, abbrev_outcome, type, value, lb, ub)) %>%
    filter(abbrev_outcome == "Surv")

  df_Prev <- rbind(
    df_targets %>% select(time, Outcome, abbrev_outcome, type, value, lb, ub),
    df_model %>% select(time, Outcome, abbrev_outcome, type, value, lb, ub)) %>%
    filter(abbrev_outcome == "Prev")

  df_PropSick <- rbind(
    df_targets %>% select(time, Outcome, abbrev_outcome, type, value, lb, ub),
    df_model %>% select(time, Outcome, abbrev_outcome, type, value, lb, ub)) %>%
    filter(abbrev_outcome == "PropSick")
```

```r
# Order
df_Surv$type <- factor(df_Surv$type, levels = c("Target","Model"))
df_Prev$type <- factor(df_Prev$type, levels = c("Target","Model"))
df_PropSick$type <- factor(df_PropSick$type, levels = c("Target","Model"))

# Plot Survival ("Surv")
plotSurv <- ggplot(df_Surv,
                   aes(x = factor(time), y = value, ymin = lb, ymax = ub,
                       color = type, shape = type)) +
  geom_point(size = 1.5, position = position_dodge(0.8)) +
  geom_errorbar(position = position_dodge(0.8), linewidth = 0.8) +
  scale_shape_manual(NULL, values = c(Target = 1, Model = 8)) +
  scale_color_manual(NULL, values = c(Target = "black", Model = "red")) +
  scale_y_continuous(breaks = number_ticks(6)) +
  scale_x_discrete(breaks = as.character(v_times)) +
  ylab("Survival probability") + xlab("") +
  theme_bw(base_size = 18) +
  theme(legend.position = "none",
        strip.background = element_rect(color = "white",
                                        fill = "white"),
        axis.text.x = element_text(angle = 0,
                                   vjust = 0.5,
                                   hjust = 0.5)) +
  coord_cartesian(ylim = c(0,1))

# Plot Prevalence ("Prev")
plotPrev <- ggplot(df_Prev,
                   aes(x = factor(time), y = value, ymin = lb, ymax = ub,
                       color = type, shape = type)) +
  geom_point(size = 1.5, position = position_dodge(0.8)) +
  geom_errorbar(position = position_dodge(0.8), linewidth = 0.8) +
  scale_shape_manual(NULL, values = c(Target = 1, Model = 8)) +
  scale_color_manual(NULL, values = c(Target = "black", Model = "red")) +
  scale_y_continuous(breaks = number_ticks(6)) +
  scale_x_discrete(breaks = as.character(v_times)) +
  ylab("Prevalence") + xlab("Time (years)") +
  theme_bw(base_size = 18) +
  theme(legend.position = "none",
        strip.background = element_rect(color = "white",
                                        fill = "white"),
        axis.text.x = element_text(angle = 0,
                                   vjust = 0.5,
                                   hjust = 0.5)) +
  coord_cartesian(ylim = c(0,1))

# Plot Proportion who are Sick ("PropSick")
plotPropSick <- ggplot(df_PropSick,
                       aes(x = factor(time), y = value, ymin = lb, ymax = ub,
                           color = type, shape = type)) +
  geom_point(size = 1.5, position = position_dodge(0.8)) +
  geom_errorbar(position = position_dodge(0.8), linewidth = 0.8) +
  scale_shape_manual(NULL, values = c(Target = 1, Model = 8)) +
  scale_color_manual(NULL, values = c(Target = "black", Model = "red")) +
  scale_y_continuous(breaks = number_ticks(6)) +
  scale_x_discrete(breaks = as.character(v_times)) +
  ylab("Proportion of Sick") + xlab("") +
  theme_bw(base_size = 18) +
  theme(legend.position = "inside",
        legend.position.inside = c(0.65,0.90),
        strip.background = element_rect(color = "white",
                                        fill = "white"),
        axis.text.x = element_text(angle = 0,
```

```r
                                      vjust = 0.5,
                                      hjust = 0.5)) +
    coord_cartesian(ylim = c(0,1))

  # Gather plots
  gg_model_out_vs_targets <- ggarrange(plotSurv, NULL,
                                        plotPrev, NULL,
                                        plotPropSick,
                                        heights = 1,
                                        widths = c(1,0.05,1,0.05,1),
                                        ncol = 5, nrow = 1,
                                        common.legend = F)

  return(gg_model_out_vs_targets)

}

#' Plot coverage
#'
#' \code{plot_coverage} plots model outputs vs targets.
#'
#' @param df_targets Data.frame containing the calibration targets.
#' @param df_coverage Data.frame containing the coverage analysis.
#'
#' @return A ggplot2 object.
#'
#' @export
plot_coverage <- function(df_targets, df_coverage){

  # Times to be plotted
  v_times <- sort(unique(df_targets$time))

  # Add type_coverage to targets
  df_targets <- df_targets %>% mutate(type_coverage = "Calibration target")

  # Get levels
  v_levels <- c("Calibration target", unique(df_coverage$type_coverage))

  # Bind data.frames
  df_model_target <- rbind(df_targets, df_coverage)
  df_model_target$type_coverage <- factor(df_model_target$type_coverage,
                                           levels = v_levels)

  # Vector of shapes
  v_shape <- c(1,rep(NA,length(v_levels)-1))
  names(v_shape) <- v_levels

  # Vector of colors
  v_colors <- c("black","#FF8000","#56B4E9","#009E73","#CCBFFF","tan","firebrick")
  names(v_colors) <- c(
    "Calibration target",
    "Initial model coverage",
    "A: Improved model coverage by resizing the probability to become Sicker when Sick",
    "B: A + improved model coverage by resizing the hazard ratio for death in Sicker",
    "C: A + B + improved model coverage by resizing the hazard ratio for death in Sick",
    "D: A + B + C + improved model coverage by resizing the hazard ratio for death in Sick",
    "E: A + B + C + D + improved model coverage by resizing the hazard ratio for death in Sick"
  )

  # Plot Survival ("Surv")
  plotSurv <- ggplot(subset(df_model_target, abbrev_outcome == "Surv"),
                     aes(x = as.factor(time),
```

```
                         y = value, ymin = lb, ymax = ub,
                         color = type_coverage, shape = type_coverage)) +
  geom_point(size = 1.5, position = position_dodge(0.8)) +
  geom_errorbar(position = position_dodge(0.8), linewidth = 0.8) +
  scale_color_manual("", values = v_colors) +
  scale_shape_manual("", values = v_shape) +
  scale_y_continuous(breaks = number_ticks(6)) +
  scale_x_discrete(breaks = as.character(v_times)) +
  ylab("Survival probability") + xlab("") +
  guides(color = guide_legend(ncol=1, byrow=TRUE)) +
  theme_bw(base_size = 18) +
  theme(legend.position = "bottom",
        strip.background = element_rect(color = "white",
                                        fill = "white"),
        axis.text.x = element_text(angle = 0,
                                   vjust = 0.5,
                                   hjust = 0.5)) +
  coord_cartesian(ylim = c(0,1))

# Plot Prevalence ("Prev")
plotPrev <- ggplot(subset(df_model_target, abbrev_outcome == "Prev"),
                   aes(x = as.factor(time), y = value, ymin = lb, ymax = ub,
                       color = type_coverage, shape = type_coverage)) +
  geom_point(size = 1.5, position = position_dodge(0.8)) +
  geom_errorbar(position = position_dodge(0.8), linewidth = 0.8) +
  scale_color_manual("", values = v_colors) +
  scale_shape_manual("", values = v_shape) +
  scale_y_continuous(breaks = number_ticks(6)) +
  scale_x_discrete(breaks = as.character(v_times)) +
  ylab("Prevalence") + xlab("Time (years)") +
  guides(color = guide_legend(ncol=1, byrow=TRUE)) +
  theme_bw(base_size = 18) +
  theme(legend.position = "bottom",
        strip.background = element_rect(color = "white",
                                        fill = "white"),
        axis.text.x = element_text(angle = 0,
                                   vjust = 0.5,
                                   hjust = 0.5)) +
  coord_cartesian(ylim = c(0,1))

# Plot Proportion who are Sick ("PropSick")
plotPropSick <- ggplot(subset(df_model_target, abbrev_outcome == "PropSick"),
                       aes(x = as.factor(time), y = value, ymin = lb, ymax = ub,
                           color = type_coverage, shape = type_coverage)) +
  geom_point(size = 1.5, position = position_dodge(0.8)) +
  geom_errorbar(position = position_dodge(0.8), linewidth = 0.8) +
  scale_color_manual("", values = v_colors) +
  scale_shape_manual("", values = v_shape) +
  scale_y_continuous(breaks = number_ticks(6)) +
  scale_x_discrete(breaks = as.character(v_times)) +
  ylab("Proportion of Sick") + xlab("") +
  guides(color = guide_legend(ncol=1, byrow=TRUE)) +
  theme_bw(base_size = 18) +
  theme(legend.position = "bottom",
        strip.background = element_rect(color = "white",
                                        fill = "white"),
        axis.text.x = element_text(angle = 0,
                                   vjust = 0.5,
                                   hjust = 0.5)) +
  coord_cartesian(ylim = c(0,1))

# Gather plots
```

```r
  gg_model_out_vs_targets <- ggarrange(plotSurv, NULL,
                                       plotPrev, NULL,
                                       plotPropSick,
                                       heights = 1,
                                       widths = c(1,0.05,1,0.05,1),
                                       ncol = 5, nrow = 1,
                                       common.legend = T,
                                       legend = "bottom")

  return(gg_model_out_vs_targets)

}


#' Plot residuals of a model vs observations
#'
#' @param Cfun modCost object obtained from \code{costFun}.
#' @param wgt_res Flag (default is TRUE) of whether weighted
#' residuals should be plotted.
#'
#' @return A ggplot2 object.
#'
#' @export
plot_costFun <- function(Cfun, wgt_res = TRUE){

  # Times to be plotted
  v_times <- sort(unique(df_targets$time))

  # Get residuals
  df_residuals <- Cfun$residuals

  # Add labels to outputs
  df_residuals$name_lab <- ""
  df_residuals$name_lab[df_residuals$name == "Surv"] <- "Survival probability"
  df_residuals$name_lab[df_residuals$name == "Prev"] <- "Prevalence"
  df_residuals$name_lab[df_residuals$name == "PropSick"] <- "Proportion of Sick"

  df_residuals$name_lab <- factor(df_residuals$name_lab,
                                  levels = c("Survival probability",
                                             "Prevalence",
                                             "Proportion of Sick"))

  if(wgt_res){

    plotCfun <- ggplot(df_residuals,
                       aes(x = x, y = res, color = name_lab, shape = name_lab)) +
      geom_point() +
      facet_wrap(~ name_lab, scales = "free_y") +
      scale_y_continuous(breaks = number_ticks(6)) +
      scale_x_continuous(breaks = v_times) +
      ylab("Weighted residuals") + xlab("Time (years)") +
      theme_bw(base_size = 18) +
      theme(legend.position = "none",
            strip.background = element_rect(color = "white",
                                            fill = "white"),
            axis.text.x = element_text(angle = 0,
                                       vjust = 0.5,
                                       hjust = 0.5))

  }else{

    plotCfun <- ggplot(df_residuals,
```

```r
                        aes(x = x, y = res.unweighted, color = name_lab, shape = name_lab)) +
      geom_point() +
      facet_wrap(~ name_lab, scales = "free_y") +
      scale_y_continuous(breaks = number_ticks(6)) +
      scale_x_continuous(breaks = v_times) +
      ylab("Residuals") + xlab("Time (years)") +
      theme_bw(base_size = 18) +
      theme(legend.position = "none",
            strip.background = element_rect(color = "white",
                                            fill = "white"),
            axis.text.x = element_text(angle = 0,
                                       vjust = 0.5,
                                       hjust = 0.5))
  }

  return(plotCfun)
}

#' Plot sensitivity function
#'
#' \code{plot_Sfun} plots the sensitivity function.
#'
#' @param Sfun Sensitivity function obtained from \code{sensFun}.
#'
#' @return A ggplot2 object.
#'
#' @export
plot_Sfun <- function(Sfun){

  # Times to be plotted
  v_times <- sort(unique(df_targets$time))

  # Get variables and target names
  v_abb_targets <- unique(Sfun$var)
  v_param_names <- colnames(Sfun)[3:length(Sfun)]
  n_params <- length(v_param_names)

  # Generate long data.frame for plots
  df_Sfun_long <- data.frame(NULL)
  for(abb_targ_i in v_abb_targets){

    df_temp <- subset(Sfun, var == abb_targ_i) %>%
      pivot_longer(cols = all_of(v_param_names),
                   names_to = "parm",
                   values_to = "value")

    df_Sfun_long <- rbind(df_temp, df_Sfun_long)

  }
  df_plot <- df_Sfun_long %>%
    rename(Parameter = parm)

  # Categorical variables
  # Parameters
  df_plot$Parameter <- factor(df_plot$Parameter,
                              levels = v_param_names,
                              labels = c("Probability to become Sicker when Sick",
                                         "Hazard ratio for death in Sick",
                                         "Hazard ratio for death in Sicker"),
                              ordered = TRUE)

  # Outcomes
```

```
  df_plot$var_label <- ""
  df_plot$var_label[df_plot$var == "Surv"] <- "Survival probability"
  df_plot$var_label[df_plot$var == "Prev"] <- "Prevalence"
  df_plot$var_label[df_plot$var == "PropSick"] <- "Proportion of Sick"

  df_plot$var_label <- factor(df_plot$var_label,
                              levels = c("Survival probability",
                                         "Prevalence",
                                         "Proportion of Sick"))

  # Colors
  v_colors <- c("#56B4E9","#CCBFFF","#009E73")
  names(v_colors) <- c("Probability to become Sicker when Sick",
                       "Hazard ratio for death in Sick",
                       "Hazard ratio for death in Sicker")

  # Line type
  v_linetype <- c(1, 2, 3)
  names(v_linetype) <- c("Probability to become Sicker when Sick",
                         "Hazard ratio for death in Sick",
                         "Hazard ratio for death in Sicker")

  # Plot
  gg_Sfun <- ggplot(df_plot,
                    aes(x = x, y = value, color = Parameter,
                        linetype = Parameter)) +
    geom_line(linewidth = 1.2) + #geom_point(size = 1.5) +
    facet_wrap( ~ var_label, nrow = 1, scales = "free_y") +
    scale_color_manual("", values = v_colors) +
    scale_linetype_manual("", values = v_linetype) +
    scale_y_continuous(breaks = number_ticks(6)) +
    scale_x_continuous(breaks = v_times) +
    ylab("Sensitivity") + xlab("Time (years)") +
    theme_bw(base_size = 18) +
    theme(legend.position = "bottom",
          legend.direction = "horizontal",
          strip.background = element_rect(color = "white",
                                          fill = "white"),
          axis.text.x = element_text(angle = 0,
                                     vjust = 0.5,
                                     hjust = 0.5))

  return(gg_Sfun)
}

#' Plot collinearity index
#'
#' \code{plot_CollIndx} plots collinearity index
#'
#' @param df_collin Data.frame containing the collinearity indices
#' from \code{collin}.
#' @param log_y Flag (default is TRUE) to whether transform y-axis to log.

#' @return A ggplot2 object.
#'
#' @export
plot_CollIndx <- function(df_collin, log_y = TRUE){

  # Run coll
  collin_max <- max(df_collin$collinearity)

  # y-axis upper limit
```

```r
if(collin_max > 15){
  y_end <- collin_max + 3
}else{
  y_end <- 18
}

# x-axis discrete values
v_x_breaks <- unique(df_collin$N)

# Order outcomes
df_collin$Outcome <- factor(df_collin$Outcome,
                            levels = c("Survival probability",
                                       "Prevalence",
                                       "Proportion of Sick",
                                       "Survival probability & Prevalence",
                                       "Survival probability & Proportion of Sick",
                                       "Prevalence & Proportion of Sick",
                                       "Survival probability & Prevalence & Proportion of Sick"))

if(log_y){

  plot_collindx <- ggplot(subset(df_collin, var != "All"),
                          aes(x = factor(N), y = collinearity)) +
    geom_point() +
    theme_bw() +
    facet_wrap(~Outcome, ncol = 3) +
    scale_y_log10(n.breaks = 6) +
    scale_x_discrete(name = "Number of parameters",
                     breaks = paste0(v_x_breaks),
                     labels  = paste0(v_x_breaks)) +
    ylab("Collinearity index") +
    geom_hline(yintercept = 15,
               col = "red",
               linetype = "dashed") +
    theme_bw(base_size = 18) +
    theme(legend.position = "bottom",
          strip.background = element_rect(color = "white",
                                           fill = "white"),
          strip.text = element_text(size = 14),
          axis.text.x = element_text(angle = 0,
                                     vjust = 0.5,
                                     hjust = 0.5),
          panel.grid.minor = element_line(color = "white"))  +
    coord_cartesian(ylim = c(NA, y_end))

  plot_collindx_all <- ggplot(subset(df_collin, var == "All"),
                              aes(x = factor(N), y = collinearity)) +
    geom_point() +
    theme_bw() +
    facet_wrap(~Outcome, ncol = 1) +
    scale_y_log10(n.breaks = 6) +
    scale_x_discrete(name = "Number of parameters",
                     breaks = df_collin$N,
                     labels  = df_collin$N) +
    ylab("Collinearity index") +
    geom_hline(yintercept = 15,
               col = "red",
               linetype = "dashed") +
    theme_bw(base_size = 18) +
    theme(legend.position = "bottom",
          strip.background = element_rect(color = "white",
                                           fill = "white"),
```

```r
          strip.text = element_text(size = 14),
          axis.text.x = element_text(angle = 0,
                                     vjust = 0.5,
                                     hjust = 0.5),
          panel.grid.minor = element_line(color = "white"))  +
    coord_cartesian(ylim = c(NA, y_end))

}else{

  plot_collindx <- ggplot(subset(df_collin, var != "All"),
                          aes(x = factor(N), y = collinearity)) +
    geom_point() +
    theme_bw() +
    facet_wrap(~Outcome, ncol = 3) +
    scale_y_continuous(n.breaks = 7) +
    scale_x_discrete(name = "Number of parameters",
                     breaks = paste0(v_x_breaks),
                     labels  = paste0(v_x_breaks)) +
    ylab("Collinearity index") +
    xlab("Number of parameters") +
    geom_hline(yintercept = 15,
               col = "red",
               linetype = "dashed") +
    theme_bw(base_size = 18) +
    theme(legend.position = "bottom",
          strip.background = element_rect(color = "white",
                                          fill = "white"),
          strip.text = element_text(size = 14),
          axis.text.x = element_text(angle = 0,
                                     vjust = 0.5,
                                     hjust = 0.5),
          panel.grid.minor = element_line(color = "white"))  +
    coord_cartesian(ylim = c(NA, y_end))

  plot_collindx_all <- ggplot(subset(df_collin, var == "All"),
                              aes(x = factor(N), y = collinearity)) +
    geom_point() +
    theme_bw() +
    facet_wrap(~Outcome, ncol = 3) +
    scale_y_continuous(n.breaks = 7) +
    scale_x_discrete(name = "Number of parameters",
                     breaks = paste0(v_x_breaks),
                     labels  = paste0(v_x_breaks)) +
    ylab("Collinearity index") +
    xlab("Number of parameters") +
    geom_hline(yintercept = 15,
               col = "red",
               linetype = "dashed") +
    theme_bw(base_size = 18) +
    theme(legend.position = "bottom",
          strip.background = element_rect(color = "white",
                                          fill = "white"),
          strip.text = element_text(size = 14),
          axis.text.x = element_text(angle = 0,
                                     vjust = 0.5,
                                     hjust = 0.5),
          panel.grid.minor = element_line(color = "white"))  +
    coord_cartesian(ylim = c(NA, y_end))

}

# Gather plots
```

```
  gg_collindx <- ggarrange(plot_collindx, NULL,
                           plot_collindx_all,
                           heights = c(1,0.05,1),
                           widths = 1,
                           ncol = 1, nrow = 3)

  return(gg_collindx)
}

#' Plot global sensitivity analysis
#'
#' \code{plot_GSA} plots Sobol' first-order and total sensitivity indices.
#'
#' @param df_sobol Data.frame containing the results from a GSA using function
#' \code{run_GSA}.
#'
#' @returns A ggplot object.
#'
#' @export
plot_GSA <- function(df_sobol){

  # Outcomes
  v_outcomes <- unique(df_sobol$Outcome)
  n_outcomes <- length(v_outcomes)

  # Categorical variables
  # Parameters
  df_sobol$par <- factor(df_sobol$par,
                         levels = c("p_S1S2","hr_S1","hr_S2"),
                         labels = c("Probability to become Sicker when Sick",
                                    "Hazard ratio for death in Sick",
                                    "Hazard ratio for death in Sicker"),
                         ordered = TRUE)

  # Empty list to store plots
  l_GSA_plot <- vector(mode = "list", length = n_outcomes)
  i <- 1 # counter
  for(outcome_i in v_outcomes){ # outcome_i = "Prevalence"

    # Plot GSA by outcome
    l_GSA_plot[[i]] <- ggplot(subset(df_sobol, Outcome == outcome_i),
                              aes(x = as.factor(x), y = original, ymin = min..c.i., ymax = max..c.i.,
                                  color = type, shape = type)) +
      geom_point(position = position_dodge(width = 0.5), size = 3) +
      geom_errorbar(position = position_dodge(width = 0.5),
                    linewidth = 1.05,
                    width = 0.25) +
      scale_y_continuous(breaks = seq(0,1,0.25)) +
      ylab("Sobol index") + xlab("Time (years)") +
      scale_color_manual("", values = c("Main effect" = "#CD0BBC",
                                        "Total effect" = "#999999")) +
      scale_shape_manual("", values = c("Main effect" = 1,
                                        "Total effect" = 2)) +
      facet_wrap(~par, ncol = 1) +
      labs(subtitle = outcome_i) +
      theme_bw(base_size = 18) +
      theme(legend.position = "bottom",
            plot.subtitle = element_text(vjust = 0.5,
                                         hjust = 0.5),
            strip.text = element_text(size = 13),
            strip.background = element_rect(color = "white",
                                            fill = "white"),
```

```r
            axis.text.x = element_text(angle = 0,
                                       vjust = 0.5,
                                       hjust = 0.5)) +
      coord_cartesian(ylim = c(0,1))

    # Increase counter
    i <- i+1

  }

  # Return plot
  gg_GSA <- ggarrange(l_GSA_plot[[1]] + xlab(""), NULL,
                      l_GSA_plot[[2]] + ylab(""), NULL,
                      l_GSA_plot[[3]] + xlab("") + ylab(""),
                      ncol = 5, nrow = 1,
                      widths = c(1,0.05,1,0.05,1),
                      heights = c(1),
                      common.legend = T,
                      legend = "bottom",
                      label.y = "Sobol index",
                      label.x = "Time (years)")

  return(gg_GSA)
}

# Calibration functions ---------------------------------------------------

#' Generate model outputs for calibration from a parameter set
#'
#' \code{calibration_out} computes model outputs for the Sick-Sicker model
#' to be used for calibration routines.
#'
#' @param v_params Vector containing the calibrated parameters.
#'
#' @return A data.frame containing the model-predicted outcomes.
#'
#' @export
calibration_out <- function(v_params){

  # Run model
  df_model_out <- sick_sicker_cstm(v_params)

  # Data.frame for each calibration target
  # Survival
  df_Surv <- data.frame(time = df_model_out$time,
                        Outcome = "Survival",
                        abbrev_outcome = "Surv",
                        type = "Model",
                        value = df_model_out$Surv) %>%
    filter(time %in% v_times)

  # Prevalence
  df_Prev <- data.frame(time = df_model_out$time,
                        Outcome = "Prevalence",
                        abbrev_outcome = "Prev",
                        type = "Model",
                        value = df_model_out$Prev) %>%
    filter(time %in% v_times)

  # Proportion who are Sick ("PropSick")
  # among all those afflicted (Sick+Sicker)
  df_PropSick <- data.frame(time = df_model_out$time,
```

```r
                            Outcome = "Proportion of sick",
                            abbrev_outcome = "PropSick",
                            type = "Model",
                            value = df_model_out$PropSick) %>%
    filter(time %in% v_times)

  # Return data.frame
  return(list(df_Surv     = df_Surv,
              df_Prev     = df_Prev,
              df_PropSick = df_PropSick))
}

#' Sample from prior distributions of calibrated parameters
#'
#' \code{sample_prior} generates a sample of calibrated parameters
#' from their prior distribution.
#'
#' @param n_samp Number of samples.
#' @param v_lb Vector containing the lower bounds of the calibrated
#' parameters.
#' @param v_ub Vector containing the upper bounds of the calibrated
#' parameters.
#'
#' @return A matrix with n number of columns as number of parameters to
#' be calibrated and \code{n_samp} rows. Each row corresponds to a
#' parameter set sampled from their prior distributions.
#'
#' @export
sample_prior <- function(n_samp, v_lb, v_ub){

  m_lhs_unit   <- randomLHS(n = n_samp, k = n_param)
  m_param_samp <- matrix(nrow = n_samp, ncol = n_param)
  colnames(m_param_samp) <- v_param_names
  for (i in 1:n_param){
    m_param_samp[, i] <- qunif(m_lhs_unit[,i],
                               min = v_lb[i],
                               max = v_ub[i])
    # ALTERNATIVE prior using beta (or other) distributions
    # m_param_samp[, i] <- qbeta(m_lhs_unit[,i],
    #                            shape1 = 1,
    #                            shape2 = 1)
  }
  return(m_param_samp)
}

#' Evaluate log-prior of calibrated parameters
#'
#' \code{log_prior} computes a log-prior value for one (or multiple) parameter
#' set(s) based on their prior distributions.
#'
#' @param v_params Vector (or matrix) containing the model parameters.
#' @param v_lb Vector containing the lower bounds for each parameter.
#' @param v_ub Vector containing the upper bounds for each parameter.
#'
#' @return A scalar (or vector) containing the log-prior values.
#'
#' @export
log_prior <- function(v_params, v_lb, v_ub){

  # Get param names
  v_param_names <- names(v_lb)
```

```r
  if(is.null(dim(v_params))) { # If vector, change to matrix
    v_params <- t(v_params)
  }
  n_samp <- nrow(v_params)
  colnames(v_params) <- v_param_names
  lprior <- rep(0, n_samp)
  for (i in 1:n_param){
    lprior <- lprior + dunif(v_params[, i],
                             min = v_lb[i],
                             max = v_ub[i],
                             log = T)
    # ALTERNATIVE prior using beta distributions
    # lprior <- lprior + dbeta(v_params[, i],
    #                          shape1 = 1,
    #                          shape2 = 1,
    #                          log = T)
  }
  return(lprior)
}

#' Evaluate prior of calibrated parameters
#'
#' \code{prior} computes a prior value for one (or multiple) parameter set(s).
#'
#' @param v_params Vector (or matrix) containing the model parameters.
#' @param v_lb Vector containing the with lower bounds for each parameter.
#' @param v_ub Vector containing the with upper bounds for each parameter.
#'
#' @return A scalar (or vector) containing the prior values.
#'
#' @export
prior <- function(v_params, v_lb, v_ub) {
  return(exp(log_prior(v_params, v_lb, v_ub)))
}

#' Log likelihood normal distribution
#'
#' \code{log_lik} computes a log-likelihood value for one (or multiple)
#' parameter set(s).
#'
#' @param v_params_calib Vector (or matrix) containing the calibrated parameters.
#'
#' @return A scalar (or vector) containing the log-likelihood values.
#'
#' @export
log_lik <- function(v_params){

  # par_vector: a vector (or matrix) of model parameters
  if(is.null(dim(v_params))) { # If vector, change to matrix
    v_params <- t(v_params)
  }
  n_samp <- nrow(v_params)
  v_abbrev_outcomes <- c("Surv", "Prev", "PropSick")
  n_outcomes <- length(v_abbrev_outcomes)
  v_llik <- matrix(0, nrow = n_samp, ncol = n_outcomes)
  llik_overall <- numeric(n_samp)
  for(j in 1:n_samp) { # j=1
    jj <- tryCatch( {
      ###   Run model for parameter set "v_params" ###
      l_model_out <- calibration_out(v_params[j, ])

      ### Targets
```

```r
      df_targets_Prev <- df_targets %>%
        filter(abbrev_outcome == "Prev")
      df_targets_PropSick <- df_targets %>%
        filter(abbrev_outcome == "PropSick")
      df_targets_Surv <- df_targets %>%
        filter(abbrev_outcome == "Surv")

      ###  Calculate log-likelihood of model outputs to targets  ###
      # TARGET 1: Survival ("Surv")
      # log likelihood
      v_llik[j, 1] <- sum(dnorm(x = df_targets_Surv$value,
                                mean = l_model_out$df_Surv$value,
                                sd = df_targets_Surv$se,
                                log = T))

      # TARGET 2: Prevalence ("Prev")
      # log likelihood
      v_llik[j, 2] <- sum(dnorm(x = df_targets_Prev$value,
                                mean = l_model_out$df_Prev$value,
                                sd = df_targets_Prev$se,
                                log = T))

      # TARGET 3: Proportion of sick in Sick state ("PropSick")
      # log likelihood
      v_llik[j, 3] <- sum(dnorm(x = df_targets_PropSick$value,
                                mean = l_model_out$df_PropSick$value,
                                sd = df_targets_PropSick$se,
                                log = T))

      # OVERALL
      llik_overall[j] <- sum(v_llik[j, ])
    }, error = function(e) NA)
    if (is.na(jj)) { llik_overall <- -Inf }
  } # End loop over sampled parameter sets
  # return LLIK
  return(llik_overall)
}

#' Likelihood
#'
#' \code{likelihood} computes a likelihood value for one (or multiple)
#' parameter set(s).
#'
#' @param v_params_calib Vector (or matrix) containing the model parameters.
#'
#' @return A scalar (or vector) containing the likelihood values.
#'
#' @export
likelihood <- function(v_params){
  return(exp(log_lik(v_params)))
}

#' Evaluate log-posterior of calibrated parameters
#'
#' \code{log_post} computes a log-posterior value for one (or multiple)
#' parameter set(s) based on the simulation model, likelihood functions and
#' prior distributions.
#'
#' @param v_params_calib Vector (or matrix) containing the model parameters.
#' @param v_lb Vector containing the lower bounds for each parameter.
#' @param v_ub Vector containing the upper bounds for each parameter.
#'
```

```r
#' @return A scalar (or vector) containing the log-posterior values.
#'
#' @export
log_post <- function(v_params, v_lb, v_ub) {
  lpost <- log_prior(v_params, v_lb, v_ub) + log_lik(v_params)
  return(lpost)
}

#' Evaluate posterior of calibrated parameters
#'
#' \code{posterior} computes a posterior value for one (or multiple)
#' parameter set(s).
#'
#' @param v_params_calib Vector (or matrix) containing the model parameters.
#' @param v_lb Vector containing the lower bounds for each parameter.
#' @param v_ub Vector containing the upper bounds for each parameter.
#'
#' @return A scalar (or vector) containing the posterior values.
#'
#' @export
posterior <- function(v_params, v_lb, v_ub) {
  exp(log_post(v_params, v_lb, v_ub))
}
```

**SickSicker_IMIS_function.R**

```r
#' Incremental Mixture Importance Sampling (IMIS package) adapted for the
#' Sick-Sicker model
#'
#' @param B The incremental sample size at each iteration of IMIS.
#' @param B.re The desired posterior sample size at the resample stage.
#' @param number_k The maximum number of iterations in IMIS
#' @param D The number of optimizers which could be 0.
#' @param v_lb Vector containing the lower bounds of the calibrated
#' parameters.
#' @param v_ub Vector containing the upper bounds of the calibrated
#' parameters.
#'
#' @return The posterior resamples.
#' @source Adrain Raftery and Le Bao. IMIS package.
#' http://cran.nexr.com/web/packages/IMIS/index.html
#'
#' @import mvtnorm
#' @export
IMIS <- function(B=1000, B.re=3000, number_k=100, D=0, v_lb, v_ub){
  B0 = B*10
  # Draw initial samples from the prior distribution
  X_all = X_k = sample_prior(B0, v_lb = v_lb, v_ub = v_ub)
  if (is.vector(X_all))        Sig2_global = var(X_all)        # the prior covariance
  if (is.matrix(X_all))        Sig2_global = cov(X_all)        # the prior covariance

  # 6 diagnostic statistics at each iteration
  stat_all = matrix(NA, 6, number_k)

  # centers of Gaussian components, prior densities, and likelihoods
  center_all = prior_all = like_all = NULL

  # covariance matrices of Gaussian components
  sigma_all = list()
  if (D>=1)        option.opt = 1             # use optimizer
```

```
if (D==0) option.opt = 0; D=1        # NOT use optimizer

for (k in 1:number_k){ # k = 1

  ptm.like = proc.time()
  prior_all = c(prior_all,
                prior(X_k, v_lb = v_lb, v_ub = v_ub))  # Calculate the prior densities
  like_all = c(like_all, likelihood(X_k))                # Calculate the likelihoods
  ptm.use = (proc.time() - ptm.like)[3]
  if (k==1){
    print(paste(B0, "likelihoods are evaluated in",
                round(ptm.use/60,2), "minutes"))
  }

  if (k==1)        envelop_all = prior_all        # envelop stores the sampling densities
  if (k>1){
    envelop_all = apply(rbind(prior_all*B0/B, gaussian_all), 2, sum)/(B0/B+D+(k-2))
  }

  # importance weight is determined by the posterior density divided
  # by the sampling density
  Weights = prior_all*like_all / envelop_all
  stat_all[1,k] = log(mean(Weights))                      # the raw marginal likelihood
  Weights = Weights / sum(Weights)
  stat_all[2,k] = sum(1-(1-Weights)^B.re)        # the expected number of unique points
  stat_all[3,k] = max(Weights)                                  # the maximum weight
  stat_all[4,k] = 1/sum(Weights^2)                        # the effective sample size
  # the entropy relative to uniform
  stat_all[5,k] = -sum(Weights*log(Weights), na.rm = TRUE) / log(length(Weights))
  stat_all[6,k] = var(Weights/mean(Weights))        # the variance of scaled weights
  if (k==1)        print("Stage   MargLike   UniquePoint   MaxWeight   ESS")
  print(c(k, round(stat_all[1:4,k], 3)))

  if (k==1 & option.opt==1){
    if (is.matrix(X_all))        Sig2_global = cov(X_all[which(like_all>min(like_all)),])

    # exclude the neighborhood of the local optima
    X_k = which_exclude = NULL

    label_weight = sort(Weights, decreasing = TRUE, index=TRUE)

    # the candidate inputs for the starting points
    which_remain = which(Weights>label_weight$x[B0])
    size_remain = length(which_remain)
    for (i in 1:D){
      important = NULL
      if (length(which_remain)>0)
        important = which_remain[which(Weights[which_remain]==max(Weights[which_remain]))]
      if (length(important)>1)        important = sample(important,1)
      if (is.vector(X_all))        X_imp = X_all[important]
      if (is.matrix(X_all))        X_imp = X_all[important,]
      # Remove the selected input from candidates
      which_exclude = union( which_exclude, important )
      which_remain = setdiff(which_remain, which_exclude)
      posterior = function(theta){        -log(prior(theta, v_lb=v_lb, v_ub=v_ub))-
          log(likelihood(theta)) }

      if (is.vector(X_all)){
        if (length(important)==0)        X_imp = center_all[1]
        optimizer = optim(X_imp, posterior, method="BFGS", hessian=TRUE,
                          control=list(parscale=sqrt(Sig2_global)/10,maxit=5000))
        print(paste("maximum posterior=", round(-optimizer$value,2),
```

```
                ", likelihood=", round(log(likelihood(optimizer$par)),2),
                ", prior=",
                round(log(prior(optimizer$par,
                                v_lb = v_lb,
                                v_ub = v_ub)),2),
                ", time used=", round(ptm.use/60,2),
                "minutes, convergence=", optimizer$convergence))
    center_all = c(center_all, optimizer$par)
    sigma_all[[i]] = solve(optimizer$hessian)
    # Draw new samples:
    X_k = c(X_k, rnorm(B, optimizer$par, sqrt(sigma_all[[i]])) )
    distance_remain = abs(X_all[which_remain]-optimizer$par)
  }
  if (is.matrix(X_all)){
    # The rough optimizer uses the Nelder-Mead algorithm.
    if (length(important)==0)        X_imp = center_all[1,]
    ptm.opt = proc.time()
    optimizer = optim(X_imp, posterior, method="Nelder-Mead",
                      control=list(maxit=1000, parscale=sqrt(diag(Sig2_global))) )
    theta.NM = optimizer$par

    # The more efficient optimizer uses the BFGS algorithm
    optimizer = optim(theta.NM, posterior, method="BFGS", hessian=TRUE,
                      control=list(parscale=sqrt(diag(Sig2_global)), maxit=1000))
    ptm.use = (proc.time() - ptm.opt)[3]
    print(paste("maximum posterior=", round(-optimizer$value,2),
                ", likelihood=", round(log(likelihood(optimizer$par)),2),
                ", prior=",
                round(log(prior(optimizer$par,
                                v_lb = v_lb,
                                v_ub = v_ub)),2),
                ", time used=", round(ptm.use/60,2),
                "minutes, convergence=", optimizer$convergence))
    center_all = rbind(center_all, optimizer$par)               # the center of new samples
    if (min(eigen(optimizer$hessian)$values)>0){
      # the covariance of new samples
      sigma_all[[i]] = solve(optimizer$hessian)
    }

    # If the hessian matrix is not positive definite, we define the covariance as following
    if (min(eigen(optimizer$hessian)$values)<=0){
      eigen.values = eigen(optimizer$hessian)$values
      eigen.values[which(eigen.values<0)] = 0
      hessian = eigen(optimizer$hessian)$vectors %*% diag(eigen.values) %*%
        t(eigen(optimizer$hessian)$vectors)
      sigma_all[[i]] = solve(hessian + diag(1/diag(Sig2_global)) )
    }
    # Draw new samples
    X_k = rbind(X_k, rmvnorm(B, optimizer$par, sigma_all[[i]]) )
    distance_remain = mahalanobis(X_all[which_remain,],
                                  optimizer$par,
                                  diag(diag(Sig2_global)) )
  }
  # exclude the neighborhood of the local optima
  label_dist = sort(distance_remain, decreasing = FALSE, index=TRUE)
  which_exclude = union(which_exclude,
                        which_remain[label_dist$ix[1:floor(size_remain/D)]])
  which_remain = setdiff(which_remain, which_exclude)
}
if (is.matrix(X_all))        X_all = rbind(X_all, X_k)
if (is.vector(X_all))        X_all = c(X_all, X_k)
```

```
    saveRDS(Sig2_global, file = paste0(path,"Sig2_global.rds"))
  }

  if (k>1 | option.opt==0){
    important = which(Weights == max(Weights))
    if (length(important)>1)        important = important[1]

    # X_imp is the maximum weight input
    if (is.matrix(X_all))        X_imp = X_all[important,]

    if (is.vector(X_all))        X_imp = X_all[important]
    if (is.matrix(X_all))        center_all = rbind(center_all, X_imp)
    if (is.vector(X_all))        center_all = c(center_all, X_imp)
    if (is.matrix(X_all)){
      distance_all = mahalanobis(X_all, X_imp, diag(diag(Sig2_global)))
    }

    # Calculate the distances to X_imp
    if (is.vector(X_all))        distance_all = abs(X_all-X_imp)
    # Sort the distances
    label_nr = sort(distance_all, decreasing = FALSE, index=TRUE)
    which_var = label_nr$ix[1:B]        # Pick B inputs for covariance calculation
    if (is.matrix(X_all)){
      Sig2 = cov.wt(X_all[which_var,],
                    wt = Weights[which_var]+1/length(Weights),
                    cor = FALSE, center = X_imp, method = "unbias")$cov
    }
    if (is.vector(X_all)){
      Weights_var = Weights[which_var]+1/length(X_all)
      Weights_var = Weights_var/sum(Weights_var)
      Sig2 = (X_all[which_var]-X_imp)^2 %*% Weights_var
    }
    sigma_all[[D+k-1]] = Sig2
    if (is.matrix(X_all)){
      X_k = rmvnorm(B, X_imp, Sig2) # Draw new samples
    }
    if (is.vector(X_all)){
      X_k = rnorm(B, X_imp, sqrt(Sig2)) # Draw new samples
      }
    if (is.matrix(X_all))        X_all = rbind(X_all, X_k)
    if (is.vector(X_all))        X_all = c(X_all, X_k)
  }

  if (k==1){
    gaussian_all = matrix(NA, D, B0+D*B)
    for (i in 1:D){
      if (is.matrix(X_all)){
        gaussian_all[i,] = dmvnorm(X_all, center_all[i,], sigma_all[[i]])
      }
      if (is.vector(X_all)){
        gaussian_all[i,] = dnorm(X_all, center_all[i], sqrt(sigma_all[[i]]))
      }
    }
  }
  if (k>1){
    if (is.vector(X_all))        gaussian_new = matrix(0, D+k-1, length(X_all) )
    if (is.matrix(X_all))        gaussian_new = matrix(0, D+k-1, dim(X_all)[1] )
    if (is.matrix(X_all)){
      gaussian_new[1:(D+k-2), 1:(dim(X_all)[1]-B)] = gaussian_all
      gaussian_new[D+k-1, ] = dmvnorm(X_all, X_imp, sigma_all[[D+k-1]])
      for (j in 1:(D+k-2)){
        gaussian_new[j, (dim(X_all)[1]-B+1):dim(X_all)[1] ] =
```

```
          dmvnorm(X_k, center_all[j,], sigma_all[[j]])
        }
      }
      if (is.vector(X_all)){
        gaussian_new[1:(D+k-2), 1:(length(X_all)-B)] = gaussian_all
        gaussian_new[D+k-1, ] = dnorm(X_all, X_imp, sqrt(sigma_all[[D+k-1]]))
        for (j in 1:(D+k-2)){
          gaussian_new[j, (length(X_all)-B+1):length(X_all) ] =
            dnorm(X_k, center_all[j], sqrt(sigma_all[[j]]))
        }
      }
      gaussian_all = gaussian_new
    }
    if (stat_all[2,k] > (1-exp(-1))*B.re)        break
  } # end of k

  nonzero = which(Weights>0)
  which_X = sample(nonzero, B.re, replace = TRUE, prob = Weights[nonzero])
  if (is.matrix(X_all))        resample_X = X_all[which_X,]
  if (is.vector(X_all))        resample_X = X_all[which_X]

  return(list(stat=t(stat_all), resample=resample_X, center=center_all))
} # end of IMIS
```

**SickSicker_analysis.R**

```
#-----------------------------------------------------------------------------#
# This script illustrates PRE-CISE and its implementation in a four-state      #
# Sick-Sicker cohort Markov model calibrated to survival probability,          #
# prevalence, and proportion of Sick using the IMIS algorithm.                 #
#-----------------------------------------------------------------------------#
rm(list = ls()) # clean environment

# Calibration Specifications ----------------------------------------------
# Model: 4-state Sick-Sicker Markov Model
# Inputs to be calibrated:
#   p_S1S2 - probability to become sicker when sick
#   hr_S1  - hazard ratio of death in sick vs healthy
#   hr_S2  - hazard ratio of death in sicker vs healthy
# Targets:
#   Surv     - survival probability
#   Prev     - prevalence (sick and sicker)
#   PropSick - proportion of Sick
# Search method: Random search using Latin-Hypercube Sampling
# Goodness-of-fit measure: Sum of Log-Likelihood

# Load libraries and functions --------------------------------------------
# calibration functionality
library(lhs)
library(matrixStats) # package used for summary statistics
library(FME)         # For identifiability and collinearity analysis
library(tidyverse)
options(dplyr.summarise.inform = FALSE) # do not show summarise info
library(mvtnorm)

# visualization
library(plotrix)
library(psych)
library(ggplot2)
library(ggthemes)
```

```r
library(ggpubr)
library(scatterplot3d) # three inputs to estimate
library(ggcube)

# global sensitivity analysis
library(sensitivity)

# calibration
source("R/IMIS_function_SickSicker.R") # from IMIS package, adapted for this analysis
# Old version
# devtools::install_version("IMIS", version = "0.1", repos = "http://cran.us.r-project.org")

# functions
source("R/helper_functions.R") # from dampack package
source("R/SickSicker_functions.R")

# Variables ------------------------------------------------------------
# Number of simulations (coverage analysis)
n_sim <- 300

# Seed number
n_seed <- 111124

# Set seed
set.seed(n_seed)

# Vector of times (in years) at which the model will be calibrated
v_times <- c(10, 20, 30)

# Target data ----------------------------------------------------------
load("data/SickSicker_CalibTargets.RData")
l_targets <- SickSicker_targets

# Gather targets in a data.frame
df_targets <- rbind(l_targets$Surv %>%
                      mutate(type = "Target",
                             Outcome = "Survival",
                             abbrev_outcome = "Surv") %>%
                      select(time, Outcome, abbrev_outcome, type, value, lb, ub, se),
                    l_targets$Prev %>%
                      mutate(type = "Target",
                             Outcome = "Prevalence",
                             abbrev_outcome = "Prev") %>%
                      select(time, Outcome, abbrev_outcome, type, value, lb, ub, se),
                    l_targets$PropSick %>%
                      mutate(type = "Target",
                             Outcome = "Proportion of Sick",
                             abbrev_outcome = "PropSick") %>%
                      select(time, Outcome, abbrev_outcome, type, value, lb, ub, se)) %>%
  filter(time %in% v_times) # filter by the years to be calibrated

# Plot the targets
plot_targets(df_targets)

# Model ----------------------------------------------------------------
# -inputs are parameters to be estimated through calibration
# -outputs correspond to the target data

# Check that it works
v_params_test <- c(p_S1S2 = 0.105, hr_S1 = 3, hr_S2 = 10)
sick_sicker_cstm(v_params_test) # It works!
```

```
# Specify calibration parameters ------------------------------------------
# Names and number of input parameters to be calibrated
v_param_names <- c("p_S1S2", # probability to become sicker when sick
                   "hr_S1",  # hazard ratio of death in sick vs healthy
                   "hr_S2")  # hazard ratio of death in sicker vs healthy
n_param <- length(v_param_names)

# Range on input search space
v_lb <- c(p_S1S2 = 0.01, hr_S1 = 1.0, hr_S2 = 5.0)  # lower bound
v_ub <- c(p_S1S2 = 0.50, hr_S1 = 4.5, hr_S2 = 15.0) # upper bound

# Number of calibration targets
v_outcome_names <- c("Survival probability", "Prevalence", "Proportion of Sick")
names(v_outcome_names) <- c("Surv", "Prev", "PropSick")
n_outcomes <- length(v_outcome_names)

# Calibration functions (check that it works) -----------------------------
# View resulting parameter set samples
pairs.panels(sample_prior(n_samp = 1000, v_lb = v_lb, v_ub = v_ub))

# Prior
log_prior(v_params = v_params_test,
          v_lb     = v_lb,
          v_ub     = v_ub)
log_prior(v_params = sample_prior(10, v_lb = v_lb, v_ub = v_ub),
          v_lb     = v_lb,
          v_ub     = v_ub)

prior(v_params = v_params_test,
      v_lb     = v_lb,
      v_ub     = v_ub)
prior(v_params = sample_prior(10, v_lb = v_lb, v_ub = v_ub),
      v_lb     = v_lb,
      v_ub     = v_ub)

# Likelihood
log_lik(v_params = v_params_test)
log_lik(v_params = sample_prior(10, v_lb = v_lb, v_ub = v_ub))

likelihood(v_params = v_params_test)
likelihood(v_params = sample_prior(10, v_lb = v_lb, v_ub = v_ub))

# Posterior
log_post(v_params = v_params_test,
         v_lb     = v_lb,
         v_ub     = v_ub)
log_post(v_params = sample_prior(10, v_lb = v_lb, v_ub = v_ub),
         v_lb     = v_lb,
         v_ub     = v_ub)

posterior(v_params = v_params_test,
          v_lb     = v_lb,
          v_ub     = v_ub)
posterior(v_params = sample_prior(10, v_lb = v_lb, v_ub = v_ub),
          v_lb     = v_lb,
          v_ub     = v_ub)

# Define functions --------------------------------------------------------
## Coverage analysis ------------------------------------------------------
# Coverage function
# m_params: Matrix containing sample of parameter sets
run_coverage <- function(m_params){
```

```
  # Number of simulations
  n_sim <- nrow(m_params)

  # Run model
  df_model <- data.frame(NULL) # empty data.frame to store results
  for(iter_i in 1:n_sim){ # iter_i = 1

    # Run model
    l_model_out <- calibration_out(v_params = m_params[iter_i, ])

    # Output of the model in a data.frame
    # Modify the next lines based on your model
    df_temp <- rbind(l_model_out$df_Surv,
                     l_model_out$df_Prev,
                     l_model_out$df_PropSick) %>%
      mutate(iter = iter_i)

    # Gather results
    df_model <- rbind(df_model, df_temp)

    # Print progress
    if(iter_i/100==round(iter_i/100,0)) {
      cat('\r',paste(round(iter_i/n_sim*100,0),"% simulations done",sep=""))
    }
  }

  # Summarise
  df_model_summ <- df_model %>%
    group_by(time, Outcome, abbrev_outcome, type) %>%
    summarise(mean_val = mean(value),
              lb       = quantile(value,probs = 0.025),
              ub       = quantile(value, probs = 0.975),
              se       = sd(value)) %>%
    ungroup() %>%
    rename(value = mean_val)

  return(list(df_model        = df_model,
              df_model_summ   = df_model_summ))
}

## Sensitivity analysis ---------------------------------------------------
# Cost function
# v_params: Vector of model parameters.
costFun <- function(v_params) {

  # Run model
  df_model_out <- sick_sicker_cstm(v_params)

  # Filter and select columns
  ## Surv
  df_model_out_Surv <- df_model_out %>%
    filter(time %in% v_times) %>%
    select(time, Surv)

  df_targets_Surv <- df_targets %>%
    filter(abbrev_outcome == "Surv") %>%
    select(time, value, se) %>%
    rename(Surv = value)

  ## PropSick
  df_model_out_PropSick <- df_model_out %>%
    filter(time %in% v_times) %>%
```

```r
    select(time, PropSick)

  df_targets_PropSick <- df_targets %>%
    filter(abbrev_outcome == "PropSick") %>%
    select(time, value, se) %>%
    rename(PropSick = value)

  ## Prev
  df_model_out_Prev <- df_model_out %>%
    filter(time %in% v_times) %>%
    select(time, Prev)

  df_targets_Prev <- df_targets %>%
    filter(abbrev_outcome == "Prev") %>%
    select(time, value, se) %>%
    rename(Prev = value)

  # Compute "costs" for each target
  cost <- modCost(model = df_model_out_Surv,
                  obs   = df_targets_Surv,
                  err   = "se")

  cost <- modCost(model = df_model_out_Prev,
                  obs   = df_targets_Prev,
                  err   = "se",
                  cost  = cost)

  cost <- modCost(model = df_model_out_PropSick,
                  obs   = df_targets_PropSick,
                  err   = "se",
                  cost  = cost)
  return(cost)
}

# Function to compute model outputs for global sensitivity analyses
# m_params:     Matrix containing sample of parameter sets
# outcome_name: Target outcome c("Survival probability","Prevalence",
#               "Proportion of sick")
# n_time:       Time (year) for which GSA is to be computed
model_out_GSA <- function(m_params, outcome_name, n_time){

  # Empty vector to store results
  v_res <- c()

  if(outcome_name == "Survival probability"){

    for(i in 1:nrow(m_params)){ # i = 1

      # Run model
      df_model_out <- sick_sicker_cstm(m_params[i,])

      # Data.frame for each calibration target
      # Survival
      df_Surv <- data.frame(time = df_model_out$time,
                            Outcome = "Survival",
                            abbrev_outcome = "Surv",
                            type = "Model",
                            value = df_model_out$Surv) %>%
        filter(time %in% n_time)

      # Store result
      v_res <- c(v_res, df_Surv$value)
```

```r
    }
  }else if(outcome_name == "Prevalence"){

    for(i in 1:nrow(m_params)){ # i = 1

      # Run model
      df_model_out <- sick_sicker_cstm(m_params[i,])

      # Data.frame for each calibration target
      # Prevalence
      df_Prev <- data.frame(time = df_model_out$time,
                            Outcome = "Prevalence",
                            abbrev_outcome = "Prev",
                            type = "Model",
                            value = df_model_out$Prev) %>%
        filter(time %in% n_time)

      # Store result
      v_res <- c(v_res, df_Prev$value)

    }
  }else if(outcome_name == "Proportion of Sick"){

    for(i in 1:nrow(m_params)){ # i = 1

      # Run model
      df_model_out <- sick_sicker_cstm(m_params[i,])

      # Data.frame for each calibration target
      # Proportion who are Sick ("PropSick") among all those
      # afflicted (Sick+Sicker)
      df_PropSick <- data.frame(time = df_model_out$time,
                                Outcome = "Proportion of Sick",
                                abbrev_outcome = "PropSick",
                                type = "Model",
                                value = df_model_out$PropSick) %>%
        filter(time %in% n_time)

      # Store result
      v_res <- c(v_res, df_PropSick$value)

    }
  }

  # Return vector
  return(v_res)
}

# Global sensitivity analysis (GSA): this function runs soboljansen function
# from sensitivity package.
# v_outcomes: Vector containing the name of the targets.
# v_times:    Vector containing the times (in years).
# X1:         First random sample (prior distribution).
# X2:         Second random sample (prior distribution).
# n_boot:     Number of bootstrap replicates
# n_conf:     Confidence level for bootstrap confidence intervals.
run_GSA <- function(v_outcomes, v_times, X1, X2, n_boot, n_conf){

  df_res_sobol <- data.frame(NULL)
  for(outcome_i in v_outcomes){ # outcome_i = "Prevalence"

    for(time_i in v_times){ # time_i = 20
```

```r
      cat(outcome_i, time_i, "\n")

      # Compute Sobol Indices
      res_sobol <- soboljansen(model = model_out_GSA, X1 = X1, X2 = X2,
                               outcome_name = outcome_i, n_time = time_i,
                               nboot = n_boot, conf = n_conf)

      # Bind results
      df_res_sobol <- rbind(df_res_sobol,
                            data.frame(par = rownames(res_sobol$S),
                                       Outcome = outcome_i,
                                       type = "Main effect",
                                       x = time_i,
                                       res_sobol$S, row.names = NULL),
                            data.frame(par = rownames(res_sobol$T),
                                       Outcome = outcome_i,
                                       type = "Total effect",
                                       x = time_i,
                                       res_sobol$T, row.names = NULL))

    }
  }
  return(df_res_sobol)
}

# Compare sensitivity analysis --------------------------------------------
# Define input search space
# Range on input search space
v_lb_Sens <- c(p_S1S2 = 0.00, hr_S1 = 1.00, hr_S2 = 1.00) # lower bound
v_ub_Sens <- c(p_S1S2 = 0.50, hr_S1 = 8.00, hr_S2 = 25.0) # upper bound

# Get two sets of priors
set.seed(n_seed)
m_params_Sens_1 <- sample_prior(n_samp = n_sim,
                                v_lb   = v_lb_Sens,
                                v_ub   = v_ub_Sens)

m_params_Sens_2 <- sample_prior(n_samp = n_sim,
                                v_lb   = v_lb_Sens,
                                v_ub   = v_ub_Sens)

# Run GSA
df_res_sobol <- run_GSA(v_outcomes = as.character(v_outcome_names),
                        v_times    = v_times,
                        X1         = m_params_Sens_1,
                        X2         = m_params_Sens_2,
                        n_boot     = 100,
                        n_conf     = 0.95)

# Bootstrap confidence interval bounds were manually restricted to
# the plausible range [0,1]
df_res_sobol <- df_res_sobol %>%
  mutate(min..c.i. = ifelse(min..c.i.<0, 0, min..c.i.),
         max..c.i. = ifelse(max..c.i.>1, 1, max..c.i.))

# Plot
plot_GSA(df_res_sobol)

# Local sensitivity
SensRes <- sensFun(func  = costFun,
                   parms = colMeans(m_params_Sens_1) # mean values
)
```

```
# Plot
plot_Sfun(SensRes)

# Implement PRE-CISE (narrow) ---------------------------------------------
# 0. Define input search space and plot first coverage
v_lb_1 <- c(p_S1S2 = 0.10, hr_S1 = 1.0, hr_S2 = 1.0) # lower bound
v_ub_1 <- c(p_S1S2 = 0.20, hr_S1 = 1.7, hr_S2 = 2.0) # upper bound

# Run initial coverage analysis
set.seed(n_seed)  # set seed
m_params_1 <- sample_prior(n_samp = n_sim,
                           v_lb   = v_lb_1,
                           v_ub   = v_ub_1)

l_coverage_1  <- run_coverage(m_params = m_params_1)
df_coverage_1 <- l_coverage_1$df_model_summ %>%
  mutate(type_coverage = "Initial model coverage")

plot_coverage(df_targets  = df_targets,
              df_coverage = df_coverage_1)

## Resize the prior distribution bounds -----------------------------------
### Adjust the prob. to become sicker when sick: p_S1S2 --------------------
# 1. Compute model-data residuals on initial coverage (mean values)
v_params_1 <- colMeans(m_params_1)
modCost1   <- costFun(v_params_1)

# Plot residuals
plot_costFun(Cfun = modCost1, wgt_res = T) # weighted residuals
plot_costFun(modCost1, wgt_res = F)        # unweighted residuals

# 2. Local sensitivity analysis
SensRes1 <- sensFun(func  = costFun,
                    parms = v_params_1)

# Plot sensitivity
plot_Sfun(SensRes1)

# Save plot
plot_Sfun(SensRes1)

# Rank parameters according to their importance
m_Sens1 <- summary(SensRes1)

# value: value of the parameter
# Mean: mean sensitivity
# L1: L1 norm
# L2: L2 norm
df_Sens1 <- as.data.frame(m_Sens1)

# Order by L2
df_Sens1[order(df_Sens1$L2, decreasing = T),]

# 3. Compute the change in the prior distribution bounds based on the
# sensitivity analysis
df_changes_1 <- data.frame(NULL)
for(target_i in c("PropSick","Prev")){ # target_i = "PropSick"

  # Filter elasticities
  df_epsi <- SensRes1 %>%
    filter(var == target_i) %>%
    select(x,p_S1S2) %>%
```

```
    rename(epsilon = p_S1S2)

  # Change in y --> change in par
  df_temp <- modCost1$residuals %>%
    filter(name == target_i) %>%
    select(name,x,obs,mod) %>%
    mutate(y_change = ifelse(mod != 0, (obs-mod)/mod, 0)) %>%
    left_join(df_epsi,
              by = "x") %>%
    mutate(par_change = y_change/epsilon,
           par_change_lb = ifelse((1+par_change)*v_lb_1["p_S1S2"]<0,
                                  NA,(1+par_change)*v_lb_1["p_S1S2"]))

  # Bind data
  df_changes_1 <- rbind(df_changes_1,
                        df_temp)

}

# Percent change for lower or upper bound
n_change_lb <- min(df_changes_1$par_change_lb, na.rm = T)

# Resize bounds
v_lb_2 <- v_lb_1
v_ub_2 <- v_ub_1
v_lb_2["p_S1S2"] <- n_change_lb

# Run coverage
set.seed(n_seed)  # set seed
m_params_2 <- sample_prior(n_samp = n_sim,
                           v_lb = v_lb_2,
                           v_ub = v_ub_2)
l_coverage_2 <- run_coverage(m_params = m_params_2)
df_coverage_2 <- l_coverage_2$df_model_summ %>%
  mutate(type_coverage =
           "A: Improved model coverage by resizing the probability to become Sicker when Sick")

plot_coverage(df_targets  = df_targets,
              df_coverage = rbind(df_coverage_1,df_coverage_2))

### Adjust the hazard ratio for death in Sicker: hr_S2 --------------------
# 1. Compute model-data residuals on previous coverage (mean values)
v_params_2 <- colMeans(m_params_2)        # mean values
modCost2   <- costFun(v_params_2)

# 2. Local sensitivity analysis
SensRes2 <- sensFun(func  = costFun,
                    parms = v_params_2)

# Plot sensitivity
plot_Sfun(SensRes2)

# Rank parameters according to their importance
m_Sens2 <- summary(SensRes2)

# value: value of the parameter
# Mean: mean sensitivity
# L1: L1 norm
# L2: L2 norm
df_Sens2 <- as.data.frame(m_Sens2)

# Order by L2
```

```r
df_Sens2[order(df_Sens2$L2, decreasing = T),]

# 3. Compute the change in the prior distribution bounds based on the
# sensitivity analysis
df_changes_2 <- data.frame(NULL)
for(target_i in c("Surv","Prev")){ # target_i = "PropSick"

  # Filter elasticities
  df_epsi <- SensRes2 %>%
    filter(var == target_i) %>%
    select(x,hr_S2) %>%
    rename(epsilon = hr_S2)

  # Change in y --> change in par
  df_temp <- modCost2$residuals %>%
    filter(name == target_i) %>%
    select(name,x,obs,mod) %>%
    mutate(y_change = ifelse(mod != 0, (obs-mod)/mod, 0)) %>%
    left_join(df_epsi,
              by = "x") %>%
    mutate(par_change = y_change/epsilon,
           par_change_ub = ifelse((1+par_change)*v_ub_2["hr_S2"]<v_lb_2["hr_S2"],
                                  NA, (1+par_change)*v_ub_2["hr_S2"]))

  # Bind data
  df_changes_2 <- rbind(df_changes_2,
                        df_temp)

}

# Percent change for lower or upper bound
n_change_ub <- max(df_changes_2$par_change_ub, na.rm = T)

# Resize bounds
v_lb_3 <- v_lb_2
v_ub_3 <- v_ub_2
v_ub_3["hr_S2"] <- n_change_ub

# Run coverage
set.seed(n_seed)  # set seed
m_params_3 <- sample_prior(n_samp = n_sim,
                           v_lb   = v_lb_3,
                           v_ub   = v_ub_3)
l_coverage_3 <- run_coverage(m_params = m_params_3)
df_coverage_3 <- l_coverage_3$df_model_summ %>%
  mutate(type_coverage =
           "B: A + improved model coverage by resizing the hazard ratio for death in Sicker")

plot_coverage(df_targets  = df_targets,
              df_coverage = rbind(df_coverage_1,df_coverage_2,df_coverage_3))

### Adjust the hazard ratio for death in Sick: hr_S1 ----------------------
# 1. Compute model-data residuals on previous coverage (mean values)
v_params_3 <- colMeans(m_params_3)      # mean values
modCost3   <- costFun(v_params_3)

# 2. Local sensitivity analysis
SensRes3 <- sensFun(func  = costFun,
                    parms = v_params_3)

# Plot sensitivity
plot_Sfun(SensRes3)
```

```r
# 3. Compute the change in the prior distribution bounds based on the
# sensitivity analysis
df_changes_3 <- data.frame(NULL)
for(target_i in c("Surv")){ # target_i = "PropSick"

  # Filter elasticities
  df_epsi <- SensRes3 %>%
    filter(var == target_i) %>%
    select(x,hr_S1) %>%
    rename(epsilon = hr_S1)

  # Change in y --> change in par
  df_temp <- modCost3$residuals %>%
    filter(name == target_i) %>%
    select(name,x,obs,mod) %>%
    mutate(y_change = ifelse(mod != 0, (obs-mod)/mod, 0)) %>%
    left_join(df_epsi,
              by = "x") %>%
    mutate(par_change = y_change/epsilon,
           par_change_ub = ifelse((1+par_change)*v_ub_3["hr_S1"]<v_lb_3["hr_S1"],
                                  NA, (1+par_change)*v_ub_3["hr_S1"]))

  # Bind data
  df_changes_3 <- rbind(df_changes_3,
                        df_temp)

}

# Percent change for lower or upper bound
n_change_ub <- max(df_changes_3$par_change_ub, na.rm = T)

# Resize bounds
v_lb_4 <- v_lb_3
v_ub_4 <- v_ub_3
v_ub_4["hr_S1"] <- n_change_ub

# Run coverage
set.seed(n_seed)  # set seed
m_params_4 <- sample_prior(n_samp = n_sim,
                           v_lb   = v_lb_4,
                           v_ub   = v_ub_4)
l_coverage_4 <- run_coverage(m_params = m_params_4)
df_coverage_4 <- l_coverage_4$df_model_summ %>%
  mutate(type_coverage =
           "C: A + B + improved model coverage by resizing the hazard ratio for death in Sick")

plot_coverage(df_targets  = df_targets,
              df_coverage = rbind(df_coverage_1,
                                  df_coverage_2,
                                  df_coverage_3,
                                  df_coverage_4))

### Readjust the hazard ratio for death in Sick: hr_S1 --------------------
# 1. Compute model-data residuals on previous coverage (mean values)
v_params_4 <- colMeans(m_params_4)      # mean values
modCost4   <- costFun(v_params_4)

# 2. Local sensitivity analysis
SensRes4 <- sensFun(func  = costFun,
                    parms = v_params_4)

# Plot sensitivity
```

```
plot_Sfun(SensRes4)

# 3. Compute the change in the prior distribution bounds based on the
# sensitivity analysis
df_changes_4 <- data.frame(NULL)
for(target_i in c("Surv")){ # target_i = "PropSick"

  # Filter elasticities
  df_epsi <- SensRes4 %>%
    filter(var == target_i) %>%
    select(x,hr_S1) %>%
    rename(epsilon = hr_S1)

  # Change in y --> change in par
  df_temp <- modCost4$residuals %>%
    filter(name == target_i) %>%
    select(name,x,obs,mod) %>%
    mutate(y_change = ifelse(mod != 0, (obs-mod)/mod, 0)) %>%
    left_join(df_epsi,
              by = "x") %>%
    mutate(par_change = y_change/epsilon,
           par_change_ub = ifelse((1+par_change)*v_ub_4["hr_S1"]<v_lb_4["hr_S1"],
                                  NA, (1+par_change)*v_ub_4["hr_S1"]))

  # Bind data
  df_changes_4 <- rbind(df_changes_4,
                        df_temp)

}

# Percent change for lower or upper bound
n_change_ub <- max(df_changes_4$par_change_ub, na.rm = T)

# Resize bounds
v_lb_5 <- v_lb_4
v_ub_5 <- v_ub_4
v_ub_5["hr_S1"] <- n_change_ub

# Run coverage
set.seed(n_seed)  # set seed
m_params_5 <- sample_prior(n_samp = n_sim,
                           v_lb   = v_lb_5,
                           v_ub   = v_ub_5)
l_coverage_5 <- run_coverage(m_params = m_params_5)
df_coverage_5 <- l_coverage_5$df_model_summ %>%
  mutate(type_coverage =
           "D: A + B + C + improved model coverage by resizing the hazard ratio for death in Sick")

plot_coverage(df_targets  = df_targets,
              df_coverage = rbind(df_coverage_1,
                                  df_coverage_2,
                                  df_coverage_3,
                                  df_coverage_4,
                                  df_coverage_5))

### Readjust the hazard ratio for death in Sick: hr_S1 --------------------
# 1. Compute model-data residuals on previous coverage (mean values)
v_params_5 <- colMeans(m_params_5)      # mean values
modCost5   <- costFun(v_params_5)

# 2. Local sensitivity analysis
SensRes5 <- sensFun(func  = costFun,
```

```r
                    parms = v_params_5)

# Plot sensitivity
plot_Sfun(SensRes5)

# 3. Compute the change in the prior distribution bounds based on the
# sensitivity analysis
df_changes_5 <- data.frame(NULL)
for(target_i in c("Surv")){ # target_i = "PropSick"

  # Filter elasticities
  df_epsi <- SensRes5 %>%
    filter(var == target_i) %>%
    select(x,hr_S1) %>%
    rename(epsilon = hr_S1)

  # Change in y --> change in par
  df_temp <- modCost5$residuals %>%
    filter(name == target_i) %>%
    select(name,x,obs,mod) %>%
    mutate(y_change = ifelse(mod != 0, (obs-mod)/mod, 0)) %>%
    left_join(df_epsi,
              by = "x") %>%
    mutate(par_change = y_change/epsilon,
           par_change_ub = ifelse((1+par_change)*v_ub_5["hr_S1"]<v_lb_5["hr_S1"],
                                  NA, (1+par_change)*v_ub_5["hr_S1"]))

  # Bind data
  df_changes_5 <- rbind(df_changes_5,
                        df_temp)

}

# Percent change for lower or upper bound
n_change_ub <- max(df_changes_5$par_change_ub, na.rm = T)

# Resize bounds
v_lb_6 <- v_lb_5
v_ub_6 <- v_ub_5
v_ub_6["hr_S1"] <- n_change_ub

# Run coverage
set.seed(n_seed)  # set seed
m_params_6 <- sample_prior(n_samp = n_sim,
                           v_lb   = v_lb_6,
                           v_ub   = v_ub_6)
l_coverage_6 <- run_coverage(m_params = m_params_6)
df_coverage_6 <- l_coverage_6$df_model_summ %>%
  mutate(type_coverage =
           "E: A + B + C + D + improved model coverage by resizing the hazard ratio for death in Sick")

plot_coverage(df_targets  = df_targets,
              df_coverage = rbind(df_coverage_1,
                                  df_coverage_2,
                                  df_coverage_3,
                                  df_coverage_4,
                                  df_coverage_5,
                                  df_coverage_6))

### Final coverage --------------------------------------------------------
# Range on input search space after resize bounds
v_lb_final <- v_lb_6
```

```r
v_ub_final <- v_ub_6

set.seed(n_seed)  # set seed
m_params_final <- sample_prior(n_samp = n_sim,
                               v_lb   = v_lb_final,
                               v_ub   = v_ub_final)

l_coverage_final <- run_coverage(m_params = m_params_final)
df_coverage_final <- l_coverage_final$df_model_summ

# Plot final coverage
plot_model_out_vs_targets(df_targets = df_targets,
                          df_model   = df_coverage_final)

## Identifiability (Collinearity) analysis --------------------------------
# Run local sensitivity analysis
SensRes <- sensFun(func  = costFun,
                   parms = colMeans(m_params_final) # mean values
)

# Combination of variables
m_vars <- matrix(1, ncol = n_outcomes, nrow = n_outcomes,
                 dimnames = list(names(v_outcome_names),
                                 names(v_outcome_names)))

m_vars[lower.tri(m_vars)] <- 0

# Empty data.frame to store results
df_collin <- data.frame(NULL)

# Run collinearity analysis on all combinations of outcomes
for(i in 1:n_outcomes){ # i = 1
  for(j in 1:n_outcomes){ # j = 2

    if(m_vars[i,j] != 1) next

    if(i == j){

      # Get variable names
      var_i <- rownames(m_vars)[i]
      outcome_i <- v_outcome_names[which(names(v_outcome_names) == var_i)]

      # Compute collinearity analysis
      df_temp <- collin(SensRes, which = var_i) %>%
        mutate(var = var_i,
               Outcome = outcome_i)

    }else{

      # Get variable names
      var_i <- rownames(m_vars)[i]
      outcome_i <- v_outcome_names[which(names(v_outcome_names) == var_i)]
      var_j <- colnames(m_vars)[j]
      outcome_j <- v_outcome_names[which(names(v_outcome_names) == var_j)]

      # Compute collinearity analysis
      df_temp <- collin(SensRes, which = c(var_i,var_j)) %>%
        mutate(var = paste(var_i,var_j, sep = "_"),
               Outcome = paste(outcome_i,outcome_j, sep = " & "))
    }

    # Bind data.frames
```

```r
    df_collin <- rbind(df_collin, df_temp)
  }
}

# All variables
df_collin <- rbind(df_collin,
                   collin(SensRes) %>%
                     mutate(var = "All",
                            Outcome = paste0(v_outcome_names,collapse = " & ")))

# Plot collinearity analysis
plot_CollIndx(df_collin = df_collin, log_y = T)

## Calibrate with PRE-CISE ---------------------------------------------
# Range on input search space
v_lb <- v_lb_final # lower bound
v_ub <- v_ub_final # upper bound

# Specify seed (for reproducible sequence of random numbers)
set.seed(n_seed)

# number of random samples
n_resamp <- 10000

# record start time of calibration
t_init <- Sys.time()

# Bayesian calibration using the IMIS algorithm (Raftery & Bao, 2010)
fit_imis <- IMIS(B        = 1000,      # the incremental sample size at each iteration of IMIS
                 B.re     = n_resamp,  # the desired posterior sample size
                 number_k = 30,        # the maximum number of iterations in IMIS
                 D        = 0,
                 v_lb     = v_lb,
                 v_ub     = v_ub)

# Calculate computation time
comp_time <- Sys.time() - t_init
comp_time

# Obtain draws from posterior
m_calib_res <- fit_imis$resample

# Calculate log-likelihood (overall fit) and posterior probability of each sample
m_calib_res <- cbind(m_calib_res,
                     "Overall_fit"    = log_lik(m_calib_res[,v_param_names]),
                     "Posterior_prob" = posterior(m_calib_res[,v_param_names],
                                                  v_lb = v_lb,
                                                  v_ub = v_ub))

# normalize posterior probability
m_calib_res[,"Posterior_prob"] <- m_calib_res[,"Posterior_prob"]/
  sum(m_calib_res[,"Posterior_prob"])

### Exploring best-fitting input sets --------------------------------------
# Plot the 10000 draws from the posterior
v_post_color <- scales::rescale(m_calib_res[,"Posterior_prob"])
s3d <- scatterplot3d(x = m_calib_res[, 1],
                     y = m_calib_res[, 2],
                     z = m_calib_res[, 3],
                     color = scales::alpha("black", v_post_color),
                     xlim = c(v_lb[1],v_ub[1]),
                     ylim = c(v_lb[2],v_ub[2]),
```

```
                     zlim = c(v_lb[3],v_ub[3]),
                     xlab = v_param_names[1],
                     ylab = v_param_names[2],
                     zlab = v_param_names[3])
# add center of Gaussian components
s3d$points3d(fit_imis$center, col = "red", pch = 8)

# Plot the 1000 draws from the posterior with marginal histograms
pairs.panels(m_calib_res[,v_param_names])

# Compute posterior mean
v_calib_post_mean <- colMeans(m_calib_res[,v_param_names])
v_calib_post_mean

# Compute posterior median and 95% credible interval
m_calib_res_95cr <- colQuantiles(m_calib_res[,v_param_names],
                                 probs = c(0.025, 0.5, 0.975))
m_calib_res_95cr

### Plot maximum-a-posteriori (MAP) ---------------------------------------
# Compute MAP parameter set
v_calib_map <- m_calib_res[which.max(m_calib_res[,"Posterior_prob"]),]

# Run model with MAP
df_out_best <- sick_sicker_cstm(v_calib_map[v_param_names])

# TARGET 1: Survival ("Surv")
plotrix::plotCI(x = l_targets$Surv$time, y = l_targets$Surv$value,
                ui = l_targets$Surv$ub,
                li = l_targets$Surv$lb,
                ylim = c(0, 1),
                xlab = "Time", ylab = "Pr Survive")
points(x = df_out_best$time,
       y = df_out_best$Surv,
       pch = 8, col = "red")
legend("topright",
       legend = c("Target", "Model-predicted output"),
       col = c("black", "red"), pch = c(1, 8))

# TARGET 2: "Prev"
plotrix::plotCI(x = l_targets$Prev$time, y = l_targets$Prev$value,
                ui = l_targets$Prev$ub,
                li = l_targets$Prev$lb,
                ylim = c(0, 1),
                xlab = "Time", ylab = "Prev")
points(x = df_out_best$time,
       y = df_out_best$Prev,
       pch = 8, col = "red")
legend("topright",
       legend = c("Target", "Model-predicted output"),
       col = c("black", "red"), pch = c(1, 8))

# TARGET 3: "PropSick"
plotrix::plotCI(x = l_targets$PropSick$time, y = l_targets$PropSick$value,
                ui = l_targets$PropSick$ub,
                li = l_targets$PropSick$lb,
                ylim = c(0, 1),
                xlab = "Time", ylab = "PropSick")
points(x = df_out_best$time,
       y = df_out_best$PropSick,
       pch = 8, col = "red")
legend("topright",
```

```
       legend = c("Target", "Model-predicted output"),
       col = c("black", "red"), pch = c(1, 8))

### Propagate calibrated parameter uncertainty ----------------------------
# Compute IMIS posterior predicted outputs
m_out_surv     <- matrix(NA, nrow = n_resamp, ncol = length(l_targets$Surv$value))
m_out_prev     <- matrix(NA, nrow = n_resamp, ncol = length(l_targets$Prev$value))
m_out_propsick <- matrix(NA, nrow = n_resamp, ncol = length(l_targets$PropSick$value))

# Run model for each posterior parameter set
for(i in 1:n_resamp){ # i = 1
  df_model_res_temp <- sick_sicker_cstm(m_calib_res[i, ])
  m_out_surv[i, ] <- df_model_res_temp$Surv
  m_out_prev[i, ] <- df_model_res_temp$Prev
  m_out_propsick[i, ] <- df_model_res_temp$PropSick[df_model_res_temp$time %in% v_times]
  if(i/100==round(i/100,0)) {
    cat('\r',paste(i/n_resamp*100,"% done",sep=""))
  }
}

# Posterior predicted mean
m_out_surv_postmean <- colMeans(m_out_surv)
m_out_prev_postmean <- colMeans(m_out_prev)
m_out_propsick_postmean <- colMeans(m_out_propsick)

# Implement PRE-CISE (aggressive approach) --------------------------------
# 0. Define input search space and plot first coverage
v_lb_1 <- c(p_S1S2 = 0.00, hr_S1 = 1.00, hr_S2 = 1.00) # lower bound
v_ub_1 <- c(p_S1S2 = 0.50, hr_S1 = 8.00, hr_S2 = 25.0) # upper bound

# Run initial coverage analysis
set.seed(n_seed)   # set seed
m_params_1 <- sample_prior(n_samp = n_sim,
                           v_lb   = v_lb_1,
                           v_ub   = v_ub_1)

l_coverage_1  <- run_coverage(m_params = m_params_1)
df_coverage_1 <- l_coverage_1$df_model_summ %>%
  mutate(type_coverage = "Initial model coverage")

plot_coverage(df_targets  = df_targets,
              df_coverage = df_coverage_1)

## Resize the prior distribution bounds -----------------------------------
### Adjust the probability to become Sicker when Sick: p_S1S2 -------------
# 1. Compute the effect of the parameters on the variables of interest
# Compute model costs on initial coverage (mean values)
v_params_1 <- colMeans(m_params_1)
modCost1 <- costFun(v_params_1)

# Plot residuals
# plot_costFun(modCost1, wgt_res = T) # weighted residuals
# plot_costFun(modCost1, wgt_res = F) # unweighted residuals

# 2. Local sensitivity analysis
SensRes1 <- sensFun(func  = costFun,
                    parms = v_params_1)

# Plot sensitivity*
plot_Sfun(SensRes1)

# Rank parameters according to their importance
```

```
m_Sens1 <- summary(SensRes1)

# value: value of the parameter
# Mean: mean sensitivity
# L1: L1 norm
# L2: L2 norm
df_Sens1 <- as.data.frame(m_Sens1)

# Order by mean sensitivity
df_Sens1[order(abs(df_Sens1$L2), decreasing = T),]

# 3. Resize the prior distribution bounds
# Start with the one the outcomes are most sensible to: p_S1S2
df_changes_1 <- data.frame(NULL)
for(target_i in c("PropSick","Prev")){ # target_i = "PropSick"

  # Filter elasticities
  df_epsi <- SensRes1 %>%
    filter(var == target_i) %>%
    select(x,p_S1S2) %>%
    rename(epsilon = p_S1S2)

  # Change in y --> change in par
  df_temp <- modCost1$residuals %>%
    filter(name == target_i) %>%
    select(name,x,obs,mod) %>%
    mutate(y_change = ifelse(mod != 0, (obs-mod)/mod, 0)) %>%
    left_join(df_epsi,
              by = "x") %>%
    mutate(par_change = y_change/epsilon,
           par_change_ub = ifelse((1+par_change)*v_ub_1["p_S1S2"] < v_lb_1["p_S1S2"],
                                  NA, (1+par_change)*v_ub_1["p_S1S2"]))

  # Bind data
  df_changes_1 <- rbind(df_changes_1,
                        df_temp)

}

# Percent change for lower or upper bound
n_change_ub <- max(df_changes_1$par_change_ub, na.rm = T)

# Resize bounds
v_lb_2 <- v_lb_1
v_ub_2 <- v_ub_1
v_ub_2["p_S1S2"] <- n_change_ub

# Run coverage
set.seed(n_seed)  # set seed
m_params_2 <- sample_prior(n_samp = n_sim,
                           v_lb = v_lb_2,
                           v_ub = v_ub_2)
l_coverage_2 <- run_coverage(m_params = m_params_2)
df_coverage_2 <- l_coverage_2$df_model_summ %>%
  mutate(type_coverage =
           "A: Improved model coverage by resizing the probability to become Sicker when Sick")

plot_coverage(df_targets  = df_targets,
              df_coverage = rbind(df_coverage_1,df_coverage_2))

### Adjust the hazard ratio for death in Sicker: hr_S2 --------------------
# 1. Compute model-data residuals on previous coverage (mean values)
```

```r
v_params_2 <- colMeans(m_params_2)
modCost2   <- costFun(v_params_2)

# 2. Local sensitivity analysis
SensRes2 <- sensFun(func  = costFun,
                    parms = v_params_2)

# Plot sensitivity
# plot_Sfun(SensRes2)

# Rank parameters according to their importance
m_Sens2 <- summary(SensRes2)

# value: value of the parameter
# Mean: mean sensitivity
# L1: L1 norm
# L2: L2 norm
df_Sens2 <- as.data.frame(m_Sens2)

# Order by L2
df_Sens2[order(df_Sens2$L2, decreasing = T),]

# 3. Compute the change in the prior distribution bounds based on the
# sensitivity analysis
df_changes_2 <- data.frame(NULL)
for(target_i in c("Surv","Prev")){ # target_i = "PropSick"

  # Filter elasticities
  df_epsi <- SensRes2 %>%
    filter(var == target_i) %>%
    select(x,hr_S2) %>%
    rename(epsilon = hr_S2)

  # Change in y --> change in par
  df_temp <- modCost2$residuals %>%
    filter(name == target_i) %>%
    select(name,x,obs,mod) %>%
    mutate(y_change = ifelse(mod != 0, (obs-mod)/mod, 0)) %>%
    left_join(df_epsi,
              by = "x") %>%
    mutate(par_change = y_change/epsilon,
           par_change_ub = ifelse((1+par_change)*v_ub_2["hr_S2"] < v_lb_2["hr_S2"],
                                  NA, (1+par_change)*v_ub_2["hr_S2"]))

  # Bind data
  df_changes_2 <- rbind(df_changes_2,
                        df_temp)
}

# Percent change for lower or upper bound
n_change_ub <- min(df_changes_2$par_change_ub, na.rm = T)

# Resize bounds
v_lb_3 <- v_lb_2
v_ub_3 <- v_ub_2
v_ub_3["hr_S2"] <- n_change_ub

# Run coverage
set.seed(n_seed)  # set seed
m_params_3 <- sample_prior(n_samp = n_sim,
                           v_lb   = v_lb_3,
                           v_ub   = v_ub_3)
```

```r
l_coverage_3 <- run_coverage(m_params = m_params_3)
df_coverage_3 <- l_coverage_3$df_model_summ %>%
  mutate(type_coverage =
           "B: A + improved model coverage by resizing the hazard ratio for death in Sicker")

plot_coverage(df_targets  = df_targets,
              df_coverage = rbind(df_coverage_1,df_coverage_2,df_coverage_3))

### Adjust the hazard ratio for death in Sick: hr_S1 -----------------------
# 1. Compute model-data residuals on previous coverage (mean values)
v_params_3 <- colMeans(m_params_3)      # mean values
modCost3   <- costFun(v_params_3)

# 2. Local sensitivity analysis
SensRes3 <- sensFun(func  = costFun,
                    parms = v_params_3)

# Plot sensitivity
# plot_Sfun(SensRes3)

# 3. Compute the change in the prior distribution bounds based on the
# sensitivity analysis
df_changes_3 <- data.frame(NULL)
for(target_i in c("Surv")){ # target_i = "PropSick"

  # Filter elasticities
  df_epsi <- SensRes3 %>%
    filter(var == target_i) %>%
    select(x,hr_S1) %>%
    rename(epsilon = hr_S1)

  # Change in y --> change in par
  df_temp <- modCost3$residuals %>%
    filter(name == target_i) %>%
    select(name,x,obs,mod) %>%
    mutate(y_change = ifelse(mod != 0, (obs-mod)/mod, 0)) %>%
    left_join(df_epsi,
              by = "x") %>%
    mutate(par_change = y_change/epsilon,
           par_change_ub = ifelse((1+par_change)*v_ub_3["hr_S1"] < v_lb_3["hr_S1"],
                                  NA, (1+par_change)*v_ub_3["hr_S1"]))

  # Bind data
  df_changes_3 <- rbind(df_changes_3,
                        df_temp)

}

# Percent change for lower or upper bound, survival t = 10
n_change_ub <- df_changes_3$par_change_ub[df_changes_3$x == 10]

# Resize bounds
v_lb_4 <- v_lb_3
v_ub_4 <- v_ub_3
v_ub_4["hr_S1"] <- n_change_ub

# Run coverage
set.seed(n_seed)  # set seed
m_params_4 <- sample_prior(n_samp = n_sim,
                           v_lb   = v_lb_4,
                           v_ub   = v_ub_4)
l_coverage_4 <- run_coverage(m_params = m_params_4)
```

```r
df_coverage_4 <- l_coverage_4$df_model_summ %>%
  mutate(type_coverage =
           "C: A + B + improved model coverage by resizing the hazard ratio for death in Sick")

l_plot_coverage <- plot_coverage(df_targets  = df_targets,
                                 df_coverage = rbind(df_coverage_1,
                                                     df_coverage_2,
                                                     df_coverage_3,
                                                     df_coverage_4))

### Final coverage ----------------------------------------------------------
# Range on input search space after resize bounds
v_lb_final <- v_lb_4
v_ub_final <- v_ub_4

set.seed(n_seed)   # set seed
m_params_final <- sample_prior(n_samp = n_sim,
                               v_lb   = v_lb_final,
                               v_ub   = v_ub_final)

l_coverage_final <- run_coverage(m_params = m_params_final)
df_coverage_final <- l_coverage_final$df_model_summ

plot_model_out_vs_targets(df_targets = df_targets,
                          df_model   = df_coverage_final)

# Implement PRE-CISE (conservative approach) ------------------------------
# 0. Define input search space and plot first coverage
v_lb_1 <- c(p_S1S2 = 0.00, hr_S1 = 1.00, hr_S2 = 1.00) # lower bound
v_ub_1 <- c(p_S1S2 = 0.50, hr_S1 = 8.00, hr_S2 = 25.0) # upper bound

# Run initial coverage analysis
set.seed(n_seed)   # set seed
m_params_1 <- sample_prior(n_samp = n_sim,
                           v_lb   = v_lb_1,
                           v_ub   = v_ub_1)

l_coverage_1  <- run_coverage(m_params = m_params_1)
df_coverage_1 <- l_coverage_1$df_model_summ %>%
  mutate(type_coverage = "Initial model coverage")

l_plot_coverage_1 <- plot_coverage(df_targets  = df_targets,
                                   df_coverage = df_coverage_1)
l_plot_coverage_1

## Resize the prior distribution bounds -----------------------------------
### Adjust the probability to become Sicker when Sick: p_S1S2 -------------
# 1. Compute the effect of the parameters on the variables of interest
# Compute model costs on initial coverage (mean values)
v_params_1 <- colMeans(m_params_1)
modCost1 <- costFun(v_params_1)

# Plot residuals
plot_costFun(modCost1, wgt_res = T) # weighted residuals
plot_costFun(modCost1, wgt_res = F) # unweighted residuals

# 2. Local sensitivity analysis
SensRes1 <- sensFun(func  = costFun,
                    parms = v_params_1)

# Plot sensitivity*
plot_Sfun(SensRes1)
```

```r
# Rank parameters according to their importance
m_Sens1 <- summary(SensRes1)

# value: value of the parameter
# Mean: mean sensitivity
# L1: L1 norm
# L2: L2 norm
df_Sens1 <- as.data.frame(m_Sens1)

# Order by mean sensitivity
df_Sens1[order(abs(df_Sens1$L2), decreasing = T),]

# 3. Resize the prior distribution bounds
df_changes_1 <- data.frame(NULL)
for(target_i in c("Surv","PropSick","Prev")){ # target_i = "PropSick"

  # Filter elasticities
  df_epsi <- SensRes1 %>%
    filter(var == target_i) %>%
    select(x,p_S1S2) %>%
    rename(epsilon = p_S1S2)

  # Change in y --> change in par
  df_temp <- modCost1$residuals %>%
    filter(name == target_i) %>%
    select(name,x,obs,mod) %>%
    mutate(y_change = ifelse(mod != 0, (obs-mod)/mod, 0)) %>%
    left_join(df_epsi,
              by = "x") %>%
    mutate(par_change = y_change/epsilon,
           par_change_ub = ifelse((1+par_change)*v_ub_1["p_S1S2"]<v_lb_1["p_S1S2"] |
                                    (1+par_change)*v_ub_1["p_S1S2"]>1,
                                  NA, (1+par_change)*v_ub_1["p_S1S2"]))

  # Bind data
  df_changes_1 <- rbind(df_changes_1,
                        df_temp)

}

# Percent change for lower or upper bound
n_change_ub <- max(df_changes_1$par_change_ub, na.rm = T)

# Resize bounds
v_lb_2 <- v_lb_1
v_ub_2 <- v_ub_1
v_ub_2["p_S1S2"] <- n_change_ub

# Run coverage
set.seed(n_seed)  # set seed
m_params_2 <- sample_prior(n_samp = n_sim,
                           v_lb = v_lb_2,
                           v_ub = v_ub_2)
l_coverage_2 <- run_coverage(m_params = m_params_2)
df_coverage_2 <- l_coverage_2$df_model_summ %>%
  mutate(type_coverage =
           "A: Improved model coverage by resizing the probability to become Sicker when Sick")

plot_coverage(df_targets  = df_targets,
              df_coverage = rbind(df_coverage_1,df_coverage_2))

### Adjust the hazard ratio for death in Sicker: hr_S2 --------------------
```

```r
# 1. Compute model-data residuals on previous coverage (mean values)
v_params_2 <- colMeans(m_params_2)
modCost2   <- costFun(v_params_2)

# 2. Local sensitivity analysis
SensRes2 <- sensFun(func  = costFun,
                    parms = v_params_2)

# Plot sensitivity
plot_Sfun(SensRes2)

# Rank parameters according to their importance
m_Sens2 <- summary(SensRes2)

# value: value of the parameter
# Mean: mean sensitivity
# L1: L1 norm
# L2: L2 norm
df_Sens2 <- as.data.frame(m_Sens2)

# Order by L2
df_Sens2[order(df_Sens2$L2, decreasing = T),]

# 3. Compute the change in the prior distribution bounds based on the
# sensitivity analysis
df_changes_2 <- data.frame(NULL)
for(target_i in c("Surv","Prev")){ # target_i = "PropSick"

  # Filter elasticities
  df_epsi <- SensRes2 %>%
    filter(var == target_i) %>%
    select(x,hr_S2) %>%
    rename(epsilon = hr_S2)

  # Change in y --> change in par
  df_temp <- modCost2$residuals %>%
    filter(name == target_i) %>%
    select(name,x,obs,mod) %>%
    mutate(y_change = ifelse(mod != 0, (obs-mod)/mod, 0)) %>%
    left_join(df_epsi,
              by = "x") %>%
    mutate(par_change = y_change/epsilon,
           par_change_ub = ifelse((1+par_change)*v_ub_2["hr_S2"]<v_lb_2["hr_S2"],
                                  NA,(1+par_change)*v_ub_2["hr_S2"]))

  # Bind data
  df_changes_2 <- rbind(df_changes_2,
                        df_temp)
}

# Percent change for lower or upper bound
n_change_ub <- mean(df_changes_2$par_change_ub, na.rm = T)

# Resize bounds
v_lb_3 <- v_lb_2
v_ub_3 <- v_ub_2
v_ub_3["hr_S2"] <- n_change_ub

# Run coverage
set.seed(n_seed)  # set seed
m_params_3 <- sample_prior(n_samp = n_sim,
                           v_lb   = v_lb_3,
```

```r
                           v_ub   = v_ub_3)
l_coverage_3 <- run_coverage(m_params = m_params_3)
df_coverage_3 <- l_coverage_3$df_model_summ %>%
  mutate(type_coverage =
           "B: A + improved model coverage by resizing the hazard ratio for death in Sicker")

l_plot_coverage <- plot_coverage(df_targets  = df_targets,
                                 df_coverage = rbind(df_coverage_1,
                                                     df_coverage_2,
                                                     df_coverage_3))

### Final coverage -------------------------------------------------------
# Range on input search space after resize bounds
v_lb_final <- v_lb_3
v_ub_final <- v_ub_3

set.seed(n_seed)   # set seed
m_params_final <- sample_prior(n_samp = n_sim,
                               v_lb   = v_lb_final,
                               v_ub   = v_ub_final)

l_coverage_final <- run_coverage(m_params = m_params_final)
df_coverage_final <- l_coverage_final$df_model_summ

plot_model_out_vs_targets(df_targets = df_targets,
                          df_model   = df_coverage_final)
```

**SIR_functions.R**

```r
# Model-decision functions -----------------------------------------------

#' Load all parameter sets
#'
#' \code{load_params_all} generates all the values of the SIR model parameters.
#'
#' @param n_t Time horizon in days.
#' @param time_step Model evaluations per day.
#' @param n_date_init Initial day on which the infections began.
#' @param n_inf_init Number of individuals initially infected.
#' @param n_tot_pop Total number of the population.
#' @param r_beta Transmission rate.
#' @param r_gamma Recovery rate.
#'
#' @returns List containing all the parameters of the decision model.
#'
#' @export
load_params_all <- function(n_t,
                            time_step,
                            n_date_init,
                            n_inf_init,
                            n_tot_pop,
                            r_beta,
                            r_gamma){

  # Vector of times
  v_times <- seq(0,n_t,time_step)

  # Initial state
  S_0 <- (n_tot_pop - n_inf_init)/n_tot_pop
  I_0 <- n_inf_init/n_tot_pop
```

```r
  R_0 <- 0

  v_states_init <- c(S = S_0,
                     I = I_0,
                     R = R_0)
  v_states_names <- names(v_states_init)

  return(list(n_t            = n_t,
              time_step      = time_step,
              v_times        = v_times,
              n_date_init    = n_date_init,
              v_states_init  = v_states_init,
              v_states_names = v_states_names,
              r_beta         = r_beta,
              r_gamma        = r_gamma))

}

#' Derivatives of the SIR model
#'
#' \code{SIR_dXdt} computes the derivatives of a Susceptible-Infectious-Recovered
#' (SIR) transmission model.
#'
#' @param time Time at which the derivatives will be computed.
#' @param v_pop Vector containing the population by health state
#' @param l_params_all List containing all the parameters of the decision model.
#'
#' @return
#' A list containing a vector of derivatives for each health state.
#'
#' @export
SIR_dXdt <- function(time, v_pop, l_params_all){

  # Getting parameters
  r_beta  <- l_params_all$r_beta
  r_gamma <- l_params_all$r_gamma

  # Population
  S <- v_pop["S"]
  I <- v_pop["I"]
  R <- v_pop["R"]

  # ODE model
  dS <- -r_beta*S*I
  dI <- r_beta*S*I - r_gamma*I
  dR <- r_gamma*I

  return(list(c(dS,dI,dR)))

}

#' SIR model
#'
#' \code{SIR_model} implements a Susceptible-Infectious-Recovered (SIR) model.
#'
#' @param l_params_all List containing all the parameters of the decision model.
#'
#' @return
#' A data.frame containing the population for each health state over time and
#' a list containing all the parameters of decision model.
#'
#' @export
```

```r
SIR_model <- function(l_params_all){
  df_out_model <- lsoda(y      = l_params_all$v_states_init,
                        times = l_params_all$v_times,
                        func  = SIR_dXdt,
                        parms = l_params_all)

  l_out_model <- list(df_out_model = as.data.frame(df_out_model),
                      l_params_all = l_params_all)

  return(l_out_model)
}

#' Total incident infections
#'
#' \code{calc_dxinc_totals} calculate total number of incident diagnosed
#' infections over time.
#'
#' @param l_out_model List containing the model output.
#' @param l_params_all List containing all the parameters of the decision model.
#'
#' @return
#' A data.frame containing the total number of incident diagnosed infections
#' over time.
#'
#' @export
calc_DXIncTot <- function(l_out_model, l_params_all){

  # Get variables
  n_date_init <- l_params_all$n_date_init

  # Get model output
  df_out_model <- l_out_model$df_out_model

  df_DXIncTot <- data.frame(Type = "Model",
                            abbrev_outcome = "DXIncTot",
                            Outcome = "Incident confirmed cases",
                            time = df_out_model$time,
                            Date = n_date_init + df_out_model$time,
                            value = df_out_model$I*n_tot_pop,
                            lb = NA,
                            ub = NA,
                            se = NA,
                            check.names = FALSE) %>%
    mutate(Date0 = Date - Date[1],
           Day = time + 1) %>%
    select(Type, abbrev_outcome, Outcome, Day, Date, Date0, value, lb, ub, se)

  return(df_DXIncTot)
}

# Visualization functions --------------------------------------------------

#' Plot model outputs vs targets
#'
#' \code{plot_model_out_vs_targets} plots model outputs vs targets.
#'
#' @param df_targets Data.frame with calibration targets.
#'
#' @return A ggplot2 object.
#' @export
plot_targets <- function(df_targets){
```

```r
  # Plot targets
  gg_targets <- ggplot(df_targets,
                       aes(x = Day, y = value, ymin = lb, ymax = ub)) +
    geom_point(shape = 1) + geom_errorbar() +
    scale_y_continuous(breaks = number_ticks(6)) +
    scale_x_continuous(breaks = number_ticks(10)) +
    # scale_x_date(date_labels = "%m/%d",
    #              breaks = number_ticks(10)) +
    ylab("Number of infections") + xlab("Time (days)") +
    theme_bw(base_size = 18) +
    theme(legend.position = "top",
          strip.background = element_rect(color = "white",
                                          fill = "white"),
          axis.text.x = element_text(angle = 0,
                                     vjust = 0.5,
                                     hjust = 0.5)) +
    coord_cartesian(ylim = c(0,800))

  return(gg_targets)

}

#' Plot model outputs vs targets
#'
#' \code{plot_model_out_vs_targets} plots model outputs vs targets.
#'
#' @param df_targets Data.frame containing the calibration targets.
#' @param df_model Data.frame containing the model outputs.
#'
#' @return A ggplot2 object.
#'
#' @export
plot_model_out_vs_targets <- function(df_targets, df_model){

  # Bind data.frames
  df_plot <- rbind(
    df_targets %>%
      select(Type, Outcome, Day, Date, Date0, value, lb, ub, se),
    df_model %>%
      select(Type, Outcome, Day, Date, Date0, value, lb, ub, se)
  )

  # Vector of dates
  v_dates <- as.Date(unique(df_plot$Date))

  # Order
  df_plot$Type <- factor(df_plot$Type, levels = c("Target","Model"))

  # Plot
  gg_model_out_vs_targets <- ggplot(df_plot,
                                    aes(x = Day, y = value, ymin = lb, ymax = ub,
                                        color = Type, shape = Type)) +
    geom_point(size = 1.6) +
    geom_errorbar(linewidth = 0.8) +
    scale_shape_manual(NULL, values = c(Target = 1,
                                        Model = 8)) +
    scale_color_manual(NULL, values = c(Target = "black",
                                        Model = "red")) +
    scale_y_continuous(breaks = number_ticks(6)) +
    scale_x_continuous(breaks = number_ticks(10)) +
    # scale_x_date(date_labels = "%m/%d",
    #              breaks = number_ticks(10)) +
```

```r
    ylab("Number of infections") + xlab("Time (days)") +
    theme_bw(base_size = 18) +
    theme(legend.position = "inside",
          legend.position.inside = c(0.15,0.85),
          strip.background = element_rect(color = "white",
                                          fill = "white"),
          axis.text.x = element_text(angle = 0,
                                     vjust = 0.5,
                                     hjust = 0.5)) +
    coord_cartesian(ylim = c(0,500))

  return(gg_model_out_vs_targets)

}

#' Plot coverage
#'
#' \code{plot_coverage} plots model outputs vs targets.
#'
#' @param df_targets Data.frame containing the calibration targets.
#' @param df_coverage Data.frame containing the coverage analysis.
#'
#' @return A ggplot2 object.
#'
#' @export
plot_coverage <- function(df_targets, df_coverage){

  # Add type_coverage to targets
  df_targets <- df_targets %>%
    mutate(type_coverage = case_when(Type == "Target" ~ "Calibration target"))

  # Get levels
  v_levels <- c("Calibration target", unique(df_coverage$type_coverage))

  # Bind data.frames
  df_model_target <- rbind(df_targets %>%
                             select(Type, Outcome, type_coverage, Day,
                                    Date, Date0, value, lb, ub, se),
                           df_coverage %>%
                             select(Type, Outcome, type_coverage, Day,
                                    Date, Date0, value, lb, ub, se))
  df_model_target$type_coverage <- factor(df_model_target$type_coverage,
                                          levels = v_levels)

  # Vector of shapes
  v_shape <- c(1,rep(NA,length(v_levels)-1))
  names(v_shape) <- v_levels

  # Vector of colors
  v_colors <- c("black","red","#FF8000","#56B4E9","#009E73","#CCBFFF","tan")
  names(v_colors) <- c(
    "Calibration target",
    "Calibration target sampled",
    "Initial model coverage",
    "A: Improved model coverage by resizing the transmission rate",
    "B: A + improved model coverage by resizing the recovery rate",
    "C: A + B + improved model coverage by resizing the transmission rate",
    "D: A + B + C + improved model coverage by resizing the recovery rate"
  )

  # Plot Survival ("Surv")
  gg_coverage <- ggplot(df_model_target,
```

```
                    aes(x = Day,
                        y = value, ymin = lb, ymax = ub,
                        color = type_coverage, shape = type_coverage)) +
    geom_point(size = 1.5, position = position_dodge(0.8)) +
    geom_errorbar(position = position_dodge(0.8), linewidth = 0.8) +
    scale_color_manual(NULL, values = v_colors) +
    scale_shape_manual(NULL, values = v_shape) +
    scale_y_continuous(breaks = number_ticks(6)) +
    scale_x_continuous(breaks = number_ticks(10)) +
    # scale_x_date(date_labels = "%m/%d",
    #              breaks = number_ticks(10)) +
    ylab("Number of infections") + xlab("Time (days)") +
    guides(color = guide_legend(ncol=1, byrow=TRUE)) +
    theme_bw(base_size = 18) +
    theme(legend.position = "inside",
          legend.position.inside = c(0.7,0.8),
          strip.background = element_rect(color = "white",
                                          fill = "white"),
          axis.text.x = element_text(angle = 0,
                                     vjust = 0.5,
                                     hjust = 0.5))  +
    coord_cartesian(ylim = c(0,820))

  return(gg_coverage)

}

#' Plot residuals of a model vs observations
#'
#' @param Cfun modCost object obtained from \code{costFun}.
#' @param wgt_res Flag (default is TRUE) of whether weighted
#' residuals should be plotted.
#'
#' @return A ggplot2 object.
#'
#' @export
plot_costFun <- function(Cfun, wgt_res = TRUE){

  df_residuals <- Cfun$residuals
  df_residuals$name_lab <- "Number of infections"

  if(wgt_res){

    plotCfun <- ggplot(df_residuals,
                       aes(x = x, y = res, color = name_lab, shape = name_lab)) +
      geom_point() +
      facet_wrap(~ name_lab, scales = "free_y") +
      scale_y_continuous(breaks = number_ticks(6)) +
      scale_x_continuous(breaks = number_ticks(6)) +
      ylab("Weighted residuals") + xlab("Time (days)") +
      theme_bw(base_size = 18) +
      theme(legend.position = "none",
            strip.background = element_rect(color = "white",
                                            fill = "white"),
            axis.text.x = element_text(angle = 0,
                                       vjust = 0.5,
                                       hjust = 0.5))

  }else{

    plotCfun <- ggplot(df_residuals,
                       aes(x = x, y = res.unweighted,
```

```r
                          color = name_lab, shape = name_lab)) +
      geom_point() +
      facet_wrap(~ name_lab, scales = "free_y") +
      scale_y_continuous(breaks = number_ticks(6)) +
      scale_x_continuous(breaks = number_ticks(6)) +
      ylab("Residuals") + xlab("Time (days)") +
      theme_bw(base_size = 18) +
      theme(legend.position = "none",
            strip.background = element_rect(color = "white",
                                            fill = "white"),
            axis.text.x = element_text(angle = 0,
                                       vjust = 0.5,
                                       hjust = 0.5))
  }

  return(plotCfun)
}

#' Plot sensitivity function
#'
#' \code{plot_Sfun} plots the sensitivity function.
#'
#' @param Sfun Sensitivity function obtained from \code{sensFun}.
#'
#' @return A ggplot2 object.
#'
#' @export
plot_Sfun <- function(Sfun){

  v_abb_targets <- unique(Sfun$var)
  v_param_names <- colnames(Sfun)[3:length(Sfun)]
  n_params <- length(v_param_names)

  # Generate long data.frame for plots
  df_Sfun_long <- data.frame(NULL)
  for(abb_targ_i in v_abb_targets){

    df_temp <- subset(Sfun, var == abb_targ_i) %>%
      pivot_longer(cols = all_of(v_param_names),
                   names_to = "parm",
                   values_to = "value")

    df_Sfun_long <- rbind(df_temp, df_Sfun_long)

  }
  df_plot <- df_Sfun_long %>%
    rename(Parameter = parm)

  # Categorical variables
  # Parameters
  df_plot$Parameter <- factor(df_plot$Parameter,
                              levels = v_param_names,
                              labels = c("Transmission rate",
                                         "Recovery rate"),
                              ordered = TRUE)

  # Outcomes
  df_plot$var_label <- "Number of infections"

  # Colors
  v_colors <- c("#56B4E9","#009E73")
  names(v_colors) <- c("Transmission rate",
```

```
                       "Recovery rate")

  # Line type
  v_linetype <- c(1, 3)
  names(v_linetype) <- c("Transmission rate",
                         "Recovery rate")

  # Plot
  gg_Sfun <- ggplot(df_plot,
                    aes(x = x, y = value, color = Parameter,
                        linetype = Parameter)) +
    geom_line(linewidth = 1.2) +
    scale_color_manual("", values = v_colors) +
    scale_linetype_manual("", values = v_linetype) +
    scale_y_continuous(breaks = number_ticks(6)) +
    scale_x_continuous(breaks = number_ticks(6)) +
    ylab("Sensitivity") + xlab("Time (days)") +
    theme_bw(base_size = 18) +
    theme(legend.position = "inside",
          legend.position.inside = c(0.78,0.85),
          legend.direction = "horizontal",
          strip.background = element_rect(color = "white",
                                          fill = "white"),
          axis.text.x = element_text(angle = 0,
                                     vjust = 0.5,
                                     hjust = 0.5))

  return(gg_Sfun)
}

#' Plot collinearity index
#'
#' \code{plot_CollIndx} plots collinearity index
#'
#' @param df_collin Data.frame containing collinearity indices from \code{collin}.
#' @param flag_each Flag (default is FALSE) to whether individual outcome
#' combinations is plotted.
#' @param log_y Flag (default is TRUE) to whether transform y-axis to log.

#' @return A ggplot2 object.
#'
#' @export
plot_CollIndx <- function(df_collin, flag_each = FALSE, log_y = TRUE){

  # Run coll
  collin_max <- max(df_collin$collinearity)

  # y-axis upper limit
  if(collin_max > 15){
    y_end <- collin_max + 3
  }else{
    y_end <- 18
  }

  # Vars
  v_vars <- unique(df_collin$var)
  n_vars <- length(v_vars)

  # x-axis discrete values
  v_x_breaks <- unique(df_collin$N)

  if(log_y){
```

```
  if(flag_each){
    plot_collindx <- ggplot(subset(df_collin, var != "All"),
                            aes(x = factor(N), y = collinearity)) +
      geom_point() +
      theme_bw() +
      facet_wrap(~Outcome, ncol = 3) +
      scale_y_log10(n.breaks = 6) +
      scale_x_discrete(name = "Number of parameters",
                       breaks = paste0(v_x_breaks),
                       labels  = paste0(v_x_breaks)) +
      ylab("Collinearity index") +
      geom_hline(yintercept = 15,
                 col = "red",
                 linetype = "dashed") +
      theme_bw(base_size = 18) +
      theme(legend.position = "bottom",
            strip.background = element_rect(color = "white",
                                             fill = "white"),
            strip.text = element_text(size = 11),
            axis.text.x = element_text(angle = 0,
                                       vjust = 0.5,
                                       hjust = 0.5),
            panel.grid.minor = element_line(color = "white"))  +
      coord_cartesian(ylim = c(NA, y_end))

    plot_collindx_all <- ggplot(subset(df_collin, var == "All"),
                                aes(x = factor(N), y = collinearity)) +
      geom_point() +
      theme_bw() +
      facet_wrap(~Outcome, ncol = 1) +
      scale_y_log10(n.breaks = 6) +
      scale_x_discrete(name = "Number of parameters",
                       breaks = df_collin$N,
                       labels  = df_collin$N) +
      ylab("Collinearity index") +
      geom_hline(yintercept = 15,
                 col = "red",
                 linetype = "dashed") +
      theme_bw(base_size = 18) +
      theme(legend.position = "bottom",
            strip.background = element_rect(color = "white",
                                             fill = "white"),
            strip.text = element_text(size = 11),
            axis.text.x = element_text(angle = 0,
                                       vjust = 0.5,
                                       hjust = 0.5),
            panel.grid.minor = element_line(color = "white"))  +
      coord_cartesian(ylim = c(NA, y_end))
  }else{

    plot_collindx_all <- ggplot(subset(df_collin, var == "All"),
                                aes(x = factor(N), y = collinearity)) +
      geom_point() +
      theme_bw() +
      facet_wrap(~Outcome, ncol = 1) +
      scale_y_log10(n.breaks = 6) +
      scale_x_discrete(name = "Number of parameters",
                       breaks = df_collin$N,
                       labels  = df_collin$N) +
      ylab("Collinearity index") +
      geom_hline(yintercept = 15,
                 col = "red",
```

```r
                  linetype = "dashed") +
      theme_bw(base_size = 18) +
      theme(legend.position = "bottom",
            strip.background = element_rect(color = "white",
                                            fill = "white"),
            strip.text = element_text(size = 11),
            axis.text.x = element_text(angle = 0,
                                       vjust = 0.5,
                                       hjust = 0.5),
            panel.grid.minor = element_line(color = "white"))  +
      coord_cartesian(ylim = c(NA, y_end))

  }

}else{

  if(flag_each){
    plot_collindx <- ggplot(subset(df_collin, var != "All"),
                           aes(x = factor(N), y = collinearity)) +
      geom_point() +
      theme_bw() +
      facet_wrap(~Outcome, ncol = 3) +
      scale_y_continuous(n.breaks = 7) +
      scale_x_discrete(name = "Number of parameters",
                       breaks = paste0(v_x_breaks),
                       labels  = paste0(v_x_breaks)) +
      ylab("Collinearity index") +
      xlab("Number of parameters") +
      geom_hline(yintercept = 15,
                 col = "red",
                 linetype = "dashed") +
      theme_bw(base_size = 18) +
      theme(legend.position = "bottom",
            strip.background = element_rect(color = "white",
                                            fill = "white"),
            strip.text = element_text(size = 11),
            axis.text.x = element_text(angle = 0,
                                       vjust = 0.5,
                                       hjust = 0.5),
            panel.grid.minor = element_line(color = "white"))  +
      coord_cartesian(ylim = c(NA, y_end))

    plot_collindx_all <- ggplot(subset(df_collin, var == "All"),
                               aes(x = factor(N), y = collinearity)) +
      geom_point() +
      theme_bw() +
      facet_wrap(~Outcome, ncol = 3) +
      scale_y_continuous(n.breaks = 7) +
      scale_x_discrete(name = "Number of parameters",
                       breaks = paste0(v_x_breaks),
                       labels  = paste0(v_x_breaks)) +
      ylab("Collinearity index") +
      xlab("Number of parameters") +
      geom_hline(yintercept = 15,
                 col = "red",
                 linetype = "dashed") +
      theme_bw(base_size = 18) +
      theme(legend.position = "bottom",
            strip.background = element_rect(color = "white",
                                            fill = "white"),
            strip.text = element_text(size = 11),
            axis.text.x = element_text(angle = 0,
```

```
                                      vjust = 0.5,
                                      hjust = 0.5),
            panel.grid.minor = element_line(color = "white"))  +
      coord_cartesian(ylim = c(NA, y_end))
  }else{

    plot_collindx_all <- ggplot(subset(df_collin, var == "All"),
                                aes(x = factor(N), y = collinearity)) +
      geom_point() +
      theme_bw() +
      facet_wrap(~Outcome, ncol = 3) +
      scale_y_continuous(n.breaks = 7) +
      scale_x_discrete(name = "Number of parameters",
                       breaks = paste0(v_x_breaks),
                       labels  = paste0(v_x_breaks)) +
      ylab("Collinearity index") +
      xlab("Number of parameters") +
      geom_hline(yintercept = 15,
                 col = "red",
                 linetype = "dashed") +
      theme_bw(base_size = 18) +
      theme(legend.position = "bottom",
            strip.background = element_rect(color = "white",
                                            fill = "white"),
            strip.text = element_text(size = 11),
            axis.text.x = element_text(angle = 0,
                                      vjust = 0.5,
                                      hjust = 0.5),
            panel.grid.minor = element_line(color = "white"))  +
      coord_cartesian(ylim = c(NA, y_end))

  }

}

if(flag_each){

  # Gather plots
  gg_collindx <- ggarrange(plot_collindx, NULL,
                           plot_collindx_all,
                           heights = c(1,0.05,1),
                           widths = 1,
                           ncol = 1, nrow = 3)

}else{
  gg_collindx <- plot_collindx_all
}

return(gg_collindx)
}

#' Plot global sensitivity analysis
#'
#' \code{plot_GSA} plots Sobol' first-order and total sensitivity indices.
#'
#' @param df_sobol Data.frame containing the results from a GSA using function
#' \code{run_GSA}.
#'
#' @returns A ggplot object.
#'
#' @export
plot_GSA <- function(df_sobol){
```

```r
  # Label variables
  df_sobol$par <- factor(df_sobol$par,
                         levels = c("r_beta","r_gamma"),
                         labels = c("Transmission rate","Recovery rate"))

  # Plot
  ggGSA <- ggplot(df_sobol,
         aes(x = x, y = original, ymin = min..c.i., ymax = max..c.i.,
             color = type, shape = type)) +
    geom_point(position = position_dodge(width = 0.5), size = 3) +
    geom_errorbar(position = position_dodge(width = 0.5),
                  linewidth = 1.05,
                  width = 0.25) +
    ylab("Sobol index") + xlab("Time (days)") +
    scale_x_continuous(breaks = number_ticks(6)) +
    scale_y_continuous(breaks = seq(0,1,0.25)) +
    scale_color_manual("", values = c("Main effect" = "#CD0BBC",
                                      "Total effect" = "#999999")) +
    scale_shape_manual("", values = c("Main effect" = 1,
                                      "Total effect" = 2)) +
    facet_wrap(~par, ncol = 1) +
    theme_bw(base_size = 18) +
    theme(legend.position = "inside",
          legend.position.inside = c(0.83,0.95),
          legend.direction = "horizontal",
          strip.background = element_rect(color = "white",
                                          fill = "white"),
          axis.text.x = element_text(angle = 0,
                                     vjust = 0.5,
                                     hjust = 0.5)) +
    coord_cartesian(ylim = c(0,1))

  return(ggGSA)

}

# Calibration functions ---------------------------------------------------
#' Generate model outputs for calibration from a parameter set
#'
#' \code{SIR_calibration_out} computes model outputs for the SIR transmission
#' model to be used for calibration routines.
#'
#' @param v_params Vector (or matrix) containing the model parameters.
#' @param l_params_all List containing all the parameters of the decision model.
#'
#' @return A data.frame containing the model-predicted outcomes.
#'
#' @export
SIR_calibration_out <- function(v_params, l_params_all){

  # Update list of parameters with calibrated parameter set
  l_params_all$r_beta  <- v_params["r_beta"]
  l_params_all$r_gamma <- v_params["r_gamma"]

  # Run model
  l_out_model  <- SIR_model(l_params_all)

  # Calculate total infections
  df_DXIncTot <- calc_DXIncTot(l_out_model  = l_out_model,
                               l_params_all = l_params_all)

  # Return data.frame
```

```r
  return(list(DXIncTot = df_DXIncTot))
}

#' Sample from prior distributions of calibrated parameters
#'
#' \code{sample_prior} generates a sample of calibrated parameters
#' from their prior distribution.
#'
#' @param n_samp Number of samples.
#' @param v_lb Vector containing the lower bounds of the calibrated
#' parameters.
#' @param v_ub Vector containing the upper bounds of the calibrated
#' parameters.
#'
#' @return A matrix with n number of columns as number of parameters to
#' be calibrated and \code{n_samp} rows. Each row corresponds to a
#' parameter set sampled from their prior distributions.
#'
#' @export
sample_prior <- function(n_samp, v_lb, v_ub){

  # Number of parameters
  n_param <- length(v_lb)

  # Create random Latin hypercube design
  m_lhs_unit   <- randomLHS(n = n_samp, k = n_param)

  # Sample
  m_param_samp <- matrix(nrow = n_samp, ncol = n_param)
  colnames(m_param_samp) <- v_param_names
  for (i in 1:n_param){
    m_param_samp[, i] <- qunif(m_lhs_unit[,i],
                               min = v_lb[i],
                               max = v_ub[i])
    # ALTERNATIVE prior using beta (or other) distributions
    # m_param_samp[, i] <- qbeta(m_lhs_unit[,i],
    #                            shape1 = 1,
    #                            shape2 = 1)
  }
  return(m_param_samp)
}

#' Evaluate log-prior of calibrated parameters
#'
#' \code{log_prior} computes a log-prior value for one (or multiple) parameter
#' set(s) based on their prior distributions.
#'
#' @param v_params Vector (or matrix) containing the model parameters.
#' @param v_lb Vector containing the lower bounds for each parameter.
#' @param v_ub Vector containing the upper bounds for each parameter.
#'
#' @return A scalar (or vector) containing the log-prior values.
#'
#' @export
log_prior <- function(v_params, v_lb, v_ub){

  # Get param names
  v_param_names <- names(v_lb)

  if(is.null(dim(v_params))) { # If vector, change to matrix
    v_params <- t(v_params)
  }
```

```r
  n_samp <- nrow(v_params)
  colnames(v_params) <- v_param_names
  lprior <- rep(0, n_samp)
  for (i in 1:n_param){
    lprior <- lprior + dunif(v_params[, i],
                             min = v_lb[i],
                             max = v_ub[i],
                             log = T)
    # ALTERNATIVE prior using beta distributions
    # lprior <- lprior + dbeta(v_params[, i],
    #                          shape1 = 1,
    #                          shape2 = 1,
    #                          log = T)
  }
  return(lprior)
}

#' Evaluate prior of calibrated parameters
#'
#' \code{prior} computes a prior value for one (or multiple) parameter set(s).
#'
#' @param v_params Vector (or matrix) containing the model parameters.
#' @param v_lb Vector containing the lower bounds for each parameter.
#' @param v_ub Vector containing the upper bounds for each parameter.
#'
#' @return A scalar (or vector) containing the prior values.
#'
#' @export
prior <- function(v_params, v_lb, v_ub) {
  return(exp(log_prior(v_params, v_lb, v_ub)))
}

#' Log likelihood normal distribution
#'
#' \code{log_lik} computes a log-likelihood value for one (or multiple)
#' parameter set(s).
#'
#' @param v_params_calib Vector (or matrix) containing the calibrated parameters.
#' @param v_dates Vector containing the dates of the available data to perform
#' calibration.
#' @param ... Further arguments to be passed to.
#'
#' @return A scalar (or vector) containing the log-likelihood values.
#'
#' @export
log_lik <- function(v_params, v_dates, ...){

  # par_vector: a vector (or matrix) of model parameters
  if(is.null(dim(v_params))) { # If vector, change to matrix
    v_params <- t(v_params)
  }

  # Set variables
  n_samp         <- nrow(v_params)
  v_target_names <- c("DXIncTot")
  n_targets      <- length(v_target_names)
  v_llik         <- matrix(0, nrow = n_samp, ncol = n_targets)
  v_llik_overall <- numeric(n_samp)

  for(j in 1:n_samp) { # j=1
    jj <- tryCatch( {
      ###   Run model for parameter set "v_params" ###
```

```r
      l_model_out <- SIR_calibration_out(v_params[j, ],
                                         # l_params_all = l_params_all)
                                         ...)
      ## Model output
      df_model_DXIncTot <- l_model_out$DXIncTot %>%
        filter(Date %in% v_dates)

      ## Targets
      df_targets_DXIncTot <- df_targets %>%
        filter(abbrev_outcome == "DXIncTot" &
                 Date %in% v_dates)

      ###  Calculate log-likelihood of model outputs to targets  ###
      # TARGET 1: Number of infections (DXIncTot)
      ## Negative-Binomial log-likelihood
      v_llik[j, 1] <- sum(dnbinom(x = df_targets_DXIncTot$value,
                                  size = 5,#1,
                                  mu = df_model_DXIncTot$value,
                                  log = T), na.rm = T)
      # ## Poisson log-likelihood
      # v_llik[j, 1] <- sum(dpois(x = df_targets_DXIncTot$value,
      #                           lambda = df_model_DXIncTot$value,
      #                           log = T), na.rm = T)

      ## OVERALL ADD TO SIR
      ## can give different targets different weights (user must change this)
      v_weights <- rep(1, n_targets)
      ## weighted sum
      v_llik_overall[j] <- v_llik[j, ] %*% v_weights

    }, error = function(e) NA)
    if (is.na(jj)) { v_llik_overall[j] <- -Inf }
  } # End loop over sampled parameter sets

  # return LLIK
  return(v_llik_overall)
}

#' Likelihood
#'
#' \code{likelihood} computes a likelihood value for one (or multiple)
#' parameter set(s).
#'
#' @param v_params_calib Vector (or matrix) containing the model parameters.
#' @param v_dates Vector containing the dates of the available data
#' to perform calibration.
#' @param ... Further arguments to be passed to.
#'
#' @return A scalar (or vector) containing the likelihood values.
#'
#' @export
likelihood <- function(v_params, v_dates, ...){
  return(exp(log_lik(v_params, v_dates, ...)))
}

#' Evaluate log-posterior of calibrated parameters
#'
#' \code{log_post} computes a log-posterior value for one (or multiple)
#' parameter set(s) based on the simulation model, likelihood functions and
#' prior distributions.
#'
#' @param v_params_calib Vector (or matrix) containing the model parameters.
```

```r
#' @param v_lb Vector containing the lower bounds for each parameter.
#' @param v_ub Vector containing the upper bounds for each parameter.
#' @param v_dates Vector containing the dates of the available data
#' to perform calibration.
#' @param ... Further arguments to be passed to.
#'
#' @return A scalar (or vector) with log-posterior values.
#'
#' @export
log_post <- function(v_params, v_lb, v_ub, v_dates, ...) {
  lpost <- log_prior(v_params, v_lb, v_ub) + log_lik(v_params, v_dates, ...)
  return(lpost)
}

#' Evaluate posterior of calibrated parameters
#'
#' \code{posterior} computes a posterior value for one (or multiple)
#' parameter set(s).
#'
#' @param v_params_calib Vector (or matrix) containing the model parameters.
#' @param v_lb Vector containing the lower bounds for each parameter.
#' @param v_ub Vector containing the upper bounds for each parameter.
#' @param ... Further arguments to be passed to.
#'
#' @return A scalar (or vector) containing the posterior values.
#'
#' @export
posterior <- function(v_params, v_lb, v_ub, ...) {
  exp(log_post(v_params, v_lb, v_ub, ...))
}
```

**SIR_IMIS_function.R**

```r
#' Incremental Mixture Importance Sampling (IMIS package) adapted for the
#' SIR model
#'
#' @param B The incremental sample size at each iteration of IMIS.
#' @param B.re The desired posterior sample size at the resample stage.
#' @param number_k The maximum number of iterations in IMIS
#' @param D The number of optimizers which could be 0.
#' @param v_lb Vector containing the lower bounds of the calibrated
#' parameters.
#' @param v_ub Vector containing the upper bounds of the calibrated
#' parameters.
#' @param l_params_all List with all parameters of decision model.
#' @param v_dates Vector containing the dates of the available data
#' to perfom calibration.
#'
#' @return The posterior resamples.
#' @source Adrain Raftery and Le Bao. IMIS package.
#' http://cran.nexr.com/web/packages/IMIS/index.html
#'
#' @import mvtnorm
#' @export
IMIS <- function(B=1000, B.re=3000, number_k=100, D=0,
                 v_lb, v_ub, l_params_all, v_dates){
  B0 = B*10
  # Draw initial samples from the prior distribution
  X_all = X_k = sample_prior(B0, v_lb = v_lb, v_ub = v_ub)
  if (is.vector(X_all))        Sig2_global = var(X_all)        # the prior covariance
```

```
if (is.matrix(X_all))        Sig2_global = cov(X_all)        # the prior covariance

# 6 diagnostic statistics at each iteration
stat_all = matrix(NA, 6, number_k)

# centers of Gaussian components, prior densities, and likelihoods
center_all = prior_all = like_all = NULL

# covariance matrices of Gaussian components
sigma_all = list()
if (D>=1)        option.opt = 1            # use optimizer
if (D==0) option.opt = 0; D=1         # NOT use optimizer

for (k in 1:number_k){ # k = 1

  ptm.like = proc.time()
  prior_all = c(prior_all,
                prior(v_params = X_k,
                      v_lb = v_lb,
                      v_ub = v_ub))                  # Calculate the prior densities
  like_all = c(like_all,
               likelihood(X_k,
                          l_params_all = l_params_all,
                          v_dates = v_dates))                # Calculate the likelihoods
  ptm.use = (proc.time() - ptm.like)[3]
  if (k==1){
    print(paste(B0, "likelihoods are evaluated in",
                round(ptm.use/60,2), "minutes"))
  }

  if (k==1)        envelop_all = prior_all        # envelop stores the sampling densities
  if (k>1){
    envelop_all = apply(rbind(prior_all*B0/B, gaussian_all), 2, sum)/(B0/B+D+(k-2))
  }

  # importance weight is determined by the posterior density divided
  # by the sampling density
  Weights = prior_all*like_all / envelop_all
  stat_all[1,k] = log(mean(Weights))                          # the raw marginal likelihood
  Weights = Weights / sum(Weights)
  stat_all[2,k] = sum(1-(1-Weights)^B.re)         # the expected number of unique points
  stat_all[3,k] = max(Weights)                                  # the maximum weight
  stat_all[4,k] = 1/sum(Weights^2)                            # the effective sample size
  # the entropy relative to uniform
  stat_all[5,k] = -sum(Weights*log(Weights), na.rm = TRUE) / log(length(Weights))
  stat_all[6,k] = var(Weights/mean(Weights))         # the variance of scaled weights
  if (k==1)        print("Stage   MargLike   UniquePoint   MaxWeight   ESS")
  print(c(k, round(stat_all[1:4,k], 3)))

  if (k==1 & option.opt==1){
    if (is.matrix(X_all))        Sig2_global = cov(X_all[which(like_all>min(like_all)),])

    # exclude the neighborhood of the local optima
    X_k = which_exclude = NULL

    label_weight = sort(Weights, decreasing = TRUE, index=TRUE)

    # the candidate inputs for the starting points
    which_remain = which(Weights>label_weight$x[B0])
    size_remain = length(which_remain)
    for (i in 1:D){
      important = NULL
```

```r
    if (length(which_remain)>0)
      important = which_remain[which(Weights[which_remain]==max(Weights[which_remain]))]
    if (length(important)>1)        important = sample(important,1)
    if (is.vector(X_all))        X_imp = X_all[important]
    if (is.matrix(X_all))        X_imp = X_all[important,]
    # Remove the selected input from candidates
    which_exclude = union( which_exclude, important )
    which_remain = setdiff(which_remain, which_exclude)
    posterior = function(theta){        -log(prior(theta, v_lb = v_lb, v_ub = v_ub))-
        log(likelihood(theta, l_params_all = l_params_all, v_dates = v_dates)) }

    if (is.vector(X_all)){
      if (length(important)==0)        X_imp = center_all[1]
      optimizer = optim(X_imp, posterior, method="BFGS", hessian=TRUE,
                        control=list(parscale=sqrt(Sig2_global)/10,maxit=5000))
      print(paste("maximum posterior=", round(-optimizer$value,2),
                  ", likelihood=",
                  round(log(likelihood(optimizer$par,
                                       l_params_all = l_params_all,
                                       v_dates = v_dates)),2),
                  ", prior=",
                  round(log(prior(optimizer$par,
                                  v_lb = v_lb,
                                  v_ub = v_ub)),2),
                  ", time used=", round(ptm.use/60,2),
                  "minutes, convergence=", optimizer$convergence))
      center_all = c(center_all, optimizer$par)
      sigma_all[[i]] = solve(optimizer$hessian)
      # Draw new samples:
      X_k = c(X_k, rnorm(B, optimizer$par, sqrt(sigma_all[[i]])) )
      distance_remain = abs(X_all[which_remain]-optimizer$par)
    }
    if (is.matrix(X_all)){
      # The rough optimizer uses the Nelder-Mead algorithm.
      if (length(important)==0)        X_imp = center_all[1,]
      ptm.opt = proc.time()
      optimizer = optim(X_imp, posterior, method="Nelder-Mead",
                        control=list(maxit=1000, parscale=sqrt(diag(Sig2_global))) )
      theta.NM = optimizer$par

      # The more efficient optimizer uses the BFGS algorithm
      optimizer = optim(theta.NM, posterior, method="BFGS", hessian=TRUE,
                        control=list(parscale=sqrt(diag(Sig2_global)), maxit=1000))
      ptm.use = (proc.time() - ptm.opt)[3]
      print(paste("maximum posterior=", round(-optimizer$value,2),
                  ", likelihood=",
                  round(log(likelihood(optimizer$par,
                                       l_params_all = l_params_all,
                                       v_dates = v_dates)),2),
                  ", prior=",
                  round(log(prior(optimizer$par,
                                  v_lb = v_lb,
                                  v_ub = v_ub)),2),
                  ", time used=", round(ptm.use/60,2),
                  "minutes, convergence=", optimizer$convergence))
      center_all = rbind(center_all, optimizer$par)                # the center of new samples
      if (min(eigen(optimizer$hessian)$values)>0){
        # the covariance of new samples
        sigma_all[[i]] = solve(optimizer$hessian)
      }

      # If the hessian matrix is not positive definite, we define the covariance as following
```

```
        if (min(eigen(optimizer$hessian)$values)<=0){
          eigen.values = eigen(optimizer$hessian)$values
          eigen.values[which(eigen.values<0)] = 0
          hessian = eigen(optimizer$hessian)$vectors %*% diag(eigen.values) %*%
            t(eigen(optimizer$hessian)$vectors)
          sigma_all[[i]] = solve(hessian + diag(1/diag(Sig2_global)) )
        }
        # Draw new samples
        X_k = rbind(X_k, rmvnorm(B, optimizer$par, sigma_all[[i]]) )
        distance_remain = mahalanobis(X_all[which_remain,],
                                       optimizer$par,
                                       diag(diag(Sig2_global)) )
      }
      # exclude the neighborhood of the local optima
      label_dist = sort(distance_remain, decreasing = FALSE, index=TRUE)
      which_exclude = union(which_exclude,
                            which_remain[label_dist$ix[1:floor(size_remain/D)]])
      which_remain = setdiff(which_remain, which_exclude)
    }
    if (is.matrix(X_all))        X_all = rbind(X_all, X_k)
    if (is.vector(X_all))        X_all = c(X_all, X_k)

    saveRDS(Sig2_global, file = paste0(path,"Sig2_global.rds"))
  }

  if (k>1 | option.opt==0){
    important = which(Weights == max(Weights))
    if (length(important)>1)        important = important[1]

    # X_imp is the maximum weight input
    if (is.matrix(X_all))        X_imp = X_all[important,]

    if (is.vector(X_all))        X_imp = X_all[important]
    if (is.matrix(X_all))        center_all = rbind(center_all, X_imp)
    if (is.vector(X_all))        center_all = c(center_all, X_imp)
    if (is.matrix(X_all)){
      distance_all = mahalanobis(X_all, X_imp, diag(diag(Sig2_global)))
    }

    # Calculate the distances to X_imp
    if (is.vector(X_all))        distance_all = abs(X_all-X_imp)
    # Sort the distances
    label_nr = sort(distance_all, decreasing = FALSE, index=TRUE)
    which_var = label_nr$ix[1:B]        # Pick B inputs for covariance calculation
    if (is.matrix(X_all)){
      Sig2 = cov.wt(X_all[which_var,],
                    wt = Weights[which_var]+1/length(Weights),
                    cor = FALSE, center = X_imp, method = "unbias")$cov
    }
    if (is.vector(X_all)){
      Weights_var = Weights[which_var]+1/length(X_all)
      Weights_var = Weights_var/sum(Weights_var)
      Sig2 = (X_all[which_var]-X_imp)^2 %*% Weights_var
    }
    sigma_all[[D+k-1]] = Sig2
    if (is.matrix(X_all)){
      X_k = rmvnorm(B, X_imp, Sig2) # Draw new samples
    }
    if (is.vector(X_all)){
      X_k = rnorm(B, X_imp, sqrt(Sig2)) # Draw new samples
    }
    if (is.matrix(X_all))        X_all = rbind(X_all, X_k)
```

```
      if (is.vector(X_all))        X_all = c(X_all, X_k)
    }

    if (k==1){
      gaussian_all = matrix(NA, D, B0+D*B)
      for (i in 1:D){
        if (is.matrix(X_all)){
          gaussian_all[i,] = dmvnorm(X_all, center_all[i,], sigma_all[[i]])
        }
        if (is.vector(X_all)){
          gaussian_all[i,] = dnorm(X_all, center_all[i], sqrt(sigma_all[[i]]))
        }
      }
    }
    if (k>1){
      if (is.vector(X_all))        gaussian_new = matrix(0, D+k-1, length(X_all) )
      if (is.matrix(X_all))        gaussian_new = matrix(0, D+k-1, dim(X_all)[1] )
      if (is.matrix(X_all)){
        gaussian_new[1:(D+k-2), 1:(dim(X_all)[1]-B)] = gaussian_all
        gaussian_new[D+k-1, ] = dmvnorm(X_all, X_imp, sigma_all[[D+k-1]])
        for (j in 1:(D+k-2)){
          gaussian_new[j, (dim(X_all)[1]-B+1):dim(X_all)[1] ] =
            dmvnorm(X_k, center_all[j,], sigma_all[[j]])
        }
      }
      if (is.vector(X_all)){
        gaussian_new[1:(D+k-2), 1:(length(X_all)-B)] = gaussian_all
        gaussian_new[D+k-1, ] = dnorm(X_all, X_imp, sqrt(sigma_all[[D+k-1]]))
        for (j in 1:(D+k-2)){
          gaussian_new[j, (length(X_all)-B+1):length(X_all) ] =
            dnorm(X_k, center_all[j], sqrt(sigma_all[[j]]))
        }
      }
      gaussian_all = gaussian_new
    }
    if (stat_all[2,k] > (1-exp(-1))*B.re)        break
  } # end of k

  nonzero = which(Weights>0)
  which_X = sample(nonzero, B.re, replace = TRUE, prob = Weights[nonzero])
  if (is.matrix(X_all))        resample_X = X_all[which_X,]
  if (is.vector(X_all))        resample_X = X_all[which_X]

  return(list(stat      = t(stat_all),
              resample = resample_X,
              center   = center_all,
              Weights  = Weights[nonzero]))
} # end of IMIS
```

**SIR_analysis.R**

```
#-------------------------------------------------------------------------------#
# This script illustrates the implementation of PRE-CISE in a deterministic    #
# Susceptible-Infectious-Recovered (SIR) transmission model calibrated to      #
# daily incident cases using the IMIS algorithm.                               #
#-------------------------------------------------------------------------------#

rm(list = ls()) # clean environment

# Calibration specifications ----------------------------------------------
```

```r
# Model: Susceptible-Infected-Recovered (SIR) model without demography
# Inputs to be calibrated:
#   r_beta   - transmission rate
#   r_gamma  - recovery rate
# Targets:
#   DXIncTot - total incident cases
# Search method: Random search using Latin-Hypercube Sampling
# Goodness-of-fit measure: Sum of Log-Likelihood

# Load libraries and functions --------------------------------------------
# calibration functionality
library(lhs)
library(matrixStats) # package used for summary statistics
library(FME)         # For identifiability and collinearity analysis
library(tidyverse)
options(dplyr.summarise.inform = FALSE) # do not show summarise info

# visualization
library(psych)
library(ggplot2)
library(ggthemes)
library(ggpubr)

# global sensitivity
library(sensitivity)

# calibration
library(mvtnorm)
source("R/IMIS_function_SIR.R") # from IMIS package, adapted for this analysis
# Old version
# devtools::install_version("IMIS", version = "0.1", repos = "http://cran.us.r-project.org")

# load functions
source("R/helper_functions.R") # from dampack package
source("R/SIR_functions.R")

# Target data -------------------------------------------------------------
# Load target data: influenza outbreak at a British boarding school in 1978
# (Anon 1978; El Attouga & El Khalifi, 2024 ; Keeling & Rohani, 2008)
# Source:
# https://www.sciencedirect.com/science/article/pii/S0960077924005939#appendix
# https://www.bmj.com/content/bmj/1/6112/586.full.pdf
df_InfluenzaData_raw <- readxl::read_xlsx("data/Influenza1978Data.xlsx")
df_InfluenzaData_raw$Date <- as.Date(df_InfluenzaData_raw$Date)

df_targets <- df_InfluenzaData_raw %>%
  mutate(Type = "Target",
         abbrev_outcome = "DXIncTot",
         Day = as.numeric(Date - Date[1] + 1),
         Date0 = Date - Date[1],
         lb = epitools::pois.exact(x = value, pt = population)$lower*population,
         ub = epitools::pois.exact(x = value, pt = population)$upper*population,
         se = (ub-lb)/(1.96*2)) %>%
  select(Type,abbrev_outcome,Outcome,population,Day,Date,Date0,value,lb,ub,se) %>%
  as.data.frame()

# Plot the targets
plot_targets(df_targets)

# Variables ---------------------------------------------------------------
# Calibrated dates
v_dates <- df_targets$Date
```

```r
# First day of calibration
n_date_init <- first(v_dates)

# Last day of calibration
n_date_end <- last(v_dates)

# Number of days until infection become extinguished
n_t <- as.numeric(n_date_end - n_date_init)

# Number of model evaluations per day
time_step <- 1

# Total number of the boys in school
n_tot_pop <- unique(df_targets$population)

# Initial number of infected boys
n_inf_init <- df_targets$value[df_targets$Date == n_date_init]

# Number of simulations (coverage analysis)
n_sim <- 300

# Seed number
n_seed <- 111124

# Set seed
set.seed(n_seed)

# Number of calibration targets
v_outcome_names <- c("Number of infections")
names(v_outcome_names) <- c("DXIncTot")
n_outcomes <- length(v_outcome_names)

# Specify calibration parameters ------------------------------------------
# Names and number of input parameters to be calibrated
v_param_names <- c("r_beta",    # transmission rate
                   "r_gamma")  # recovery rate
n_param <- length(v_param_names)

# Range on input search space
v_lb <- c(r_beta = 0.50, r_gamma = 0.10)  # lower bound
v_ub <- c(r_beta = 3.00, r_gamma = 1.00)  # upper bound

# Model -------------------------------------------------------------------
# -inputs are parameters to be estimated through calibration
# -outputs correspond to the target data

# Set of parameter test
v_params_test <- c(r_beta = 1.66, r_gamma = 1/2.2)

# List of parameters
l_params_all <- load_params_all(n_t         = n_t,
                                time_step   = time_step,
                                n_date_init = n_date_init,
                                n_tot_pop   = n_tot_pop,
                                n_inf_init  = n_inf_init,
                                r_beta      = v_params_test["r_beta"],
                                r_gamma     = v_params_test["r_gamma"])

# Run model, check it works
l_out_model <- SIR_model(l_params_all = l_params_all)
l_out_model$df_out_model
```

```r
# Calibration functions (check that it works) -----------------------------
# View resulting parameter set samples
pairs.panels(sample_prior(n_samp = 1000, v_lb = v_lb, v_ub = v_ub))

# Prior
log_prior(v_params = v_params_test,
          v_lb     = v_lb,
          v_ub     = v_ub)
log_prior(v_params = sample_prior(10, v_lb = v_lb, v_ub = v_ub),
          v_lb     = v_lb,
          v_ub     = v_ub)

prior(v_params = v_params_test,
      v_lb     = v_lb,
      v_ub     = v_ub)
prior(v_params = sample_prior(10, v_lb = v_lb, v_ub = v_ub),
      v_lb     = v_lb,
      v_ub     = v_ub)

# Likelihood
log_lik(v_params     = v_params_test,
        l_params_all = l_params_all,
        v_dates      = v_dates)
log_lik(v_params     = sample_prior(10, v_lb = v_lb, v_ub = v_ub),
        l_params_all = l_params_all,
        v_dates      = v_dates)

likelihood(v_params     = v_params_test,
           l_params_all = l_params_all,
           v_dates      = v_dates)
likelihood(v_params     = sample_prior(10, v_lb = v_lb, v_ub = v_ub),
           l_params_all = l_params_all,
           v_dates      = v_dates)

# Posterior
log_post(v_params     = v_params_test,
         v_lb         = v_lb,
         v_ub         = v_ub,
         l_params_all = l_params_all,
         v_dates      = v_dates)
log_post(v_params     = sample_prior(10, v_lb = v_lb, v_ub = v_ub),
         v_lb         = v_lb,
         v_ub         = v_ub,
         l_params_all = l_params_all,
         v_dates      = v_dates)

posterior(v_params     = v_params_test,
          v_lb         = v_lb,
          v_ub         = v_ub,
          l_params_all = l_params_all,
          v_dates      = v_dates)
posterior(v_params     = sample_prior(10, v_lb = v_lb, v_ub = v_ub),
          v_lb         = v_lb,
          v_ub         = v_ub,
          l_params_all = l_params_all,
          v_dates      = v_dates)

# Define functions -------------------------------------------------------
## Coverage analysis -----------------------------------------------------
# Coverage function
# l_params_all: List containing all the parameters of decision model.
# m_params: Matrix containing sample of parameter sets
```

```
run_coverage <- function(l_params_all, m_params){

  # Number of simulations
  n_sim <- nrow(m_params)

  # Run model
  df_model <- data.frame(NULL) # empty data.frame to store results
  for(iter_i in 1:n_sim){ # iter_i = 1

    # Update list of parameters
    l_params_all$r_beta  <- m_params[iter_i, "r_beta"]
    l_params_all$r_gamma <- m_params[iter_i, "r_gamma"]

    # Run model
    l_out_model <- SIR_model(l_params_all = l_params_all)

    # Output of the model in a data.frame
    df_temp <- calc_DXIncTot(l_out_model  = l_out_model,
                             l_params_all = l_params_all)

    # Gather results
    df_model <- rbind(df_model, df_temp)

    # Print progress
    if(iter_i/100==round(iter_i/100,0)) {
      cat('\r',paste(round(iter_i/n_sim*100,0),"% simulations done",sep=""))
    }
  }

  # Summarise
  df_model_summ <- df_model %>%
    group_by(Type, Outcome, Day, Date, Date0) %>%
    summarise(mean_val = mean(value),
              lb       = quantile(value,probs = 0.025),
              ub       = quantile(value, probs = 0.975),
              se       = sd(value)) %>%
    ungroup() %>%
    rename(value = mean_val)

  return(list(df_model        = df_model,
              df_model_summ   = df_model_summ))
}

## Sensitivity analysis ---------------------------------------------------
# Cost function for one-at-a-time sensitivity analysis
# v_params:     Vector containing the model parameters.
# l_params_all: List containing all the parameters of decision model.
# v_dates:      Vector containing dates at which cost function is to be computed.
costFun <- function(v_params, l_params_all, v_dates) {

  # Update list of parameters
  l_params_all$r_beta  <- v_params["r_beta"]
  l_params_all$r_gamma <- v_params["r_gamma"]

  # Run model
  l_out_model <- SIR_model(l_params_all = l_params_all)

  # Output of the model in a data.frame
  df_DXIncTot <- calc_DXIncTot(l_out_model  = l_out_model,
                               l_params_all = l_params_all)

  # Filter and select columns
```

```
  ## Number of infections
  df_model_DXIncTot <- df_DXIncTot %>%
    filter(abbrev_outcome == "DXIncTot" & Date %in% v_dates) %>%
    mutate(time = Day) %>%
    rename(DXIncTot = value) %>%
    select(time, DXIncTot)

  df_targets_DXIncTot <- df_targets %>%
    filter(abbrev_outcome == "DXIncTot" & Date %in% v_dates) %>%
    mutate(time = Day) %>%
    select(time, value, se) %>%
    rename(DXIncTot = value)

  # Compute "costs" for each target
  cost <- modCost(model = df_model_DXIncTot,
                  obs   = df_targets_DXIncTot,
                  err   = "se")
  return(cost)
}

# Function to compute model outputs for global sensitivity analyses
# m_params:     Matrix containing sample of parameter sets.
# l_params_all: List containing all the parameters of decision model.
# n_Date:       Date at which GSA is to be computed.
model_out_GSA <- function(m_params, l_params_all, n_Date){

  v_res <- c()
  for(i in 1:nrow(m_params)){

    # Update list of parameters with calibrated parameter set
    l_params_all$r_beta  <- m_params[i,"r_beta"]
    l_params_all$r_gamma <- m_params[i,"r_gamma"]

    # Run model
    l_out_model  <- SIR_model(l_params_all)

    # Calculate total infections
    df_DXIncTot <- calc_DXIncTot(l_out_model  = l_out_model,
                                 l_params_all = l_params_all) %>%
      filter(Date == n_Date)

    # Bind results
    v_res <- c(v_res, df_DXIncTot$value)

  }

  # Return vector of values
  return(v_res)
}

# Global sensitivity analysis (GSA): this function runs soboljansen function
# from sensitivity package.
# l_params_all: List containing all the parameters of decision model.
# v_dates:      Vector containing dates at which cost function is to be computed.
# X1:           First random sample (prior distribution).
# X2:           Second random sample (prior distribution).
# n_boot:       Number of bootstrap replicates
# n_conf:       Confidence level for bootstrap confidence intervals.
run_GSA <- function(l_params_all, v_dates, X1, X2, n_boot, n_conf){

  df_res_sobol <- data.frame(NULL)
  for(i in 2:length(v_dates)){ # i = 2
```

```
    cat("Day", i, "\n")

    # Compute Sobol Indices
    res_sobol <- soboljansen(model          = model_out_GSA,
                             X1             = X1,
                             X2             = X2,
                             l_params_all = l_params_all,
                             n_Date         = v_dates[i],
                             nboot          = n_boot,
                             conf           = n_conf)

    # Bind results
    df_res_sobol <- rbind(df_res_sobol,
                          data.frame(par  = rownames(res_sobol$S),
                                     type = "Main effect",
                                     x    = i,
                                     res_sobol$S,
                                     row.names = NULL),
                          data.frame(par  = rownames(res_sobol$T),
                                     type = "Total effect",
                                     x    = i,
                                     res_sobol$T,
                                     row.names = NULL))

  }
  return(df_res_sobol)
}

# Compare sensitivity analysis --------------------------------------------
# Define input search space
v_lb_Sens <- c(r_beta = 0.30, r_gamma = 0.10)  # lower bound
v_ub_Sens <- c(r_beta = 3.50, r_gamma = 1.20)  # upper bound

## Global sensitivity analysis
# Get two sets of priors
set.seed(n_seed)    # set seed
m_params_X1 <- sample_prior(n_samp = n_sim,
                            v_lb   = v_lb_Sens,
                            v_ub   = v_ub_Sens)

m_params_X2 <- sample_prior(n_samp = n_sim,
                            v_lb   = v_lb_Sens,
                            v_ub   = v_ub_Sens)

# Run GSA
df_res_sobol <- run_GSA(l_params_all = l_params_all,
                        v_dates      = v_dates,
                        X1           = m_params_X1,
                        X2           = m_params_X2,
                        n_boot       = 100,
                        n_conf       = 0.95)

# Bootstrap confidence interval bounds were manually restricted to
# the plausible range [0,1]
df_res_sobol <- df_res_sobol %>%
  mutate(min..c.i. = ifelse(min..c.i.<0, 0, min..c.i.),
         max..c.i. = ifelse(max..c.i.>1, 1, max..c.i.))

# Plot Sobol' indices
plot_GSA(df_sobol = df_res_sobol)

## Local sensitivity
```

```r
# Compute model-data residuals on previous coverage (mean values)
v_params <- colMeans(m_params_X1)        # mean values

# Sensitivity analysis
SensRes <- sensFun(func         = costFun,
                   parms        = v_params,
                   l_params_all = l_params_all,
                   v_dates = v_dates)

plot_Sfun(SensRes)

# Collinearity analysis on accumulated targets ----------------------------
# Define input search space
v_lb_0 <- c(r_beta = 0.40, r_gamma = 0.10)  # lower bound
v_ub_0 <- c(r_beta = 3.00, r_gamma = 1.00)  # upper bound

# Run initial coverage analysis
set.seed(n_seed)  # set seed
m_params_1 <- sample_prior(n_samp = 1000,
                           v_lb   = v_lb_0,
                           v_ub   = v_ub_0)

# Dates index
v_dates_index  <- 1:(n_t+1)

# Empty data.frames to store results
df_collin_iter <- data.frame(NULL)
for(j in seq_along(v_dates_index)[-(n_t+1)]){ # j = 2

  # Select dates
  v_index_samp <- sort(v_dates_index[1:(1+j)])
  v_dates_samp <- v_dates[v_index_samp]

  # 1. Compute model-data residuals on previous coverage (mean values)
  v_params <- colMeans(m_params_1)
  modCost   <- costFun(v_params     = v_params,
                       l_params_all = l_params_all,
                       v_dates      = v_dates_samp)

  # 2. Local sensitivity analysis
  SensRes <- sensFun(func         = costFun,
                     parms        = v_params,
                     l_params_all = l_params_all,
                     v_dates      = v_dates_samp)

  m_Sens <- summary(SensRes)

  # 3. Collinearity analysis
  df_collin <- collin(SensRes) %>%
    mutate(n_targets = j+1,
           var     = "All",
           Outcome = paste0(v_outcome_names,collapse = " & ")) %>%
    relocate(n_targets, var)

  # Bind data.frames
  df_collin_iter <- rbind(df_collin_iter, df_collin)

}

# Implement PRE-CISE (wide) ----------------------------------------------
# 0. Define input search space and plot first coverage
v_lb_1 <- c(r_beta = 0.30, r_gamma = 0.10)  # lower bound
```

```r
v_ub_1 <- c(r_beta = 3.50, r_gamma = 1.20)  # upper bound

# Run initial coverage analysis
set.seed(n_seed)  # set seed
m_params_1 <- sample_prior(n_samp = n_sim,
                           v_lb   = v_lb_1,
                           v_ub   = v_ub_1)

l_coverage_1  <- run_coverage(l_params_all = l_params_all, m_params = m_params_1)
df_coverage_1 <- l_coverage_1$df_model_summ %>%
  mutate(type_coverage = "Initial model coverage")

l_plot_coverage_1 <- plot_coverage(df_targets  = df_targets,
                                   df_coverage = df_coverage_1)

l_plot_coverage_1

## Resize the prior distribution bounds ----------------------------------
### Adjust transmission rate: r_beta -------------------------------------
# 1. Compute model-data residuals on initial coverage (mean values)
v_params_1 <- colMeans(m_params_1)
modCost1   <- costFun(v_params     = v_params_1,
                      l_params_all = l_params_all,
                      v_dates      = v_dates)

# Plot residuals
# plot_costFun(Cfun = modCost1, wgt_res = T) # weighted residuals
# plot_costFun(modCost1, wgt_res = F)        # unweighted residuals

# 2. Local sensitivity analysis
SensRes1 <- sensFun(func         = costFun,
                    parms        = v_params_1,
                    l_params_all = l_params_all,
                    v_dates      = v_dates)

# Plot sensitivity
plot_Sfun(SensRes1)

# Rank parameters according to their importance
m_Sens1 <- summary(SensRes1)

# value: value of the parameter
# Mean: mean sensitivity
# L1: L1 norm
# L2: L2 norm
df_Sens1 <- as.data.frame(m_Sens1)

# Order by L2
df_Sens1[order(df_Sens1$L2, decreasing = T),]

# 3. Compute the change in the prior distribution bounds based on the
# sensitivity analysis
df_changes_1 <- data.frame(NULL)
for(target_i in c("DXIncTot")){ # target_i = "DXIncTot"

  # Filter elasticities
  df_epsi <- SensRes1 %>%
    filter(var == target_i) %>%
    select(x,r_beta) %>%
    rename(epsilon = r_beta)

  # Change in y --> change in par
```

```
  df_temp <- modCost1$residuals %>%
    filter(name == target_i) %>%
    select(name,x,obs,mod) %>%
    mutate(y_change = ifelse(mod != 0, (obs-mod)/mod, 0)) %>%
    left_join(df_epsi,
              by = "x") %>%
    mutate(par_change = ifelse(epsilon == 0, NA, y_change/epsilon),
           par_change_lb = ifelse(par_change < 0,
                                  NA, (1+par_change)*v_lb_1["r_beta"]),
           par_change_ub = ifelse(par_change > 0 |
                                    (1+par_change)*v_ub_1["r_beta"] < 0.3,
                                  NA, (1+par_change)*v_ub_1["r_beta"]))

  # Bind data
  df_changes_1 <- rbind(df_changes_1,
                        df_temp)

}

# Change for lower or upper bound
n_change_lb <- mean(df_changes_1$par_change_lb, na.rm = T)
n_change_ub <- mean(df_changes_1$par_change_ub, na.rm = T)

# Resize bounds
v_lb_2 <- v_lb_1
v_ub_2 <- v_ub_1
v_lb_2["r_beta"] <- n_change_lb
v_ub_2["r_beta"] <- n_change_ub

# Run coverage
set.seed(n_seed)  # set seed
m_params_2 <- sample_prior(n_samp = n_sim,
                           v_lb = v_lb_2,
                           v_ub = v_ub_2)
l_coverage_2 <- run_coverage(l_params_all = l_params_all,
                             m_params     = m_params_2)
df_coverage_2 <- l_coverage_2$df_model_summ
df_coverage_2$type_coverage <-
  "A: Improved model coverage by resizing the transmission rate"

plot_coverage(df_targets  = df_targets,
              df_coverage = rbind(df_coverage_1,df_coverage_2))

### Adjust recovery rate: r_gamma -----------------------------------------
# 1. Compute model-data residuals on previous coverage (mean values)
v_params_2 <- colMeans(m_params_2)        # mean values
modCost2   <- costFun(v_params     = v_params_2,
                      l_params_all = l_params_all,
                      v_dates      = v_dates)

# 2. Local sensitivity analysis
SensRes2 <- sensFun(func         = costFun,
                    parms        = v_params_2,
                    l_params_all = l_params_all,
                    v_dates      = v_dates)

# Plot sensitivity
plot_Sfun(SensRes2)

# Rank parameters according to their importance
m_Sens2 <- summary(SensRes2)
```

```
# value: value of the parameter
# Mean: mean sensitivity
# L1: L1 norm
# L2: L2 norm
df_Sens2 <- as.data.frame(m_Sens2)

# Order by L2
df_Sens2[order(df_Sens2$L2, decreasing = T),]

# 3. Compute the change in the prior distribution bounds based on the
# sensitivity analysis
df_changes_2 <- data.frame(NULL)
for(target_i in c("DXIncTot")){ # target_i = "DXIncTot"

  # Filter elasticities
  df_epsi <- SensRes2 %>%
    filter(var == target_i) %>%
    select(x,r_gamma) %>%
    rename(epsilon = r_gamma)

  # Change in y --> change in par
  df_temp <- modCost2$residuals %>%
    filter(name == target_i) %>%
    select(name,x,obs,mod) %>%
    mutate(y_change = ifelse(mod != 0, (obs-mod)/mod, 0)) %>%
    left_join(df_epsi,
              by = "x") %>%
    mutate(par_change = ifelse(epsilon == 0, NA, y_change/epsilon),
           par_change_lb = ifelse(par_change < 0,
                                  NA, (1+par_change)*v_lb_2["r_gamma"]),
           par_change_ub = ifelse(par_change > 0 |
                                    (1+par_change)*v_ub_2["r_gamma"] < 0.3,
                                  NA, (1+par_change)*v_ub_2["r_gamma"]))

  # Bind data
  df_changes_2 <- rbind(df_changes_2,
                        df_temp)

}

# Change for lower or upper bound
n_change_lb <- mean(df_changes_2$par_change_lb, na.rm = T)
n_change_ub <- mean(df_changes_2$par_change_ub, na.rm = T)

# Resize bounds
v_lb_3 <- v_lb_2
v_ub_3 <- v_ub_2
v_lb_3["r_gamma"] <- n_change_lb
v_ub_3["r_gamma"] <- n_change_ub

# Run coverage
set.seed(n_seed)  # set seed
m_params_3 <- sample_prior(n_samp = n_sim,
                           v_lb   = v_lb_3,
                           v_ub   = v_ub_3)
l_coverage_3 <- run_coverage(l_params_all = l_params_all,
                             m_params     = m_params_3)
df_coverage_3 <- l_coverage_3$df_model_summ
df_coverage_3$type_coverage <-
  "B: A + improved model coverage by resizing the recovery rate"

plot_coverage(df_targets  = df_targets,
```

```
              df_coverage = rbind(df_coverage_1,
                                  df_coverage_2,
                                  df_coverage_3))

### Readjust transmission rate: r_beta ------------------------------------
# 1. Compute model-data residuals on previous coverage (mean values)
v_params_3 <- colMeans(m_params_3)
modCost3   <- costFun(v_params     = v_params_3,
                      l_params_all = l_params_all,
                      v_dates      = v_dates)

# 2. Local sensitivity analysis
SensRes3 <- sensFun(func         = costFun,
                    parms        = v_params_3,
                    l_params_all = l_params_all,
                    v_dates      = v_dates)

# Plot sensitivity
plot_Sfun(SensRes3)

# Rank parameters according to their importance
m_Sens3 <- summary(SensRes3)

# value: value of the parameter
# Mean: mean sensitivity
# L1: L1 norm
# L2: L2 norm
df_Sens3 <- as.data.frame(m_Sens3)

# Order by L2
df_Sens3[order(df_Sens3$L2, decreasing = T),]

# 3. Compute the change in the prior distribution bounds based on the
# sensitivity analysis
df_changes_3 <- data.frame(NULL)
for(target_i in c("DXIncTot")){ # target_i = "DXIncTot"

  # Filter elasticities
  df_epsi <- SensRes3 %>%
    filter(var == target_i) %>%
    select(x,r_beta) %>%
    rename(epsilon = r_beta)

  # Change in y --> change in par
  df_temp <- modCost3$residuals %>%
    filter(name == target_i) %>%
    select(name,x,obs,mod) %>%
    mutate(y_change = ifelse(mod != 0, (obs-mod)/mod, 0)) %>%
    left_join(df_epsi,
              by = "x") %>%
    mutate(par_change = ifelse(epsilon == 0, NA, y_change/epsilon),
           par_change_lb = ifelse(par_change < 0,
                                  NA, (1+par_change)*v_lb_3["r_beta"]),
           par_change_ub = ifelse(par_change > 0 |
                                    (1+par_change)*v_ub_3["r_beta"] < 0.3,
                                  NA, (1+par_change)*v_ub_3["r_beta"]))

  # Bind data
  df_changes_3 <- rbind(df_changes_3,
                        df_temp)

}
```

```r
# Change for lower or upper bound
n_change_lb <- mean(df_changes_3$par_change_lb, na.rm = T)
n_change_ub <- mean(df_changes_3$par_change_ub, na.rm = T)

# Resize bounds
v_lb_4 <- v_lb_3
v_ub_4 <- v_ub_3
v_lb_4["r_beta"] <- n_change_lb
v_ub_4["r_beta"] <- n_change_ub

# Run coverage
set.seed(n_seed)  # set seed
m_params_4 <- sample_prior(n_samp = n_sim,
                           v_lb = v_lb_4,
                           v_ub = v_ub_4)
l_coverage_4 <- run_coverage(l_params_all = l_params_all,
                             m_params     = m_params_4)
df_coverage_4 <- l_coverage_4$df_model_summ
df_coverage_4$type_coverage <-
  "C: A + B + improved model coverage by resizing the transmission rate"

plot_coverage(df_targets  = df_targets,
              df_coverage = rbind(df_coverage_1,
                                  df_coverage_2,
                                  df_coverage_3,
                                  df_coverage_4))

# Range on input search space after resize bounds
v_lb_final <- v_lb_4
v_ub_final <- v_ub_4

# Final coverage
set.seed(n_seed)  # set seed
m_params_final <- sample_prior(n_samp = n_sim,
                               v_lb   = v_lb_final,
                               v_ub   = v_ub_final)

l_coverage_final <- run_coverage(l_params_all = l_params_all,
                                 m_params     = m_params_final)
df_coverage_final <- l_coverage_final$df_model_summ
plot_model_out_vs_targets(df_targets = df_targets,
                          df_model   = df_coverage_final)

## Identifiability (Collinearity) analysis --------------------------------
# Compute model-data residuals on previous coverage (mean values)
v_params <- colMeans(m_params_final)        # mean values

# Sensitivity analysis
SensRes <- sensFun(func         = costFun,
                   parms        = v_params,
                   l_params_all = l_params_all,
                   v_dates      = v_dates)

df_collin <- collin(SensRes) %>%
  mutate(var     = "All",
         Outcome = paste0(v_outcome_names,collapse = " & "))

# Plot collinearity analysis
plot_CollIndx(df_collin = df_collin, flag_each = F, log_y = T)

## Calibrate ------------------------------------------------------------
# Range on input search space
```

```r
v_lb <- v_lb_final # lower bound
v_ub <- v_ub_final # upper bound

# Specify seed (for reproducible sequence of random numbers)
set.seed(n_seed)

# number of random samples
n_resamp <- 10000

# record start time of calibration
t_init <- Sys.time()

# Bayesian calibration using the IMIS algorithm (Raftery & Bao, 2010)
fit_imis <- IMIS(B             = 1000,      # incremental sample size at each iteration of IMIS
                 B.re          = n_resamp,  # desired posterior sample size
                 number_k      = 20,        # maximum number of iterations in IMIS
                 D             = 0,
                 v_lb          = v_lb,
                 v_ub          = v_ub,
                 l_params_all = l_params_all,
                 v_dates       = v_dates)

# Calculate computation time
comp_time <- Sys.time() - t_init
comp_time

# obtain draws from posterior
m_calib_res <- fit_imis$resample

# Calculate log-likelihood (overall fit) and posterior probability of each sample
m_calib_res <- cbind(m_calib_res,
                     "Overall_fit"    = log_lik(m_calib_res[,v_param_names],
                                                l_params_all = l_params_all,
                                                v_dates      = v_dates),
                     "Posterior_prob" = posterior(m_calib_res[,v_param_names],
                                                  v_lb         = v_lb,
                                                  v_ub         = v_ub,
                                                  l_params_all = l_params_all,
                                                  v_dates      = v_dates))

# Normalize posterior probability
m_calib_res[,"Posterior_prob"] <- m_calib_res[,"Posterior_prob"]/
  sum(m_calib_res[,"Posterior_prob"])

### Exploring best-fitting input sets -------------------------------------
# Plot the 1000 draws from the posterior with marginal histograms
pairs.panels(m_calib_res[,v_param_names])

# Compute posterior mean
v_calib_post_mean <- colMeans(m_calib_res[,v_param_names])
v_calib_post_mean

# Compute posterior median and 95% credible interval
m_calib_res_95cr <- colQuantiles(m_calib_res[,v_param_names],
                                 probs = c(0.025, 0.5, 0.975))
m_calib_res_95cr

### Plot maximum-a-posteriori ---------------------------------------------
# Compute MAP parameter set
v_calib_map <- m_calib_res[which.max(m_calib_res[,"Posterior_prob"]),]

# Run model with MAP
```

```r
l_out_best <- SIR_calibration_out(v_params     = v_calib_map[v_param_names],
                                  l_params_all = l_params_all)

# Plot
plotrix::plotCI(x = df_targets$Day, y = df_targets$value,
                ui = df_targets$ub,
                li = df_targets$lb,
                ylim = c(0, 800),
                xlab = "Time (days)", ylab = "Number of infections")
points(x = l_out_best$DXIncTot$Day,
       y = l_out_best$DXIncTot$value,
       pch = 8, col = "red")
legend("topright",
       legend = c("Target", "Model-predicted output"),
       col = c("black", "red"), pch = c(1, 8))

### Propagate calibrated parameter uncertainty ----------------------------
# Compute IMIS posterior predicted outputs
m_out <- matrix(NA, nrow = n_resamp, ncol = nrow(df_targets))

# Run model for each posterior parameter set
for(i in 1:n_resamp){ # i = 1
  l_out_model <- SIR_calibration_out(v_params     = m_calib_res[i, ],
                                     l_params_all = l_params_all)
  df_out_model <- l_out_model$DXIncTot
  m_out[i, ] <- df_out_model$value
  if(i/100==round(i/100,0)) {
    cat('\r',paste(i/n_resamp*100,"% done",sep=""))
  }
}

# Posterior predicted mean
m_out_postmean <- colMeans(m_out)

# Implement PRE-CISE (narrow) --------------------------------------------
# 0. Define input search space and plot first coverage
v_lb_1 <- c(r_beta = 0.30, r_gamma = 0.60)  # lower bound
v_ub_1 <- c(r_beta = 1.00, r_gamma = 0.90)  # upper bound

# Run initial coverage analysis
set.seed(n_seed) # set seed
m_params_1 <- sample_prior(n_samp = n_sim,
                           v_lb   = v_lb_1,
                           v_ub   = v_ub_1)

l_coverage_1  <- run_coverage(l_params_all = l_params_all,
                              m_params     = m_params_1)
df_coverage_1 <- l_coverage_1$df_model_summ %>%
  mutate(type_coverage = "Initial model coverage")

plot_coverage(df_targets  = df_targets,
              df_coverage = df_coverage_1)

## Resize the prior distribution bounds -----------------------------------
### Adjust transmission rate: r_beta --------------------------------------
# 1. Compute model-data residuals on initial coverage (mean values)
v_params_1 <- colMeans(m_params_1)
modCost1   <- costFun(v_params     = v_params_1,
                      l_params_all = l_params_all,
                      v_dates      = v_dates)

# Plot residuals
```

```r
plot_costFun(Cfun = modCost1, wgt_res = T) # weighted residuals
plot_costFun(modCost1, wgt_res = F)        # unweighted residuals

# 2. Local sensitivity analysis
SensRes1 <- sensFun(func         = costFun,
                    parms        = v_params_1,
                    l_params_all = l_params_all,
                    v_dates      = v_dates)

# Plot sensitivity
plot_Sfun(SensRes1)

# Rank parameters according to their importance
m_Sens1 <- summary(SensRes1)

# value: value of the parameter
# Mean: mean sensitivity
# L1: L1 norm
# L2: L2 norm
df_Sens1 <- as.data.frame(m_Sens1)

# Order by L2
df_Sens1[order(df_Sens1$L2, decreasing = T),]

# 3. Compute the change in the prior distribution bounds based on the
# sensitivity analysis
df_changes_1 <- data.frame(NULL)
for(target_i in c("DXIncTot")){ # target_i = "DXIncTot"

  # Filter elasticities
  df_epsi <- SensRes1 %>%
    filter(var == target_i) %>%
    select(x,r_beta) %>%
    rename(epsilon = r_beta)

  # Change in y --> change in par
  df_temp <- modCost1$residuals %>%
    filter(name == target_i) %>%
    select(name,x,obs,mod) %>%
    mutate(y_change = ifelse(mod != 0, (obs-mod)/mod, 0)) %>%
    left_join(df_epsi,
              by = "x") %>%
    mutate(par_change = ifelse(epsilon == 0, NA, y_change/epsilon),
           par_change_ub = ifelse(par_change < 0 |
                                    (1+par_change)*v_ub_1["r_beta"]>5,
                                  NA, (1+par_change)*v_ub_1["r_beta"]))

  # Bind data
  df_changes_1 <- rbind(df_changes_1,
                        df_temp)

}

# Change for lower or upper bound
n_change_ub <- mean(df_changes_1$par_change_ub, na.rm = T)

# Resize bounds
v_lb_2 <- v_lb_1
v_ub_2 <- v_ub_1
v_ub_2["r_beta"] <- n_change_ub

# Run coverage
```

```
set.seed(n_seed) # set seed
m_params_2 <- sample_prior(n_samp = n_sim,
                           v_lb = v_lb_2,
                           v_ub = v_ub_2)
l_coverage_2 <- run_coverage(l_params_all = l_params_all, m_params = m_params_2)
df_coverage_2 <- l_coverage_2$df_model_summ
df_coverage_2$type_coverage <-
  "A: Improved model coverage by resizing the transmission rate"

plot_coverage(df_targets  = df_targets,
              df_coverage = rbind(df_coverage_1,
                                  df_coverage_2))

### Adjust recovery rate: r_gamma -----------------------------------------
# 1. Compute model-data residuals on previous coverage (mean values)
v_params_2 <- colMeans(m_params_2)        # mean values
modCost2   <- costFun(v_params     = v_params_2,
                      l_params_all = l_params_all,
                      v_dates      = v_dates)

# 2. Local sensitivity analysis
SensRes2 <- sensFun(func         = costFun,
                    parms        = v_params_2,
                    l_params_all = l_params_all,
                    v_dates      = v_dates)

# Plot sensitivity
plot_Sfun(SensRes2)

# Rank parameters according to their importance
m_Sens2 <- summary(SensRes2)

# value: value of the parameter
# Mean: mean sensitivity
# L1: L1 norm
# L2: L2 norm
df_Sens2 <- as.data.frame(m_Sens2)

# Order by L2
df_Sens2[order(df_Sens2$L2, decreasing = T),]

# 3. Compute the change in the prior distribution bounds based on the
# sensitivity analysis
df_changes_2 <- data.frame(NULL)
for(target_i in c("DXIncTot")){ # target_i = "DXIncTot"

  # Filter elasticities
  df_epsi <- SensRes2 %>%
    filter(var == target_i) %>%
    select(x,r_gamma) %>%
    rename(epsilon = r_gamma)

  # Change in y --> change in par
  df_temp <- modCost2$residuals %>%
    filter(name == target_i) %>%
    select(name,x,obs,mod) %>%
    mutate(y_change = ifelse(mod != 0, (obs-mod)/mod, 0)) %>%
    left_join(df_epsi,
              by = "x") %>%
    mutate(par_change = ifelse(epsilon == 0, NA, y_change/epsilon),
           par_change_lb = ifelse(par_change > 0 |
                                  (1+par_change)*v_lb_2["r_gamma"] < 0,
```

```
                                  NA, (1+par_change)*v_lb_2["r_gamma"]))

  # Bind data
  df_changes_2 <- rbind(df_changes_2,
                        df_temp)

}

# Change for lower or upper bound
n_change_lb <- min(df_changes_2$par_change_lb, na.rm = T)

# Resize bounds
v_lb_3 <- v_lb_2
v_ub_3 <- v_ub_2
v_lb_3["r_gamma"] <- n_change_lb

# Run coverage
set.seed(n_seed) # set seed
m_params_3 <- sample_prior(n_samp = n_sim,
                           v_lb   = v_lb_3,
                           v_ub   = v_ub_3)
l_coverage_3 <- run_coverage(l_params_all = l_params_all, m_params = m_params_3)
df_coverage_3 <- l_coverage_3$df_model_summ
df_coverage_3$type_coverage <-
  "B: A + improved model coverage by resizing the recovery rate"

plot_coverage(df_targets  = df_targets,
              df_coverage = rbind(df_coverage_1,
                                  df_coverage_2,
                                  df_coverage_3))

### Readjust transmission rate: r_beta ------------------------------------
# 1. Compute model-data residuals on previous coverage (mean values)
v_params_3 <- colMeans(m_params_3)
modCost3   <- costFun(v_params     = v_params_3,
                      l_params_all = l_params_all,
                      v_dates = v_dates)

# 2. Local sensitivity analysis
SensRes3 <- sensFun(func         = costFun,
                    parms        = v_params_3,
                    l_params_all = l_params_all,
                    v_dates = v_dates)

SensRes3

# Plot sensitivity
plot_Sfun(SensRes3)

# Rank parameters according to their importance
m_Sens3 <- summary(SensRes3)

# value: value of the parameter
# Mean: mean sensitivity
# L1: L1 norm
# L2: L2 norm
df_Sens3 <- as.data.frame(m_Sens3)

# Order by L2
df_Sens3[order(df_Sens3$L2, decreasing = T),]

# Transmission rate: r_beta
```

```r
# 3. Compute the change in the prior distribution bounds based on the
# sensitivity analysis
df_changes_3 <- data.frame(NULL)
for(target_i in c("DXIncTot")){ # target_i = "DXIncTot"

  # Filter elasticities
  df_epsi <- SensRes3 %>%
    filter(var == target_i) %>%
    select(x,r_beta) %>%
    rename(epsilon = r_beta)

  # Change in y --> change in par
  df_temp <- modCost3$residuals %>%
    filter(name == target_i) %>%
    select(name,x,obs,mod) %>%
    mutate(y_change = ifelse(mod != 0, (obs-mod)/mod, 0)) %>%
    left_join(df_epsi,
              by = "x") %>%
    mutate(par_change = ifelse(epsilon == 0, NA, y_change/epsilon),
           par_change_lb = ifelse(par_change < 0,
                                  NA, (1+par_change)*v_lb_3["r_beta"]),
           par_change_ub = ifelse(par_change > 0 |
                                    (1+par_change)*v_ub_3["r_beta"] < 0.3,
                                  NA, (1+par_change)*v_ub_3["r_beta"]))

  # Bind data
  df_changes_3 <- rbind(df_changes_3,
                        df_temp)

}

# Change for lower or upper bound
n_change_lb <- min(df_changes_3$par_change_lb, na.rm = T)
n_change_ub <- max(df_changes_3$par_change_ub, na.rm = T)

# Resize bounds
v_lb_4 <- v_lb_3
v_ub_4 <- v_ub_3
v_lb_4["r_beta"] <- n_change_lb
v_ub_4["r_beta"] <- n_change_ub

# Run coverage
set.seed(n_seed) # set seed
m_params_4 <- sample_prior(n_samp = n_sim,
                           v_lb = v_lb_4,
                           v_ub = v_ub_4)
l_coverage_4 <- run_coverage(l_params_all = l_params_all, m_params = m_params_4)
df_coverage_4 <- l_coverage_4$df_model_summ
df_coverage_4$type_coverage <-
  "C: A + B + improved model coverage by resizing the transmission rate"

plot_coverage(df_targets  = df_targets,
              df_coverage = rbind(df_coverage_1,
                                  df_coverage_2,
                                  df_coverage_3,
                                  df_coverage_4))

# Range on input search space after resize bounds
v_lb_final <- v_lb_4
v_ub_final <- v_ub_4

# Final coverage
```

```r
set.seed(n_seed)
m_params_final <- sample_prior(n_samp = n_sim,
                               v_lb   = v_lb_final,
                               v_ub   = v_ub_final)

l_coverage_final <- run_coverage(l_params_all = l_params_all,
                                 m_params     = m_params_final)
df_coverage_final <- l_coverage_final$df_model_summ
plot_model_out_vs_targets(df_targets = df_targets,
                          df_model   = df_coverage_final)

## Identifiability (Collinearity) analysis --------------------------------
# Compute model-data residuals on previous coverage (mean values)
v_params <- colMeans(m_params_final)        # mean values

# Sensitivity analysis
SensRes <- sensFun(func         = costFun,
                   parms        = v_params,
                   l_params_all = l_params_all,
                   v_dates      = v_dates)

df_collin <- collin(SensRes) %>%
  mutate(var     = "All",
         Outcome = paste0(v_outcome_names,collapse = " & "))

# Plot collinearity analysis
plot_CollIndx(df_collin = df_collin, flag_each = F, log_y = T)
```
